# DISCOVERY OF BREAST CANCER AND AUTISM CAUSES Method of combining multiple researches to determine non-infectious disease causes

**DOI:** 10.1101/2024.06.10.24308732

**Authors:** Alan Olan

## Abstract

In this work an author analyses causes of Breast Cancer and Autism using a novel method presented in article “Method of Combining Multiple Researches to Determine Non-Infectious Disease Causes, Analysis of Depression and Celiac Disease Causes”(Alan Olan, Biology and Medicine,vol.16 Iss.1 No: 1000635). The method is using a special **algorithm based in math which allows to find disease causes for a specific non-infectious disease using results of multiple researches** regarding risk factors of the disease. This method allows to combine dozens of such researches together with dozens of researches on biochemistry and physiology to extract implicit information on the causes of non-infectious disease out this entire research.

The use of method requires to find number of causes for a specific non-infectious disease using a special formula and data on the incidence rate of disease in specific population. These disease causes are two or more physiological changes beyond approximately 1-sigma interval which if they are co-existing long enough must trigger the non-infectious disease (triggering is not optional). After that the method requires to find disease causation factors out of multiple risk factors found for a disease. Only some of risk factors are real disease causing factors. This is achieved using a set of special disease causation criteria discussed in the article above and provided here for a reference as well.

The method often allows to find few dozen of disease causation factors and more for a non-infectious disease out of existing medical research. These disease causation factors all point to the same set of limited number of physiological parameters changes beyond 1-sigma. The number of these physiological changes usually vary depending on the non-infectious disease from a minimum of 2 to a maximum of 6 or very rare more. In this case the few dozens of disease causation factors make changes (we can say “point”) to the same set of 3 physiological parameters(for example).The method then allows to find these physiological parameter changes (which are real cause of the non-infectious disease) using a properties based in math.

After this the method allows to determine which physiological parameter of this group is impacted by each of the dozens of disease causation factors previously found. Then method allows to group these factors according to the physiological parameter they impact. The disease causation factors taken out of each group of these factors and combined together will cause a change beyond 1-sigma to all required for disease triggering physiological parameters. **These combinations of disease causation factors applied long enough will cause a non-infectious disease**. The occurrence of the disease causation factors is random but once they act together the non-infectious disease triggering is a must unless the factors are removed fast enough. Final step of the method is validation of its results using other research or the already discussed disease causation criteria in order to eliminate any errors in steps of the method which we could potentially make.

Once the simultaneously taking place physiological changes causing a non-infectious disease has been found the method allows to build a hypothesis of the disease pathology by using them and “connecting the dots”. The example of this process shown in the work as well. The hypothesis of Autism pathology is proposed as one example of this.

Using this method and applying over ***29 existing selected studies*** at the same time an author analyses Breast Cancer and as a result the work gives the causes of the Breast Cancer disease as a set of physiological parameters changes beyond 1-sigma interval (slightly less, actually) and also as a set of disease causing external factors which combinations in an individual must cause Breast Cancer as per presented model. Using the method an author also shows that Breast cancer has 4 simultaneously acting causes (physiological parameter changes).

The author then continues an introduction to the method and applying over ***34 existing selected studies*** the author analyses Autism and as a result the work gives the causes of Autism as a set of physiological parameters changes beyond 1-sigma interval (slightly less, actually) and also as a set of disease causing external factors which combinations in an individual must cause Autism. Using the method an author show that Autism has 2 simultaneously acting causes for boys and 3 simultaneously acting causes for girls at present time. For Autism, this explains why girls’ rate of Autism much less than in boys because the more physiological changes are required to trigger the non-infectious disease the less its incidence rate [1].It used to be more physiological changes required to cause Autism in the past, in 1980s and earlier. But some external factors has removed multiple defensive mechanisms by moving some physiological parameters beyond 1-sigma to a new “norm” and left us with only 2 for boys and 3 for girls as it is shown in this work. *This removal of defenses has caused a significant raise of Autism in recent years*.

The article introduces to the basics of the method, provides required formulas for calculations and then move to a detailed analysis of these two non-infectious diseases. As the method is novel the appendix has an analogy to explain the idea of the method at “high level”. The author’s introduction to the method will allow other medical researchers to use their own and existing research to determine the causes of non-infectious diseases as per presented model, using a simple algorithm.

## Introduction

In article [1] *“A Connection between Factors Causing Diseases and Diseases Frequencies: Its Application in Finding Disease Causes” (Alan Olan*, Journal of Clinical Trials, Vol.13, Issue 4.) we introduced a model of non-infectious disease. According to this model which is matching to empirical evidence, the non-infectious disease is caused by changes to multiple physiological parameters when their values go beyond 1-sigma interval, slightly less actually. This means a non-infectious disease must occur if 2 or more particular physiological parameters changes beyond 1-sigma exist for while at the same time. Also, based on the model the criteria was introduced to determine if a risk factor is causing a non-infectious disease or not. In order to be a cause of non-infectious disease the risk factor **K** calculated as (RR - 1) or (OR-1) to be **3.55+/-50%** (or 355%+/-50%) if the factor is causing a change in **1** physiological parameter, and should have value of **19.67+/-50%** (1967%+/-50%) if it is causing a change to 2 physiological parameters beyond 1-sigma.We call these conditions a **Disease Causation criteria.** The Disease Causation criteria determines whether a factor is one of multiple which are causing disease. The disease causing factor cannot cause a disease as a standalone as per model in article [1] because it is usually changing only 1 physiological parameter out of multiple required. Non-infectious disease, as per the model presented in article [1], is caused by multiple physiological parameters changed beyond 1-sigma.

The article [1] introduced a formula to calculate number of non-infectious disease causes if the disease rate (incidence rate usually) is known for the disease in a specific population. The formula is shown below (**n** is number of disease causes (as number of physiological parameter changed beyond 1-sigma) :

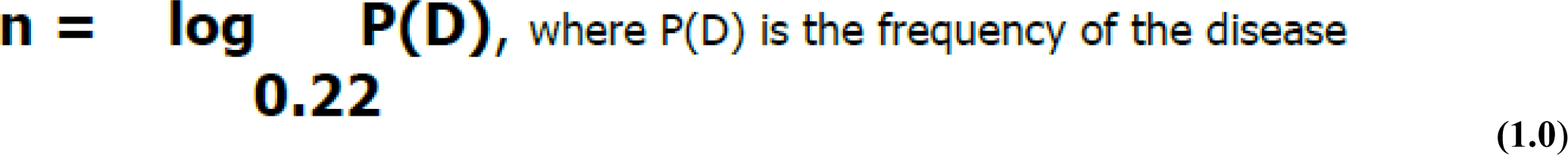

In this formula P(D) is usually represented by **incidence rate** of non-infectious disease (annual disease rate). Using formula 1.0 and Disease Causation criteria it was shown how to find physiological parameters changes which are causing some non-infectious diseases. In this article we introduce a method which is based on this model, which allow researchers to analyze an existing research about a specific non-infectious disease and determine *a full set* of disease causing physiological parameters for a non-infectious disease in much more complicated cases. Also, a method would allow to map a found disease causing factor to a physiological parameter which is changed by the factor beyond 1-sigma interval. For example, if we know that Diabetes is a disease causing factor for Hypertension there is a need to know which specific physiological factor impacted by Diabetes is really a cause of Hypertension. There are multiple physiological factors impacted by Diabetes and the method will allow to find a single one out of so many (allows finding “a needle in a haystack”) which is really causing Hypertension.

The method we introduce is based in math and those who mathematically inclined can find its foundation in the Appendix of this article. Here we will introduce a basic idea of the method and steps on how to practically use it.

## Explanation of the method

A method is introducing an *algorithm* based in math (but *not requiring* to use it much) which allows to process data from results of existing medical researches in few steps and *produce a new information* which consist of disease causes represented by a set of **physiological parameters changes beyond 1-sigma** (slightly less, actually) and also, a few **separate groups of disease causing external factors**. These few groups of factors are such that if you take a disease causing factor from each group as a standalone it will *not* cause a disease but combined together with factors from all other groups *must* cause a non-infectious disease.

Let’s say using the formula for a number of Disease causes (1.0) we found the number of causes as physiological parameter changes beyond 1-sigma. We also found from an existing research a few factors which are really causing a specific disease using a Disease Causation criteria provided. Now we need to determine which specific physiological parameters are really causing a disease. We only know their number and don’t know yet what they are. We know the factors which are causing a change to these physiological parameters and we can use them to find these unknown parameters.

In order to do this we need to list many physiological parameters related to the factor and we can find them from existing scientific research (for example, we can look for all biomarkers of Diabetes and list as many as we can find). Once we list the physiological parameters in the header of some table we need to list the factors found to be causing the disease along a most left column (vertically) of the same table. Based on the researches’ data we already know which physiological parameters are related to those factors causing disease and we mark the intersection between the factor and the related parameter with letter “R” (or other letter). Please note, we use a term “related” as it is unknown if the physiological parameter change is a cause a disease or not yet and all we know is that its change is present if the factor is present. Now we start analyzing which factors are having same parameter in common and we will call it **an *intersection* if unrelated disease causing factors A and B both having the same physiological parameter changed.** We need to find all these intersections. Those intersections will be a superset of the physiological parameters which are causing a disease. The details of why this happens are provided in a detailed math explanation in this article. Here we can say that a match in just one physiological parameter changed (*important*: we looking for **a match by name**, not by value) between 2 different factors is not a random coincidence but a pattern. This pattern is determined by a fact that factors are causing a change of the physiological parameter beyond 1-sigma interval (or sometimes by other reasons explained later).

Here is *a simplified* explanation how the method works. Suppose there is a non-infectious disease with 2 unknown disease causes (quantity of 2 we have determined using formula 1.0). Suppose we determined already 2-3 disease causing factors (we can do it using Disease Causation criteria).Each disease causing factor is changing physiological parameters randomly beyond *their original(or normal) range* which means a quantity of physiological parameters changes is random, the **parameters’ names**, values are different as well, etc. The physiological parameters changes for a disease causing factors are random as they cannot be determined unless we do an experiment and determine what they have changed under influence of each of these disease causing factors (we assume we do an experiment first time). Note, the physiological parameter changes are *random for a disease causing factor* but they may be almost *the same for different people* impacted by this factor.

Suppose via the experiments we found that a disease causing factor 1 has changed 20 different physiological parameters beyond their original (or normal) range and a disease causing factor 2 has changed 50 different physiological parameters beyond their original (or normal) range. We just explained that *only 2 unknown* physiological parameters are really causing this disease. In order to determine **their names** we use a fact that due to random “selection” of physiological parameters out of so many (few thousands and more exist in human body) by ***unrelated*** *disease causing* factors, a match by **their names** and values between these 2 groups of physiological parameters practically is not possible by accident (this is shown mathematically in this article). If we find **a match of physiological parameters by name it is a pattern** under these specified conditions, it is not random. For example, if one type of *disease causing* factors (suppose a chocolate consumption) is causing a change in hemoglobin levels and a totally different *disease causing* factor (suppose a long exercising) is causing a change in hemoglobin levels as well then it is a pattern *under these specified conditions*.It becomes a pattern which we are interested in *because* it happens between groups belonging to *2 different disease causing* factors. We know these 2 *disease causing factors* are impacting same physiological parameter, meaning a parameter with the **same name**. For this reason, in order to find a pattern (in our case a disease causation physiological parameter) we need to find a place where 2 physiological parameters changes in both groups are the **same by name** (*important*: not by value) for these 2 different disease causing factors. This place where 2 disease causing factors have **the same name** of physiological parameter changed beyond original(or normal range) we call “an intersection”. If we found such a match between physiological parameters **by name** (*important:* not by value) we found a pattern - a physiological parameter which is causing a disease. See a pic 1. Also, in Appendix 1 we provide a simple analogy which could help to clarify or visualize the explained concept further.

**Pic 1.**
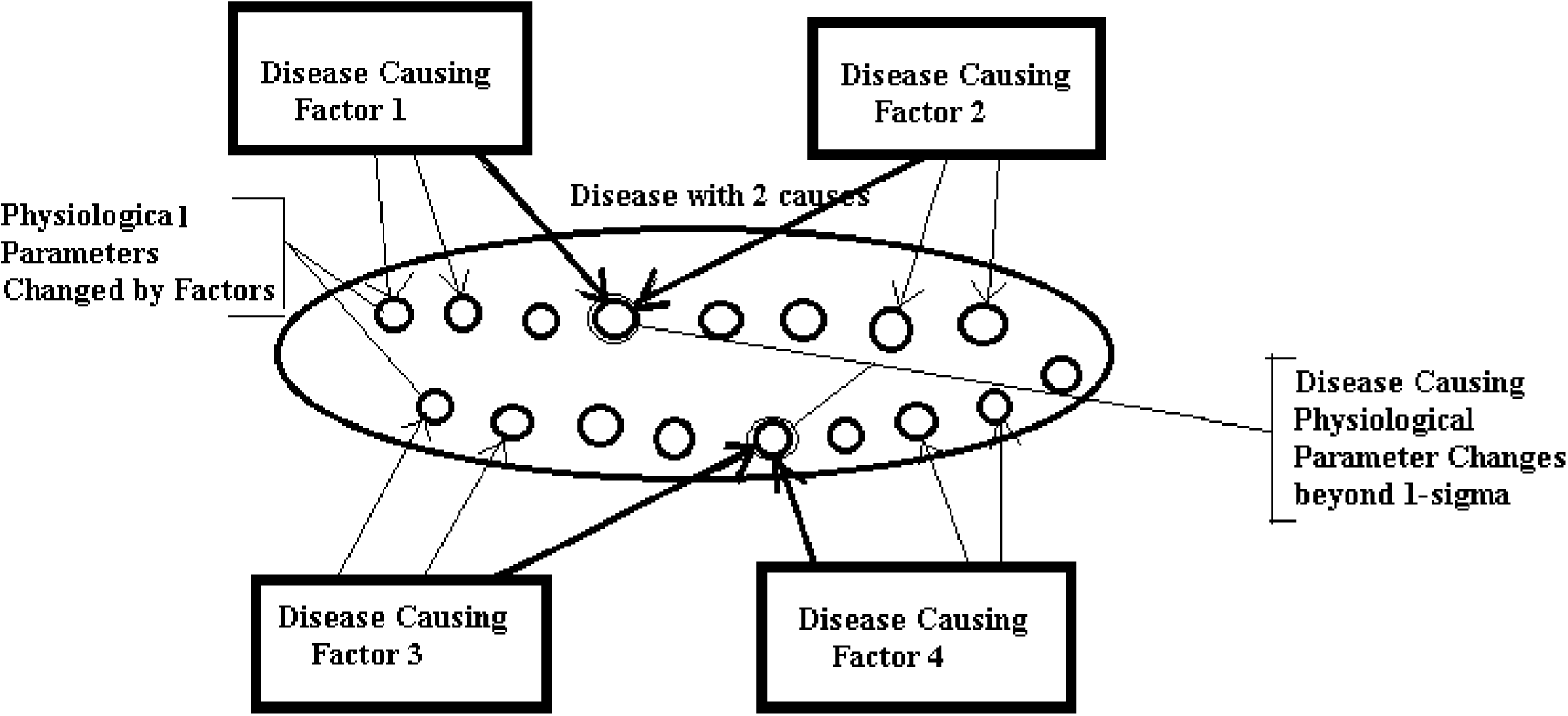
(Disease with 2 causes, as per formula (1.0): Disease causing factors are causing multiple physiological changes. **Disease causing** physiological changes are common for Factors 1 and 2, for Factors 3 and 4.We find them by determining an intersection in a physiological parameter)

When we find all these intersections we will find a list (or a matrix) of the parameters which are superset of physiological parameters which changes go beyond 1-sigma and are causing a disease. As there might be other patterns causing a match in parameters the number of parameters can exceed the number of parameters we determined via Number of Disease Causes formula (1.0). We will need to eliminate other parameters based on the methods provided in this article to get a final set of Physiological Parameters which changes go beyond 1-sigma (actually slightly less).These changes when they are present at the same time *must* trigger a disease. The illustration of this process shown on Pic 1.

Once we know the final set of these physiological parameters and as we know up front which factor is related to which physiological parameter we can find which specific physiological parameter is impacted by the specific factor in such a way that a parameter goes beyond 1-sigma interval. In other words we get an understanding that a factor is impacting a specific physiological parameter. Now, if an individual is affected by a set of factors which are causing a change in ALL these physiological parameters required then the individual will get sick after sometime unless the harmful factors are removed fast enough. This knowledge can help to prevent a disease in the individual or help potentially cure it or reduce severity of the disease.

## Using a method in practice

In a nutshell, in order to use this method to find a set of physiological parameters which are causing a disease next steps should be taken:

1. Find the number of causes (as physiological parameters changes) for the non-infectious disease using a formula for a disease causes *(1.0) (see also our article:, **“*****A Connection between Factors Causing Diseases and Diseases Frequencies: Its Application in Finding Disease Causes”**, *Alan Olan*, Journal of Clinical Trials, Vol.13, Issue 4.)
2. Find as many as possible, the factors which are causing a disease using a Disease Causation criteria on existing experimental data or new research (*see our article: **“*****A Connection between Factors Causing Diseases and Diseases Frequencies: Its Application in Finding Disease Causes”**, *Alan Olan*, Journal of Clinical Trials, Vol.13, Issue 4.)
3. For each factor found, you need to list as many as possible parameters related to it in some table. For example, if a factor is a consumption of some food then include which nutrients, chemicals it contains including harmful ones as per existing researches. If a factor is a disease then list which physiological parameters are known to be impacted one way or another as a symptom, a cause, etc. You need to place one factor vertically and appropriate impacted physiological parameters horizontally(in the table’s header) in this table. The column where a factor is related to a physiological parameter can be marked with cross “X” or letter “R”.
4. If the factor is by itself a physiological parameter it can be taken as one of the physiological parameters you are searching for during this step in the method. It can be corrected with next step if needed.
5. Now, you need to find where the columns which contain physiological parameters are causing a crossing between factors. This means you need to **look for a cross (“X” sign) in the same column for 2 or more factors in the table**. We are basically finding which same (by name, not value) physiological parameter is changed by 2 disease causing factors.
6. Build a list or a matrix **Pm** consisting of the parameters which are causing intersections. For example, like Pm: { r1, r19, r23, r45 }. In practice Pm can look like **Pm = { “Blood Pressure”, “Estrogen Level Change”, “Oxygen level”, “ROS level” }**
7. Eliminate redundant physiological parameters from matrix **Pm** if required (as per rules provided in this article). If the number of calculated parameters per formula (1.0) is less than number of parameters found via analysis of intersections then the redundant parameters need to be eliminated. The number of parameters found via analysis of intersection in a table should match to the number calculated via a formula (1.0) or be less if not all parameters can be found due to a lack of experimental data, etc. Also, if some physiological parameters found as result of analysis of intersection in a table are causing some disease causing factors to change more parameters then predicted by disease causing criteria then some physiological parameters may need to be eliminated as well. The disease causing factor can usually change only 1 or 2 physiological parameters beyond 1-sigma interval (slightly less than this interval, actually).
8. Verify the results of using algorithm by checking if there are appropriate experimental data consistent with the physiological parameters found. Also, you can verify if a physiological parameter found satisfies a Disease Causation criteria above ***if*** there is any research on the risk of this parameter.

You can see that **a method allows to use a simple algorithm** on a set of existing experimental data in order to determine disease causes (as a set of physiological parameter changes beyond 1-sigma interval) and **then verify the results using other research** (which was not used doing steps of the algorithm) either by using Disease Causation criteria or by simply checking how this research is consistent with results of your findings.

## Example of method’s application for a disease with 2 causes

Let’s look at hypothetical disease with 2 causes and apply to it the method described above. We created a Table 1. We can see in **Table 1** that all physiological parameters are listed on the top and all found from the experiments Factors which are causing this hypothetical disease are listed on the left. Now, we can see that parameter **r20** is an intersection for Factor 3 and Factor 4, param **r12** is an intersection for Factor 2 and Factor 3 and Factor 4. A param **r17** is an intersection for Factor 1, Factor 2. We can build our matrix **Pm : { r12, r20, r17 }**.

**Table 1.** (Areas in gray are where an intersections happen in parameters r12, r20 and r17 accordingly. Area surrounded by bold frame is a final set of parameters for matrix Pm)

| Factors causing Disease | Physiological Parameters - Entire Set for ALL FACTORS as R1, R2, ... Rn |  |  |  |  |  |  |  |  |  |  |  |  |  |
| --- | --- | --- | --- | --- | --- | --- | --- | --- | --- | --- | --- | --- | --- | --- |
|  | r12 | r20 | r2 | r8 | r17 | r75 | r34 | r56 | r39 | r12 | r55 | r23 | r78 | r11 |
| Factor 1 |  |  | x |  | x |  |  |  | x | x |  | x |  |  |
| Factor 2 | x |  |  |  | x |  |  | x |  |  | x |  |  | x |
| Factor 3 | x | x |  |  |  |  | x |  |  |  |  |  |  |  |
| Factor 4 | x | x |  |  |  | x |  |  |  |  |  |  | x |  |

In practice, this matrix may look like a list **Pm : { ‘Blood Pressure Change’, ‘Oxygen Level Change’, ‘Estrogen Change’).** This matrix contains physiological parameters which changes beyond 1-sigma are causing a disease if present at the same time.

Now, we see the matrix **Pm : { r12, r20, r17 }** has 3 elements but our disease only require 2. We need to eliminate the incorrect one. We can notice that if we take a valid combination of 2 param as {r12, r17 } then a param r12 is showing that *Factor 2 is impacting r12 and r17 parameters but Factor 2 only can cause 1 parameter* impact (as determined by Disease Causation criteria) so the combination is not correct. If we eliminate r12 and take a combination of {r20, r17 } then both params are satisfying our requirement (assuming there is no info their changes are within 1-sigma). So from this table we can see that final physiological parameter’s set which is causing this disease is **{r20. r17 }**.In this case Factors 3 and 4 are causing a change in parameters **r20**, Factor 1 and 2 are causing a change in physiological parameter **r17**. We has determined 2 physiological causes for a disease with 2 causes.

## Criteria for choosing intersections of physiological parameters

As we could see there is a very small chance of random intersection of physiological parameters belonging to 2 different factors and also, there are cases when more parameters found then determined by disease causation criteria. In order to select those parameters which are really causing a disease we need to use these criteria below.

1. We need to check if factors which are impacting physiological parameters found to be intersections are belonging to **same type of disease** or they are **very similar in nature** (like BMI impact and high weight impact) and we can ignore them as the intersection is most likely due to similarity.
2. Make sure **a factor is causing a change the physiological parameter** which intersect (not opposite) As the factors should cause a change in physiological parameters..
3. Make sure both factors are causing change in the physiological parameter which intersect or the intersection is not valid.
4. If a factor causing insignificant change (less 1-sigma interval) in physiological parameter then the parameters need to be ignored as a point of intersection with this factor. The factors as we discussed need to make change in physiological parameter so its value will be beyond *1-sigma* interval.
5. Make sure the **physiological parameters changes in the point of intersection happen in the same direction** (either increase or decrease). If the changes move in different directions then the parameters are considered as 2 different parameters and not as one.
6. **The intersection of parameters for a similar factor is not considered correct** intersection (as it is the same factor, for example Increased Weigh and BMI over 30 are similar factors)
7. If the experiment found that a factor which causing a disease is impacting 1 physiological parameter but the factor consists of combinations of 2 separate factors (for example observation was done for presence of diabetes or obesity together so they work as a factor) then **the combined factor can be allowed to have 2 intersections despite having impact on 1 physiological parameter** as one of the parts of the factor could be present at the time (for example either a diabetes causing 1 change in physiological parameter or obesity causing the change in another physiological parameter)
8. **Element cannot cross itself**, for example if factor is IgA and physiological parameter IgA then they don’t create a crossing. Usually, if a factor found as physiological parameter then we already know this is a physiological parameter causing a disease.
9. In many cases it is possible to eliminate the redundant intersection by determining via a known research if the physiological parameter increases or decreases with the presence of specific factor. For example, does lipoprotein level decreases or increases with diabetes as causation factor? If same parameter where we see a intersection, increases in presence of one factor and decreases in presence of another then the intersection is invalid.
10. Once a set of physiological parameters was found **they can be also validated with already provided Disease Causation criteria to determine if the factor is a cause of disease** (a parameter should increase a risk of disease 3.5 +/-50% times) using an existing research. For example, if we find that an increased lipoprotein is causing a stroke and if some existing research confirms that a risk of its increase is **3.5 +/-50%** then it confirms a lipoprotein as a parameter was found correctly.
11. In some cases biochemical analysis may help to eliminate an element from matrix Pm

## Algorithm to eliminate redundancy

An approximate algorithm which can be used to eliminate redundant physiological parameters may look like this:

1. Find out if all parameters which intersect in your table satisfy *a hard condition* that a factor only can cause as many changes to physiological parameters as determined via a disease causation criteria (which in most case is 1 and in some 2 or very rare 3).If some physiological parameters are causing not permitted number of changes by a disease causing factor then some parameters need to be eliminated (ignored as intersections).
2. Check the factors which having intersections. If the factors are similar or related then their intersections can be due to similarity and ignored.
3. Select a set of physiological parameters which from a common sense will make most of factors intersect in them. Validate if your selection is following a rule number 1) above. If not use the elimination criteria provided in this article to help resolve conflicts.
4. If 3 factors which are not related intersect in the same physiological parameter then treat a physiological parameter as a high priority intersection as it is extremely low probability that this intersection is not a pattern.
5. If a physiological parameter is selected as a real intersection surround it with a bold rectangle to differentiate from original intersections.
6. Remember that if 2 factors intersect it is a pattern which is determined by next facts 1) that factors are causing a same changes beyond 1-sigma to a physiological parameter **or** 2) the intersection occur due to dependency between related factors or related physiological parameters but the changes are not necessarily beyond 1-sigma. In other words in this case the intersection happens due to this relationships not necessarily due to 1-sigma change (but it can be both at the same time: due to relationship and also due to 1-sigma change).

## METHODOLOGY

### Analysis of Non-Infectious Disease Causes

Below we provide analysis of causes of 2 non-infectious diseases - Breast cancer and Autism as per method presented above. Using these 2 examples we demonstrate in more details how to use the method.

## 1. BREAST CANCER: DETERMINATION OF CAUSES

Breast cancer is the common term for a set of breast tumor subtypes with distinct molecular and cellular origins and clinical behavior. Most of these are epithelial tumors of ductal or lobular origin. Breast cancer happens when there are changes in the genetic material (DNA). Often, the exact cause of these genetic changes is unknown.

In USA (Pennsylvania state) the rate of new breast cancers (incidence rate) between 2016-2020 was 130.6 per 100000 women (CDC, “Rate of New Cancers in the United States”, https://gis.cdc.gov/Cancer/USCS/#/AtAGlance/). Calculating number of causes using formula (1.0) presented above we get:

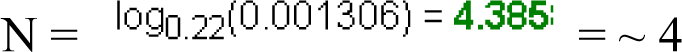

As per calculations above in USA the breast cancer happens due to **4** physiological parameters changes beyond 1-sigma and taking place at the same time. As shown in article [1] despite the different incidence rates in different populations this number will stay the same.

Let’s list the factors which are causing a Breast Cancer and explain how they are determined based on Disease Causation criteria provided above and developed in article ***“*A Connection between Factors Causing Diseases and Diseases Frequencies: Its Application in Finding Disease Causes”** *(Alan Olan,* Journal of Clinical Trials, Vol.13, Issue 4.):

1. A study shows: “The risk of **breast cancer approximately doubled among subjects with blood levels of β-carotene at the lowest quartile**, as compared with those at the highest quartile (**odds ratio = 2.21**; 95% confidence interval (CI): 1.29, 3.79).”, “The **odds ratio for the lower quartile of total carotenoids** was **2.31** (95% CI: 1.35, 3.96). “ (”Serum Carotenoids and Breast Cancer”, Paolo Toniolo, Anne Linda Van Kappel, Arslan Akhmedkhanov, et al, *American Journal of Epidemiology*, Volume 153, Issue 12, 15 June 2001, Pages 1142–1147 ”). We get **OR=2.31** for total carotenoids and this within a range of Disease Causation criteria for one physiological parameter change which is **4.5+/-50%(2.27-6.81).** This means that **low total carotenoids is a disease causation factor which impacts one physiological parameter out of 4** required to cause the Breast Cancer as per Disease Causation criteria. It won’t cause Breast Cancer as standalone factor, it will need other 3 causes out of 4 to be present to cause a Breast Cancer. **It is interesting to notice how long it takes for the breast cancer to develop after observing low carotenes: “**The time delay between the dates of blood sample and diagnosis among the 270 cases ranged between **6 months and 11.2 years** (mean, 4.0 (standard deviation, 2.4) years; median, 3.7 years).”. It shows that to accumulate 3 other required factors causing the Breast Cancer it has required up to 11.2 years ! But if all factors exist it may take as less as about 6 months to develop breast cancer. We place this disease causation factor in Table 1.1 header, column **8.**
2. Another study shows: “Women with **cystic breast disease experience a 2-3 fold increase in risk for breast cancer**” (“Henderson BE, Pike MC, Bernstein L, Ross RK. 1996. *Breast Cancer, chapter 47 in Cancer Epidemiology and Prevention*. 2nd ed. Schottenfeld D and Fraumeni JF Jr.,eds. Oxford University Press. pp: 1022-1035.). If we take **OR=3** then it is within a range of Disease Causation criteria for OR which is **4.5+/-50%(2.27-6.81).** It means that **cystic breast disease is 1 causation factor for Breast Cancer which impacts** one unknown physiological parameter to change beyond 1-sigma interval so it causes Breast Cancer when combined with 3 others out of 4 determined. Cystic breast disease cannot cause Breast Cancer as standalone, it requires 3 other causes to be present at the same time to trigger Breast Cancer in women as per model presented in article [1] by Olan et al.We place this disease causation factor in Table 1.1 header, column **3**
3. Another study shows that **Breast Cancer risk increases with increase in breast density**: “Using women with BI-RADS density code 1 as baseline; women with cod e 2 had RR 1.69 (95%CI 1.56-1.84); women with code 3, RR 2.06 (95%CI 1.89-2.25); and women with code 4, **RR 2.37**(95%CI1.05-2.74).” (*“Breast density and risk of breast cancer”*, Elsebeth Lynge, Ilse Vejborg, Martin Lillholm, et al, Int J Cancer. 2023 Mar 15; 152(6): 1150–1158, doi: 10.1002/ijc.34316). We can see that women with BI-RADS code 4 have **RR = 2.37** which is within Disease Causation criteria **4.5+/-50%(2.27-6.81).** It means that **having density of breast in BI-RADS code 4 is one causation factor of breast cancer**. It mean this is a factor which is impacting one physiological parameter change beyond 1-sigma interval (actually, slightly less). This factor cannot cause a breast cancer as standalone as it impacts only 1 physiological parameter change and for Breast Cancer as we determined 4 physiological changes beyond 1 sigma are required). We add this factor to Table 1.1 header in column **7**.
4. A study shows: “**Among women aged 50–59 years**, with a fat intake in the lowest quartile, **the risk of breast cancer increased with increasing consumption of alcohol**. A consumption of 24 g or more per day was associated **with an 18-fold increased risk** compared with abstainers. For women in other age groups, alcohol consumption had no significant association with breast cancer risk.” *(“Alcohol consumption and breast cancer risk in Denmark.”*, Ewertz, M. Cancer Causes Control 2, 247–252 (1991). https://doi.org/10.1007/BF00052141). We take OR = 18 here, that means risk factor **K** = OR -1 = **17** and for K risk factor the Disease Causation criteria which is related to 2 physiological parameter changes is **19.67+/-50% (9.84-29.47)** as per article [1] so the OR = 18 (K = 17) is within the range for Disease Causation criteria. It means the **alcohol consumption in women under the conditions above is a disease causation factor for Breast Cancer** which impacts **2 physiological parameters** to go beyond approximately 1-sigma interval (actually, it is slightly more narrow interval than 1-sigma). We are adding this factor to Table 1.1 header in column **9**.
5. A study shows: ”…early follicular blood samples from breast cancer cases (*n*=197) in the Nurses Health Study II (NHSII) demonstrated **a significantly elevated breast cancer risk in women with higher total and free estradiol** (RR_Q4 vs. Q1_= 2.1; 95% CI, 1.1–4.1; and, **RR_Q4 vs. Q1_= 2.4;** 95% CI, 1.3–4.5; respectively). Importantly, the magnitude of the effect estimate was more pronounced among the ER+/PR+ cases compared with all breast cancer cases (RR_Q4 vs. Q1_= 2.7; 95% CI, 1.2–6.0 for follicular total estradiol)”*(“Estrogen metabolism and breast cancer.”*, Samavat H, Kurzer MS., Cancer Lett. 2015 Jan 28;356(2 Pt A):231-43. doi: 10.1016/j.canlet.2014.04.018).”. As we can see **RR_Q4 vs. Q1_= 2.4** is within a Disease Causation criteria for 1 physiological change **4.5+/-50%(2.27-6.81).** It means a **high level (4th quartile) of *free estradiol* is a disease causation factor for Breast Cancer** impacting one physiological parameter out of 4.It won’t cause a Breast Cancer as standalone factor but together with other factors (which are impacting other 3 physiological parameters) being present at the same time *must* trigger the disease. We place the **“Free Estradiol”** disease causation factor in the header of Table 1.1, column 1.
6. A study show: “**Inflammatory breast cancer (IBC)** has a rapid, aggressive disease course. A recent case-control study from the Breast Cancer Surveillance Consortium database (1994-2009) showed that **obesity is associated with higher premenopausal IBC risk** (**RR, 3.62**; 95% CI, 1.30-10.04) for all cases and for those with ER-positive **(RR, 3.53**; 95% CI, 1.20-10.39) and ER-negative (**RR, 4.67**; 95% CI, 1.45-15.02) IBC. An older case-comparison study that included 68 IBCs also reported that a **BMI > 26.65 kg/m^2^** was associated with an up **to 4-fold higher risk** of premenopausal IBC”. (“*Obesity and adverse breast cancer risk and outcome: Mechanistic insights and strategies for intervention*.”, Picon-Ruiz M, Morata-Tarifa C,et al, JMCA Cancer J Clin. 2017 Sep;67(5):378-397. doi: 10.3322/caac.21405.). We see that for obesity **RR = 3.62, RR =3.53, RR= 4.67** all satisfy Disease Causation criteria **4.5+/-50%(2.27-6.81)** and it means **that obesity is a causation factor for Inflammatory breast cancer** which impacts 1 physiological change required to cause Breast Cancer out of 4 causes needed. We put this factor in Table 1.1 header, column **11.**
7. A study on free estradiol shows**: “**Data from 2 large studies demonstrated that postmenopausal women in the highest quintile of plasma free *E*_2_ experienced at least a 2.58-fold (95% CI 1.76–3.78) **higher rate of breast cancer** over the ensuing 10 years than those in the lowest quintile.” (“*Effects of estrogen on breast cancer development: Role of estrogen receptor independent mechanisms.*” Yue W, Wang JP, Li Y, Fan P, Liu G, et al. Int J Cancer. 2010 Oct 15;127(8):1748-57. doi: 10.1002/ijc.25207.). We can see that **OR=2.58** is within range of **4.5+/-50%(2.27-6.81)** and so per Disease Causation criteria the **high level of free Estradiol in postmenopausal women is a disease causation factor of Breast Cancer** impacting a change in 1 physiological parameter beyond 1-sigma. We put this factor in Table 1.1 header, column **5.**
8. A study of women women aged 50–79 which were not on hormone replacement therapy(HRT) showed: **“**Among HRT non-users, **heavier women (baseline body mass index (BMI) > 31.1) had an elevated risk of postmenopausal breast cancer (relative risk (RR) = 2.52**; 95% confidence interval (CI) = 1.62–3.93), compared to slimmer women (baseline BMI ≤ 22.6)” (“Obesity, body size, and risk of postmenopausal breast cancer: the Women’s Health Initiative (United States).”, Morimoto, L.M., White, E., Chen, Z. *et al. Cancer Causes Control* 13, 741–751 (2002). https://doi.org/10.1023/A:1020239211145). We see that **RR=2.52** is within of **4.5+/-50%(2.27-6.81)** and this means that **being a heavy women (BMI > 31.1) is a disease causation factor for Breast Cance**r which impact 1-physiological parameter to go beyond 1-sigma interval (actually, slightly less than this). We place this factor in Table 1.1 header, column **2**.
9. A study of rare germline variants in women shows: **“**We found **evidence of association with breast cancer risk for four genes, with estimated adjusted ORs of 5.3** [95% CI: 2.1–16.2] for ***BRCA1***, **4.0** [95% CI: 1.9–9.1] for **BRCA2**, **3.4** [95% CI: 1.4–8.4] for ***ATM*** and **4.3** [95% CI: 1.0–17.0] for ***PALB2***” (*“Population-based estimates of breast cancer risk for carriers of pathogenic variants identified by gene-panel testing”*, Southey MC, Dowty JG, Riaz M, Steen JA et al, NPJ Breast Cancer. 2021 Dec 9;7(1):153. doi: 10.1038/s41523-021-00360-3). All ORs such as **OR=5.3** for BRCA1***, OR=4.0*** *for* BRCA2**, OR = 3.4** for *ATM* **, OR = 4.3** for *PALB2 genes* are within **4.5+/-50%(2.27-6.81)** interval of Disease Causation criteria for 1 physiological parameter change and it means **the mutations in those genes are causation factor for a Breast Cancer affecting 1 physiological parameter** so it goes beyond approximately 1-sigma interval. This unknown parameter is 1 of 4 physiological changes beyond 1-sigma interval which are causing Breast Cancer if co-exist at the same time. *It won’t cause the Breast Cancer standalone though*. We will find those unknown parameters further. We place this genetic related factor in Table 1.1 header, column 6.
10. A study on Estrogen replacement therapy in women reports: “ Current **use of either oral estrogen alone or in combination with progestin pills was associated with increased risk of lobular cancer**, with particularly high risk associated with current combination therapy (**OR, 3.91**; 95% CI, 2.05-7.44). Recent oral HRT use of 57 or more months was associated with **a 3-fold increase in risk of lobular cancer** (**OR, 3.07**; 95% CI, 1.55-6.06) and a 50% increase in nonlobular tumors (OR, 1.52; 95% CI, 1.01-2.29).” (“Hormone Replacement Therapy in Relation to Breast Cancer.”, Chen C, Weiss NS, et al, *JAMA.* 2002;287(6):734–741. doi:10.1001/jama.287.6.734). We see that OR = 3.91 and OR 3.07 are both within a range of **4.5+/-50%(2.27-6.81)** interval of Disease Causation criteria for 1 physiological parameter change. It means that these kinds of estrogen replacement therapy is a causation factor for the breast cancer which changing 1 physiological parameter to go beyond 1-sigma and it is 1 out of 4 parameters required to cause the disease. Estrogen replacement therapy of this kind won’t cause a breast cancer as standalone factor, only with combination of other 3 physiological parameters. We place the found factor in Table 1.1, column **4**.
11. A study on anemia during a breast cancer says: “Multivariate analysis with all relevant prognostic factors in a Cox proportional hazards regression model showed **that preoperative anemia was a significant prognostic factor in breast cancer patients** (Table 4). T-status (≥T_3_), N-status (N_1_, N_2_), strongly positive PR status and HER-2 positivity were significantly associated with LRFS, and **anemic patients had a 4.939-fold increased relative risk of developing local relapse** compared with nonanemic patients.” (”Impact of preoperative anemia on relapse and survival in breast cancer patients.”, Zhang, Y., Chen, Y., Chen, D. *et al. BMC Cancer* 14, 844 (2014). https://doi.org/10.1186/1471-2407-14-844). We see that **RR = 4.939** which is in range of **4.5+/-50%**(**2.27-6.8**1) interval of Disease Causation criteria. It means anemia in breast cancer patients is a causation factor for breast cancer relapse which impact 1 physiological change to be beyond 1-sigma. We place this factor in Table 1.1, column 10.

With this we finished determination of factors which are causing Breast Cancer which we were able to discover in existing research. There can be other factors which are not known to us or which will be discovered in future research. The unknown factors still will impact the same set of physiological parameters which we will determine later in this process.

Now we need to find the physiological parameters **related** to the disease causation factors we found. We don’t know if their changes are causing the disease but we know the changes to these physiological parameters are present then the disease causation factors are present. Let’s list them here:

1. A study of women with a cystic breast disease (with simple and complex cysts) found: “The breast cyst fluid **levels of leptin, adiponectin, and resistin were significantly decreased** compared to plasma in both study groups. Contrarily, levels of **visfatin/NAMPT and TNF-*α* in breast cyst fluid were significantly increased** in relation to plasma in both study groups. In turn, IL-6 levels in breast cyst fluid and plasma were similar in both study groups.”(*“Evaluation of Adipokines, Inflammatory Markers, and Sex Hormones in Simple and Complex Breast Cysts’ Fluid”,* Paweł Madej, Grzegorz Franik, Piotr Kurpas, et al, *Disease Markers*, vol. 2016, Article ID 5174929, 6 pages, 2016. https://doi.org/10.1155/2016/5174929.) We are adding physiological parameters “**Leptin in breast much lower then in Plasma”, “Adiponectin lower”,** and **“Resistin lower”** in most left column of Table 1.1 and add letters “R” for them in **column 3** for Cystic Breast disease and add “R” in the same **column 3** across **“TNF-alfa increase”** parameter.. This will signify relationships between the Cystic disease as a disease causation factor and the physiological parameters changes observed when this factor is present.
2. Additional physiological change for Cystic Disease were reported in a study: “Breast cysts are associated with an increased risk of breast cancer. Some biomarkers such **as estrogen receptor alpha (ERa), progesterone receptor (PR), and cyclin D1, show similar patterns of expression** in epithelial cells lining breast cysts as malignant epithelial cells in local and invasive ductal breast cancer. ” (*“Microcysts and breast cancer: a study of biological markers in archival biopsy material.*”, Tran DD, Lawson JS., Breast Cancer Res Treat. 2002 Oct;75(3):213-20. doi: 10.1023/a:1019969730552. PMID: 12353810.) We place these factors in the most left column of Table 1.1 and put the “R” sign in the **column 3** for a “Cystic Breast Disease” factor.
3. A study on Cystic breast reports: “ We identified **two biomarkers, 15-hydroxyprostaglandin <u>dehydrogenase</u> and 3-hydroxymethylglutaryl-CoA reductase, that were expressed specifically by apocrine type I cysts as well as by apocrine metaplastic cells in type II microcysts,** terminal ducts, and intraductal papillary lesions. No expression of these markers was observed in non-malignant terminal ductal lobular units, type II flat cysts” (*“Apocrine Cysts of the Breast: Biomarkers, Origin, Enlargement, and Relation with Cancer Phenotype”*,Julio E. Celis, Pavel Gromov, José M.A. Moreira, MCP, Volume 5, Issue 3, March 2006, Pages 462-483, https://doi.org/10.1074/mcp.M500348-MCP200). We add these 2 biomarkers to the most left column and add letters “R” across them in column 3 for “Cystic breast disease”
4. Another study on alcohol consumption states: “Muti *et al*. (<u>1998</u>) **measured estradiol level in blood collected from premenopausal women during luteal phase** on the same month, day, hour and minute 1 year apart and **found a significant positive association between estradiol level and alcohol intake** and a higher prevalence of drinkers in subjects with consistently higher estradiol level.” (*“Sex hormones in alcohol consumption: a systematic review of evidence.*“, Erol A, Ho AM, Winham SJ, Karpyak VM., Addict Biol. 2019 Mar;24(2):157-169. doi: 10.1111/adb.12589. Epub 2017 Dec 27. PMID: 29280252; PMCID: PMC6585852.) Also, another study states: “In this large **study of premenopausal women**, we **observed higher luteal estrogen and SHBG concentrations** and lower levels of free testosterone **among women who consumed alcohol compared with nondrinkers.**” (*“Alcohol Consumption in Relation to Plasma Sex Hormones, Prolactin, and Sex Hormone–Binding Globulin in Premenopausal Women.”* Kelly A. Hirko, Donna Spiegelman, Walter C. Willett1, et al, Cancer Epidemiol Biomarkers Prev; 23(12) December 2014). We add **“Estradiol increase”** to left most column (if not added yet) and put the letter “R” in the **column 9** for Alcohol consumption.
5. A study on alcohol consumption by women states: ”… results indicate that moderate **alcohol consumption (15–30 g of alcohol per day) increases serum leptin levels in postmenopausal women**” (*“Relationship Between Serum Leptin Levels and Alcohol Consumption in a Controlled Feeding and Alcohol Ingestion Study”*, Mark J. Roth, David J. Baer, et al, JNCI: Journal of the National Cancer Institute, Volume 95, Issue 22, 19 November 2003, Pages 1722–1725, https://doi.org/10.1093/jnci/djg090) We add letter “R” across **“Leptin Increase”**(if it is not there yet) parameter in **Column 9** for ”**…Alcohol consumption over 24 g”**
6. A study on alcohol biomarkers states: “**Alcohol is acutely as well as chronically toxic to the live**r, and hepatic enzymes such as **gamma-glutamyl transferase (GGT), alanine aminotransferase (ALT), and aspartate transaminase (AST) therefore leak into the blood** as part of the toxic response to high alcohol intakes.” (”Biomarkers of moderate alcohol intake and alcoholic beverages: a systematic literature review.”,Trius-Soler, M., Praticò, G., Gürdeniz, G. *et al.*, *Genes Nutr* 18, 7 (2023). https://doi.org/10.1186/s12263-023-00726-1). We are adding this physiological parameters to the most left column (if not added yet) and add letters “R” across them in **Column 9** for ”..alcohol intake”.
7. Regarding an obesity in women a study says: “**Adipose tissue is biologically active secreting adipokines, cytokines and estrogens.** In the obese, a variety of hormonal, metabolic and inflammatory changes occur that promote the pathogenesis of breast cancer.” (“Blood biomarkers reflect the effects of obesity and inflammation on the human breast transcriptome.”, Cho BA, Iyengar NM, Zhou XK, Morrow M., Carcinogenesis. 2021 Oct 26;42(10):1281-1292. doi: 10.1093/carcin/bgab066. PMID: 34314488; PMCID: PMC8546933.) We add these physiological parameters in the most left column of Table 1.1 and add “R” letters in **Column 11** for “BMI Higher 26.65 kg” factor.
8. A study on mammographic density (MD) informs: “When **stratifying by menopausal status, progesterone was positively associated with baseline MD among premenopausal** (+ 1.78 cm2 per doubling of hormone; P = 0.004) (Table 3), **but not postmenopausal** (− 0.07 cm2; P = 0.888), women (Table 4). **SHBG was positively associated with baseline MD in both premenopausal and postmenopausal women** (+ 6.58 cm2, P = 0.004; + 2.60 cm2, P = 0.042, respectively). Other hormones did not reach statistical significance in the stratified analyses” (*“Hormonal determinants of mammographic density and density change.”*, Gabrielson M, Azam S, Hardell E, et al, Breast Cancer Res. 2020 Aug 26;22(1):95. doi: 10.1186/s13058-020-01332-4). We add **“SHBG**” physiological parameter to the most left column and put “R” across it in **column 7** for a Breast Density,
9. Another study on Breast Density states: “Both **endogenous and exogenous estrogen may influence mammographic density**. Mammographic density decreases after menopause when ovarian function declines. Hormonal replacement therapy, with combination of estrogen and progesterone, increases mammographic density [2], while tamoxifen, which has antiestrogenic effect, decreases mammographic density [3]. **Mammographic density therefore can be regarded as a marker of the effect of estrogen on the breast tissue.”** (*“Does breast density show difference in patients with estrogen receptor-positive and estrogen receptor-negative breast cancer measured on MRI?”*, Chen JH, Hsu FT, Shih HN, et al, Ann Oncol. 2009 Aug;20(8):1447-9. doi: 10.1093/annonc/mdp362). This dependency between estrogen and breast density is observed when there is a change in estrogen but the change in breast density is not consistently shows the change in estrogen so we put **“Estradiol increases”**(if it is not there yet) parameter in left most column and add letter “Rc” (as conditional dependency) in **column 7** for Breast Density. We also place letter “R” in **column 4** for “Using Estrogen Replacement…” and **column 5** for “Postmenapause high estrogen” as the estrogen level growth is observed in both of these cases.
10. A study on Breast Density and biomarkers in premenopausal women found: “**Moderate positive correlations were observed among amino acids, acylcarnitines, and phosphatidylcholines** (with respective average correlations of 0.40, 0.31, and 0.37, not tabulated) and were stronger **among lysophosphatidylcholines and sphingomyelins** (with respective average correlations of 0.57 and 0.67, not tabulated)” (*”Biomarkers of mammographic density in premenopausal women.”,* His, M., Lajous, M., Gómez-Flores-Ramos, L. *et al, Breast Cancer Res* **23**, 75 (2021). https://doi.org/10.1186/s13058-021-01454-3). We add these physiological parameter changes in most left column of Table 1.1 and add letters “R” across them in **column 7** for “High Breast Density, BI-RADS code 4” to signify relationships between the physiological parameters and the disease causation factor.
11. According to research on anemia, anemia in cancers is related to TNF-α, GATA-1, etc: “**TNF-α inhibits hemoglobin production in a proportional fashion to the down-regulation of GATA-1** and also affects erythropoiesis induced by erythropoietin (Epo). TNF-α induces a decrease in the expression of FOG-1, a co-activator of GATA-1, as well as a proteasome-dependent decrease of GATA-1. In addition TNF-α suppresses the acetylated form of GATA-1, the post-translational modification required for DNA binding.” (*“Anemia in cancer”,* M. Dicato, L. Plawny, M. Diederich, Annals of Oncology Volume 21, Supplement 7, October 2010 https://doi.org/10.1093/annonc/mdq284). We add **“TNF-α increase”** physiological parameter(if not there yet) in the most left column of Table 1.1 and put letter “R” in **column 10** for “Anemia in Breast cancer” as TNF-α increase is one of the causes of anemia and also we add **“Down-regulation of GATA-1”** as physiological parameter in the most left column and add “R” for this parameter in same **column 7**. Both these parameters changes can be observed in cancer patients with anemia.
12. An obesity increases inflammation and a study states: “In normal-weight individuals, M2 macrophages present in adipose tissue promote anti-inflammatory signals. However, **obesity-related changes in adipocyte secretory profile (including, e.g., excess synthesis of interleukin 6 (IL-6) and tumor necrosis factor-alpha (TNFα)**) attract pro-inflammatory M1 macrophages to perpetuate pro-inflammatory cytokine signaling to trigger adipocyte cell death.” (*“Estrogens in Adipose Tissue Physiology and Obesity-RelatedDysfunction”*,KuryłowiczsA.,Biomedicines.2023,11(3):690.https://doi.org/10.3390/biomedicines1103 0690). As there is a dependency between being overweight and TNF-alfa increase and IL-6 increase we place these parameters in the most left column of Table 1.1 and add “R” in **column 2** for “Postmenopause Weight increase,BMI > 31.1” and **column 11** for “Premenapause women with BMI Higher 26.65 kg / m^2”
13. Another study on obesity and leptin states: “IPA analysis identified **leptin signaling as significantly upregulated in breast epithelial organoids from obese women compared with lean women**. Both obese CM and leptin treatment induced DNA damage in BRCA1+/-MCF10A cells while lean CM did not have this effect.” (*“Leptin Mediates Obesity-Induced DNA Damage in BRCA1 Breast Epithelial Cells”*, Priya Bhardwaj, Rohan Bareja, Sofya Oshchepkova, et al, Journal of the Endocrine Society, Volume 5, Issue Supplement_1, April-May 2021, Page A1024, https://doi.org/10.1210/jendso/bvab048.2095). We add leptin growth as parameter in most left column and “R” for this parameter in **column 2** for “Increased Weight” and **column 11** for **BMI Higher 26.65 kg.**
14. A study on impact of hormone replacement therapy and leptin has found: “A **positive linear correlation was found between leptin plasma levels and BMI only in obese patients** (r = 0.58; p < 0.01) both before and after estrogen treatment” (*“Leptin Levels in Menopause: Effect of Estrogen Replacement Therapy.”*, Rosa Maria Cento; Caterina Proto, et al,Hormone Research (2000) 52 (6): 269–273, https://doi.org/10.1159/000023493). We are adding **“Leptin level increase”** (if not added yet) to the most left column and letter “R” to **column 2** for “Postmenopause Weight increase, BMI > 31.1”
15. A study on carotenes informs on their impacts on decreased deoxyribonucleic acid **(DNA)**: “In studies in which **subjects were fed less than 25 mg/day of β-carotene**, either from foods or as a supplement, changes in the markers for antioxidant activity were minimal. Exceptions noted were **decreased deoxyribonucleic acid strand breaks observed when 22 mg/day of β-carotene was administered as carrot juice** (Pool-Zobel et al., 1997) and lowered copper-induced oxidation of low-density lipoprotein when 12 or 24 mg/day of β-carotene was given along with vitamins C and E” (*“Dietary Reference Intakes for Vitamin C, Vitamin E, Selenium, and Carotenoids.”,* Washington (DC): National Academies Press (US); 2000. 8, β-Carotene and Other Carotenoids. Available from: https://www.ncbi.nlm.nih.gov/books/NBK225469/). We place a **“Increase in DNA strands breaks”** as physiological parameter changed in the most left column of Table 1.1 and put letter “R” accordingly in the **column 8** for a “Low Total Carotenes” to signify a relationship between the physiological parameter changes and the disease causing factor.
16. An article on beta-carotene impact in healthy people informs: ”…**daily intake of β-carotene, a nontoxic antioxidant, reduces lipid peroxidation as assessed by serum lipid peroxide levels**.”(*“β-Carotene decreases markers of lipid peroxidation in healthy volunteers.”*, Gottlieb, K., Zarling, E. J., et al(1993). *Nutrition and Cancer*, *19*(2), 207–212. https://doi.org/10.1080/01635589309514251). We add the an **“Increase in lipid peroxidation”** as physiological parameter in the most left column of Table 1.1 and add letter “R”in **column 8** (“Low Total Carotenes”) to signify a relationship between the physiological parameters and a disease causation factor..
17. An article on beta-carotenoids informs: “**Provitamin A carotenoids are plant pigments that include beta-carotene**, alpha-carotene, and beta-cryptoxanthin [1]. The body converts provitamin A carotenoids into vitamin A in the intestine via the beta-carotene monooxygenase type 1 BCMO1 enzyme [1,3,6], although conversion rates may have genetic variability” (*“Vitamin A and Carotenoids.Fact Sheet for Health Professionals”*, NIH, https://ods.od.nih.gov/factsheets/VitaminA-HealthProfessional/). We place a **“Reduction of Provitamin A carotenoids”** as physiological parameter in most left column of Table 1.1 and add letter “R” in in **Column 8** (“Low Total Carotenes “) to signify a relationship.
18. An article on BRCA1 gene states: “**The BRCA1 protein is involved in repairing damaged DNA**. In the nucleus of many types of normal cells, the BRCA1 protein interacts with several other proteins to mend breaks in DNA. These breaks can be caused by natural and medical radiation or other environmental exposures, and they also occur when chromosomes exchange genetic material in preparation for cell division.” (*“BRCA1 gene”*, National Library of Medicine, Medline Plus, https://medlineplus.gov/genetics/gene/brca1). Another article on BRCA1 and BRCA2 mutations states: “**BRCA1 and BRCA2 play a crucial role in maintaining genome integrity by repairing double-strand DNA breaks *via* the homologous recombination repair** (HRR) pathway. Any **mutations that cause functional disruption of these proteins may prove to be highly deleteriou**s, leading to the development of cancer. In addition, BRCA1 and BRCA2 also play a critical role in cell division where they are transported to the cytosol to participate in regulating various molecular events during mitosis. Mutations impacting these important functions of BRCA1/2 can affect the delicate balance of the tightly regulated cellular processes that may lead to progression of disease.” (*“Role of BRCA Mutations in the Modulation of Response to Platinum Therapy.”*, Sanghamitra Mylavarapu1, Asmita Das,Frontiers in Oncology,Volume 8, 2018, https://doi.org/10.3389/fonc.2018.00016). We can place letter “R” across physiological parameter an **Increased DNA strand breaks** in a **Column 6 *(“BRCA1*, BRCA2, etc gene mutations (1cs)”)** as BRCA1 mutation may break this function of DNA Repair.

This way we fill out the rest of **Table 1.1** with physiological parameters and mark relationships between them and the found disease causation factors with letters **“R”**. We can use small letter near “R” to signify some details of these relationship, for example **“Rc”** where c - as conditional relationship. These relationships will need to be further analyzed and made more precise if necessary. **If it is required to increase a precision of conclusions about the physiological parameters causing a disease** the additional research on disease causation factors or physiological parameters can be used in a similar manner as above. In order to simplify placement of letters “R” the table’s columns can be enumerated and this enumeration may need to be added in few places as such a table may get pretty long.

Now we need to move to the next step of the method and for this let’s find out *intersections* (which we defined above) of the physiological parameters for the external factors which found to be causing Breast Cancer. We can see intersections in 4 rows in a Table 1.1. They are *rows* with multiple letters “R”which are marked in a gray color. Based on the intersections observed in Table 1.1 we include next physiological parameters in our matrix **Pm** below :

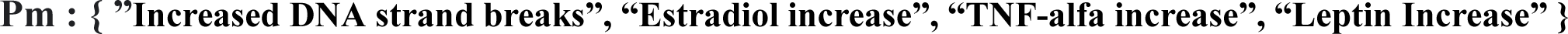

As we discussed in method’s explanation section, the first **Pm** matrix usually is not final and may require some corrections. We see our matrix **Pm** contains 4 parameters and it is the same as number of changes which are causing a Breast Cancer disease. So we don’t need to eliminate redundant physiological parameters but we need to notice the *intersection for some of these physiological parameters cannot be correct* because they would cause some factors to impact 2 or more physiological parameters while we have already determined above (using a disease causation criteria) that most of our disease causation factors for Breast Cancer must cause only 1 physiological change beyond 1-sigma and not more. We can see that a factor **“Premenapause women with BMI Higher 26.65 kg / m^2 (1cs)”** has crossings in **2** physiological parameters but can only cross 1 (as per disease causation criteria provided above). It currently impacts these physiological parameters as can be seen in Table 1.1: **TNF-Alfa increase, Estradiol Increase.** A physiological parameter **“TNF-alfa increase”** has 3 crossings (we don’t consider ”Postmenopause Weight increase,BMI > 31.1 (1cs)” and “Premenapause women with BMI Higher 26.65 kg / m^2 (1cs)” as different factors) and due to this the *probability that this intersection in TNF-alfa is random is extremely low.* **Estradiol increase** has 5 crossing and the probability of this crossings to be random is extremely low too. We need to see that causing this redundant intersection then? How to adjust our Pm matrix so all physiological parameter cross appropriately?

We notice that **“Premenapause women with BMI Higher 26.65 kg / m^2 (1cs)”** disease causation factor is applicable to *premenpause women* and for them the estrogen production in adipose tissue is not playing a large role as for *postmenopause women* as in premenopause women estradiol secretion happens mostly in ovaries.So we can safely assume that despite the fact what adipose tissue can cause Estradiol increase in premenopause women the role of such an increase likely less than 1-sigma for this disease causation factor (and we know also the factor can have only 1 impact). This leaves us with a change beyond 1-sigma in only 1 physiological parameter which is **TNF-Alfa increase**. It means that being “Premenapause women with BMI Higher 26.65 kg / m^2 (1cs)” is disease causation factor which impacts **TNF-Alfa increase** in premenopause women and this change is one of 4 physiological disease causes for a Breast Cancer.

Let’s notice how we used here **a property that the physiological parameter should be changed beyond 1-sigma to cause a disease** in order to remove a redundant crossing and correct matrix **Pm.** Now, we can also see an intersection in IL-6 in Table 1.1 but we don’t consider it is a valid intersection as it is for a related factors ”Postmenopause Weight increase,BMI > 31.1 (1cs)” and “Premenapause women with BMI Higher 26.65 kg / m^2 (1cs)” both related to obesity. So our new and final matrix **Pm** stays the same but the **Table 1.1** updated properly with a bold frame to incorporate right physiological changes as per our discussion above. The final matrix Pm is :

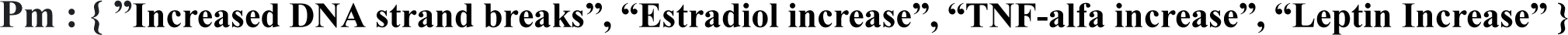

## Physiological Causes of Breast Cancer (Results of Analysis)

Based on the analysis of factors above the **set of 4 physiological changes beyond 1-sigma which are taking place at the same time are causing a Breast Cancer** disease. This set is as below.

1. Increased DNA strand breaks (beyond ∼ 1-sigma)
2. TNF-alfa increased beyond ∼ 1-sigma
3. Leptin level **increase** beyond ∼ 1-sigma
4. Estradiol increase (above ∼1-sigma)

This means a particular **women will get a breast cancer** after some time if she continuously has **Increased DNA strand breaks (beyond ∼ 1-sigma) AND TNF-alfa increased beyond ∼ 1-sigma AND Leptin level increase beyond ∼ 1-sigma AND Estradiol increase (above ∼1-sigma).** This process resembles a fuse which always blew off if the electrical current exceeds the fuse’s limit. The only random factors here are either you turn on air conditioning or electrical iron. As soon as you exceed the limit of the fuse it will always blew off and it does not matter if the cause was the air conditioning or iron. **It is not optional for the fuse to blew off.** The same concept applies as we can see for a breast cancer or other non-infectious disease pathology.

The **rows in Table 1.1 surrounded by bold** rectangles represent the physiological parameters found as result of analysis and which are causing Breast Cancer disease. Once they determined we can move these physiological parameters along with disease causing factors which are causing the change in them from **Table 1.1** to the **Table 1.2**. This table will contain the results of analysis: Disease Causation factors for Breast Cancer, physiological parameters which change beyond 1-sigma when these factors are present and relationship between Disease Causation factor and each physiological parameter.

## Validation of Method’s Results for Breast Cancer

To verify we completed all the steps of the method correctly so far let’s take a look how an existent research is consistent with our findings of Breast Cancer causes.

A research states: “Since **DNA double-strand breaks (DSBs) contribute to the genomic instability that drives cancer development**, DSB repair pathways serve as important mechanisms for tumor suppression. Thus, genetic lesions, such as BRCA1 and BRCA2 mutations, that disrupt DSB repair are often associated with cancer susceptibility.” (“DNA double-strand break repair pathway choice and cancer.”Aparicio T, Baer R, Gautier J., et al, DNA Repair (Amst). 2014 Jul;19:169-75. doi: 10.1016/j.dnarep.2014.03.014.). Which is consistent with our finding that Increased DNA strand breaks (beyond ∼ 1-sigma) are one of 4 causes of Breast Cancer. It does not cause Breast Cancer standalone but requires 3 other physiological changes.

Another article on TNF-Alfa informs: “Collectively, these **data reveal a pro-tumorigenic role of TNF-α during breast cancer progression and metastasis.** We systemize the knowledge regarding TNF-α-related therapies in breast cancer”(*“The dual role of tumor necrosis factor-alpha (TNF-α) in breast cancer: molecular insights and therapeutic approaches.”*, Cruceriu, D., Baldasici, O., Balacescu, O. et al., Cell Oncol. 43, 1–18 (2020). https://doi.org/10.1007/s13402-019-00489-1). This is consistent with a discovery of TNF-alfa increase as one of the causes of Breast Cancer.

A study on Leptin states: “ It controls adipose tissue growth and cell proliferation including breast tissue.<u>^77^</u> **Leptin levels were related to breast cancer aggressiveness** and can predict the type, grade, stage, lymph node involvement, hormone receptors, and recurrence in breast cancer.<u>^78^</u> **Leptin affects breast cancer biology in an endocrine, paracrine, and autocrine manner**”(“Linkage Between Obesity Leptin and Breast Cancer”. Atoum MF, Alzoughool F, Al-Hourani H., Breast Cancer (Auckl). 2020 Jan 10;14:1178223419898458. doi: 10.1177/1178223419898458). This is a consistent with the results of our method regarding Increased Leptin level as a cause of Breast Cancer.

**Table 1.1.**
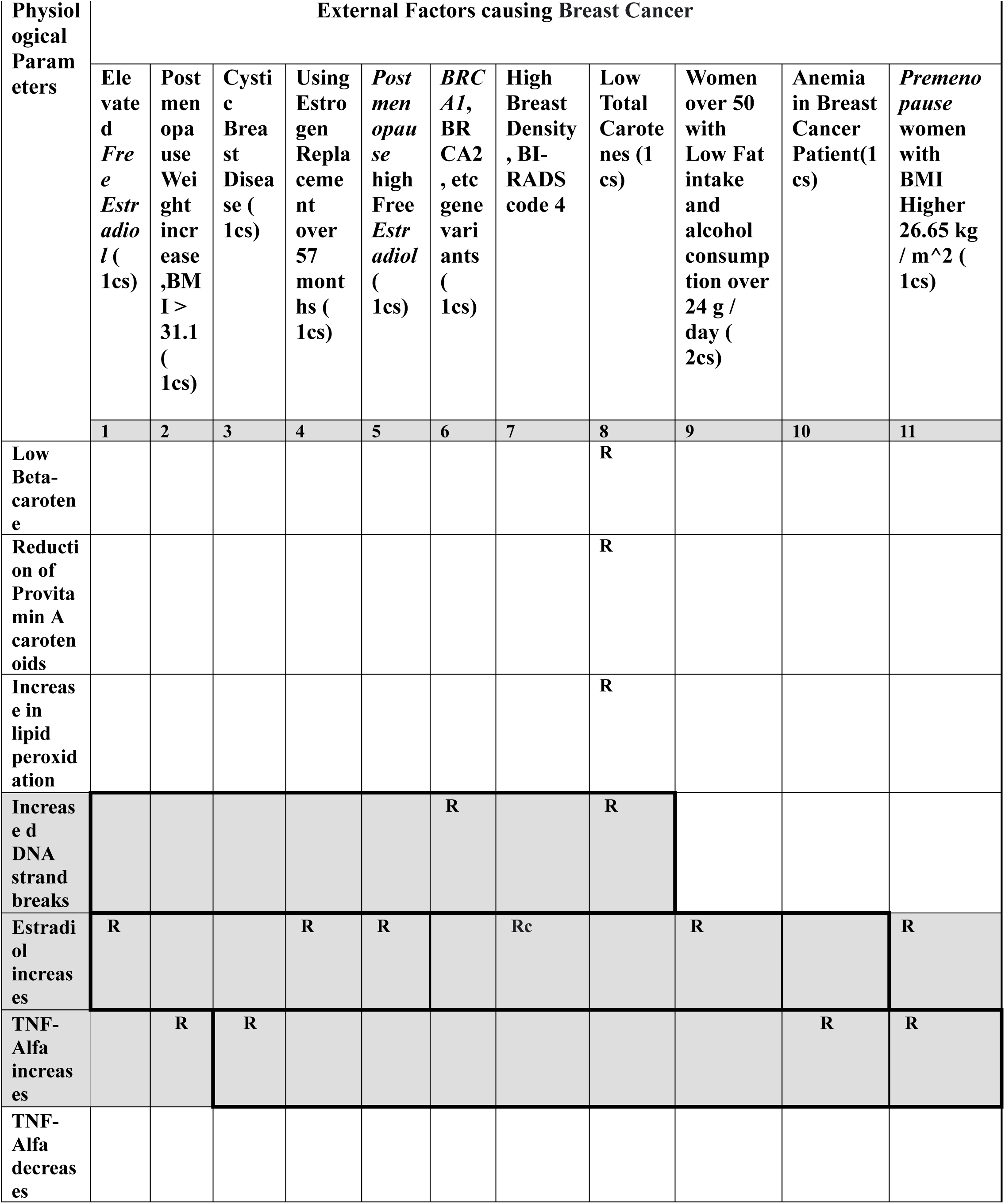
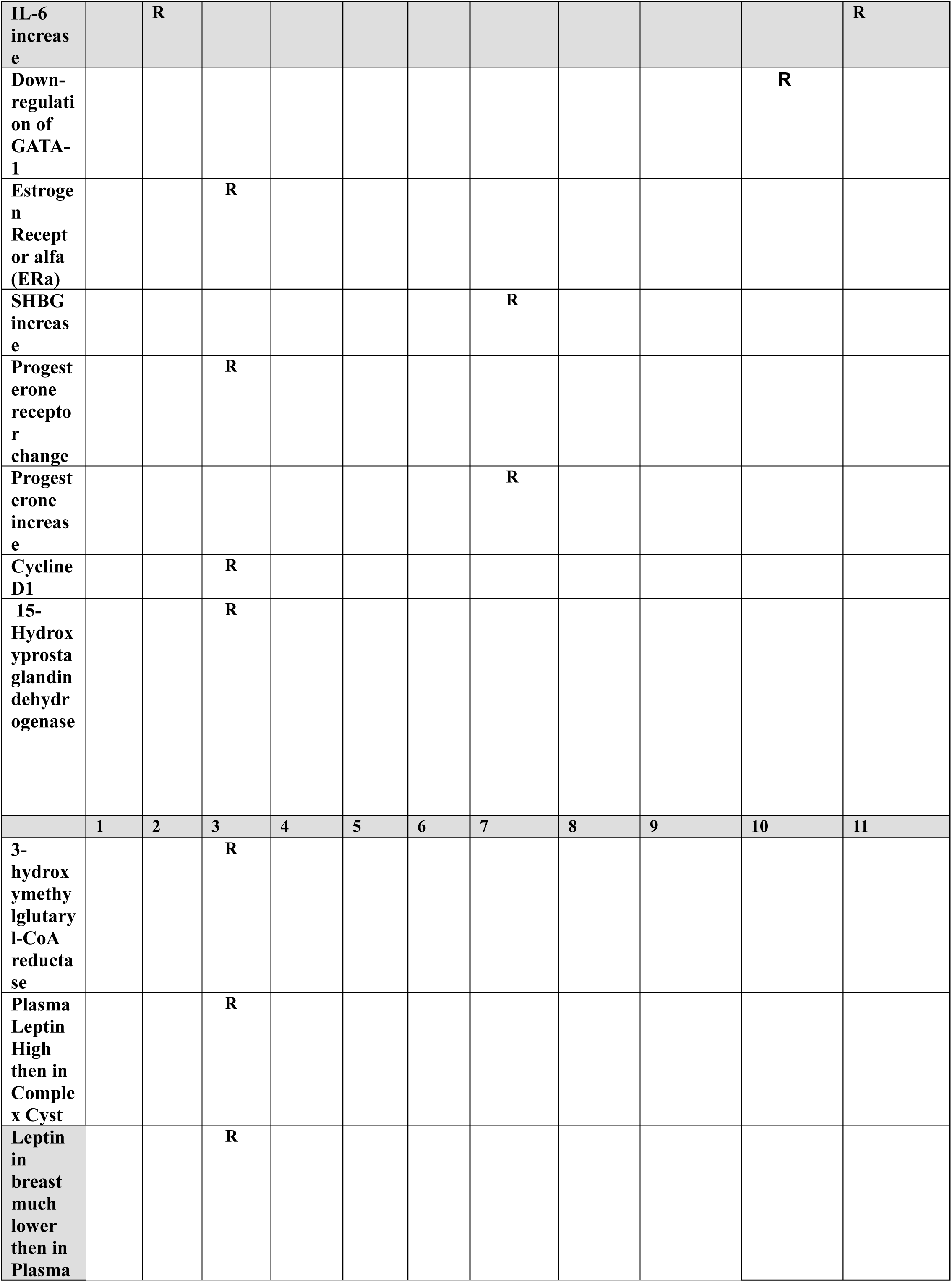
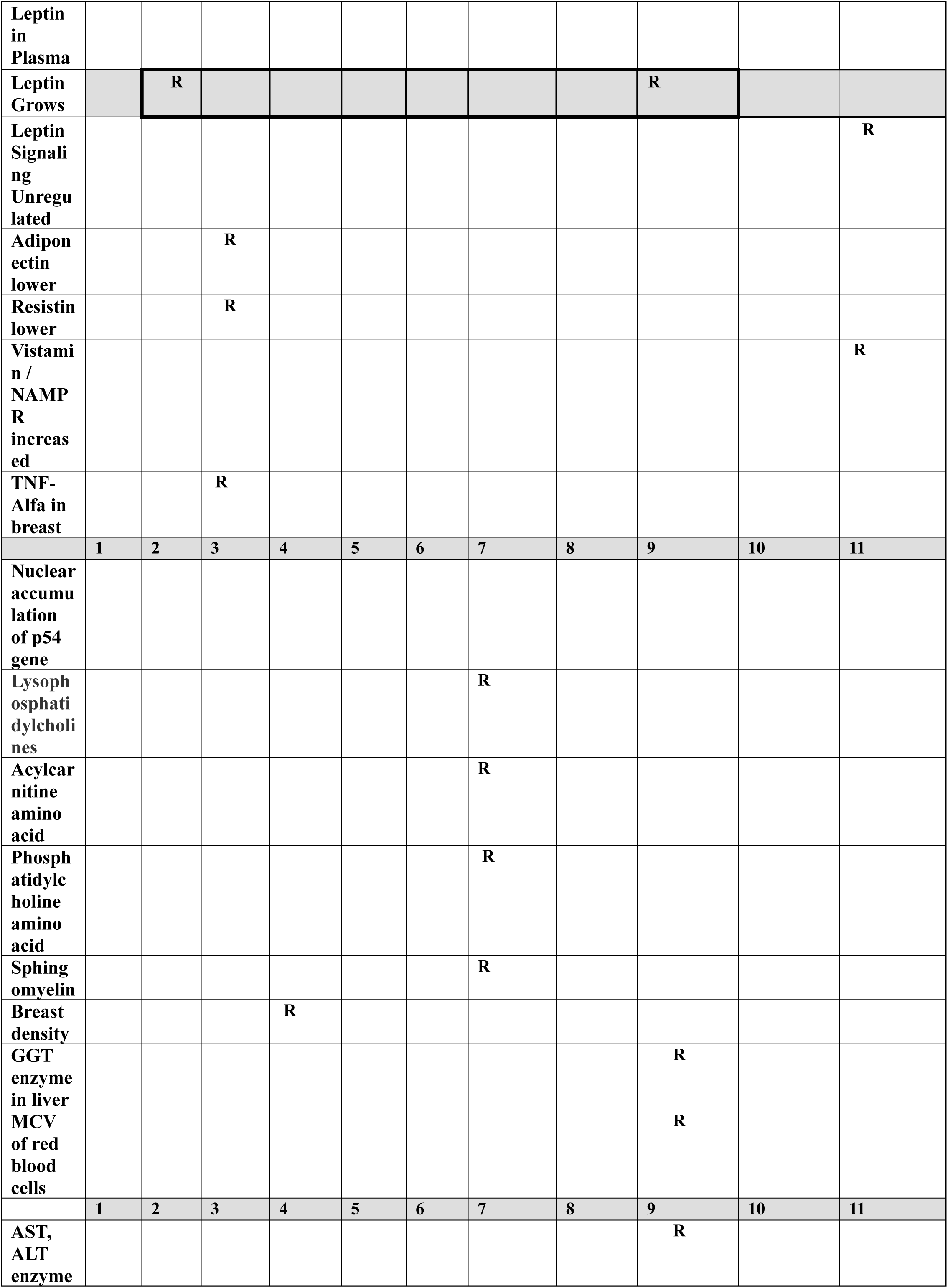
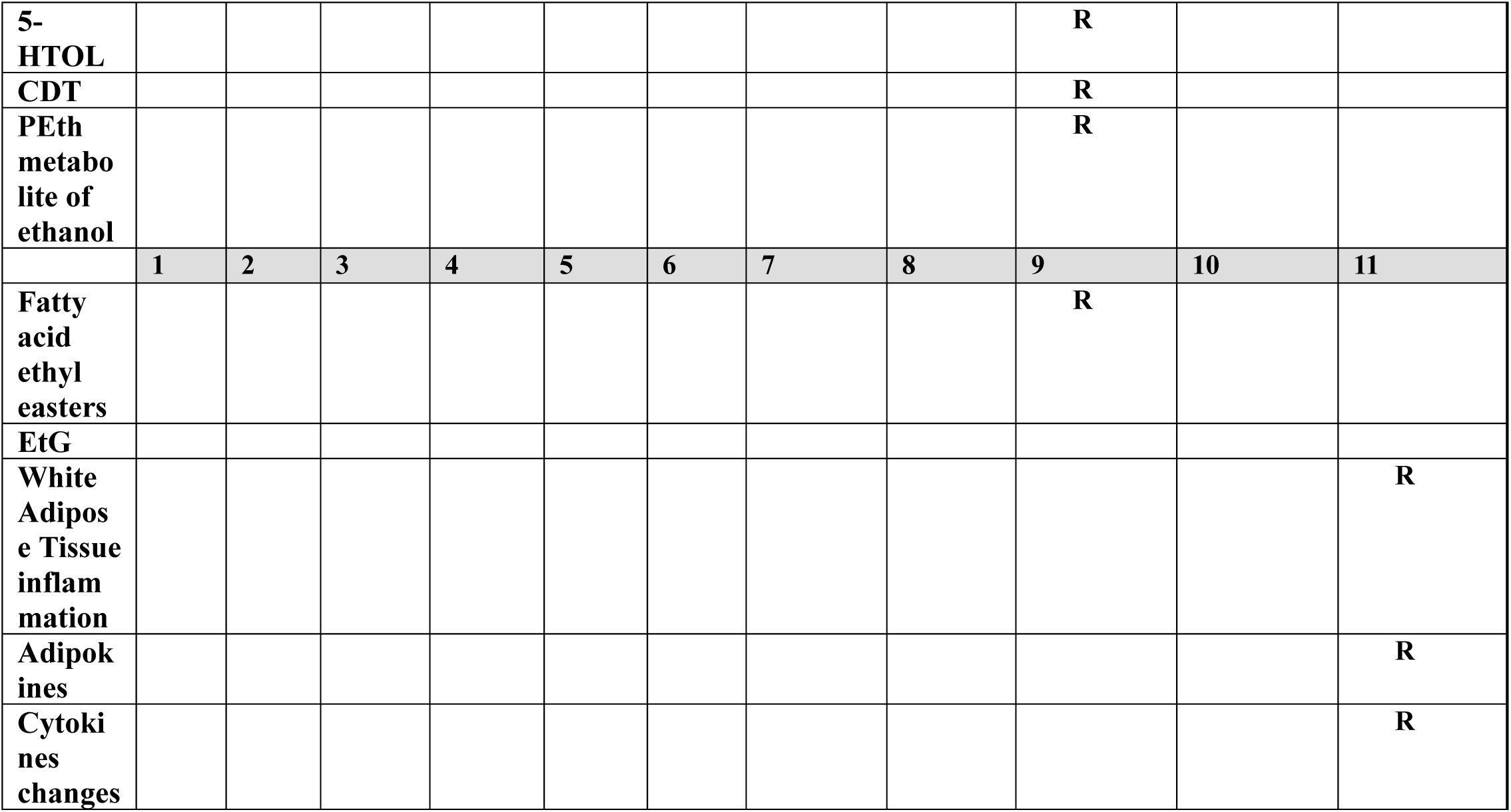
(**In the header**: factors determined to be Disease Causation factors for Breast Cancer using Disease Causation criteria. **In the most left column:** physiological parameter changes observed when the factors are present. “R” - signifies a relationship between such a physiological parameter change and a disease causing factor. Areas in gray are *intersections* of physiological parameters for few factors. *Bold frames* are final set of physiological parameters causing Breast Cancer)

An article on role of estrogen in cancers states: “The **oncogenic function of estrogens is considered in both classical and non-classical hormone-sensitive carcinomas such as prostate, breast**, endometrial, lung, colon, and ovarian cancers. The **molecular basis of cancer initiation by estrogen has been suggested through the production of aromatic estrogen metabolites** (catechol estrogens quinones) that are derived from normally formed catechol estrogens. Chemically, depurinating DNA-adducts are formed by the reaction of 4-OHE_1/2_ or 2-OHE_1/2_ with Adenine/Guanine bases which leads to DNA mutations ”(*“Estrogens and the risk of breast cancer: A narrative review of literature.”*, Al-Shami K, Awadi S, et al, Heliyon. 2023 Sep 17;9(9):e20224. doi: 10.1016/j.heliyon.2023.e20224.). This is also consistent with our finding that an increased estrogen (beyond 1-sigma) is one of 4 required cause of Breast Cancer. It won’t cause Breast Cancer as a standalone though.

We have found that 4 physiological causes of Breast Cancer determined by the presented method are consistent with the existing research which *was not used by the method* before. This way we confirmed the steps we have taken were done correctly and our method’s results are correct.

## A Room for an Error

So far we determined 4 physiological parameters changes beyond 1-sigma which *must c*ause a breast cancer if they are taking place a the same time long enough. From a practical perspective it give us a room for an error in determination of these parameters. The disease cannot start unless 4 of them changed so if we determined 1 parameter incorrectly but 3 others correctly we still should be able to prevent the disease by controlling these 3 physiological parameters to be within 1-sigma interval. We also should be able to use these 3 parameters to treat or possibly even cure a disease in early stages.

The **Table 1.3** shows external factors which will cause Breast cancer if combined together. A combination of 4 of each factors from each column will cause Breast cancer in an individual. If the factors represent the 2 causes then the number of factors can be less accordingly.

Using the **Table 1.3** we can conclude, for example, that **if a women has Low Total Carotenes AND has Cystic Breast disease and High Breast Density (BI-RADS code 4) AND she is Postmenopause with Weight increased, BMI > 31.1 (1cs) such a women will get a Breast Cancer** if she continuously has these factors.

This is not optional, the only option to remove one or more of these factors to prevent the disease. We notice here that we have 4 disease causation factors here where each impacting only 1 physiological parameter so it goes beyond 1-sigma. These 4 physiological changes beyond 1-sigma at the end are causing breast cancer and not the disease causing factors them self. Using a Table 1.3 we must combine disease causation factors in such a way that they impact al**l 4 physiological changes** which were determined for Breast Cancer disease.

**Table 1.2.** (**In the header:** 4 physiological parameters which change beyond 1-sigma if simultaneously present causes Breast Cancer. **In most left column:** some of Disease Causation factors for Breast Cancer which *only* can cause it if combined together so all 4 physiological parameters required to trigger Breast Cancer are changed. **1cs**, **2cs** (on the left, stand for a cause) - means the disease causation factor changes 1 or 2 physiological parameters beyond 1-sigma as determined by Disease Causation criteria. **x -** means that an Disease Causation factor is causing a change/related to the change in physiological parameter beyond 1-sigma so this parameter becomes one of 4 causes of Breast Cancer.)

| External Factors | Physiological parameters which changes are causing a Breast Cancer<br>(change should be beyond 1-sigma) |  |  |  |
| --- | --- | --- | --- | --- |
|  | Increased DNA strand breaks | TNF-alfa increase | Estradiol increase | Leptin Increase |
| Low Total Carotene ( 1cs) | **x** |  |  |  |
| *BRCA1*, *BRCA2*, etc gene mutations ( 1cs) | **x** |  |  |  |
| Premenapause women with BMI Higher 26.65 kg / m<sup>2</sup> ( 1cs) |  | **x** |  |  |
| Cystic Breast disease ( 1 cs) |  | **x** |  |  |
| Using estrogen replacement over 57 months (1 cs) |  |  | **x** |  |
| Postmenapause high free estradiol (1cs) |  |  | **x** |  |
| High Breast Density, (BI-RADS code 4) |  |  | **x** |  |
| Women over 50 with low fat intake but **Alcohol** consumption over 24g / day ( 2 cs) |  |  | **x** | **x** |
| Anemia after cancer ( 1 cs) |  | **x** |  |  |
| Postmenopause Weight increase,BMI > 31.1 ( 1cs) |  |  |  | **x** |

The results derived from **Table 1.3** can be validated empirically even on the small set of patients. We would need to observe their medical history and see if they had these factors present together for some time before they became sick with a breast cancer. Different combinations from the table above can be observed during such experiments.

## Building Hypothesis of Breast Cancer Pathology

Now when we determined 4 physiological changes beyond 1-sigma which are causing Breast Cancer we can demonstrate how the presented method simplifies building a hypothesis of disease pathology by connecting these found physiological changes together. We have determined using a method that Breast Cancer is caused by these 4 physiological changes **: ”Increased DNA strand breaks”** beyond 1-sigma**, “Estradiol increase”** beyond 1-sigma**, “TNF-alfa increase”** beyond 1-sigma**, “Leptin Increase”** beyond 1-sigma.

**Table 1.3.**
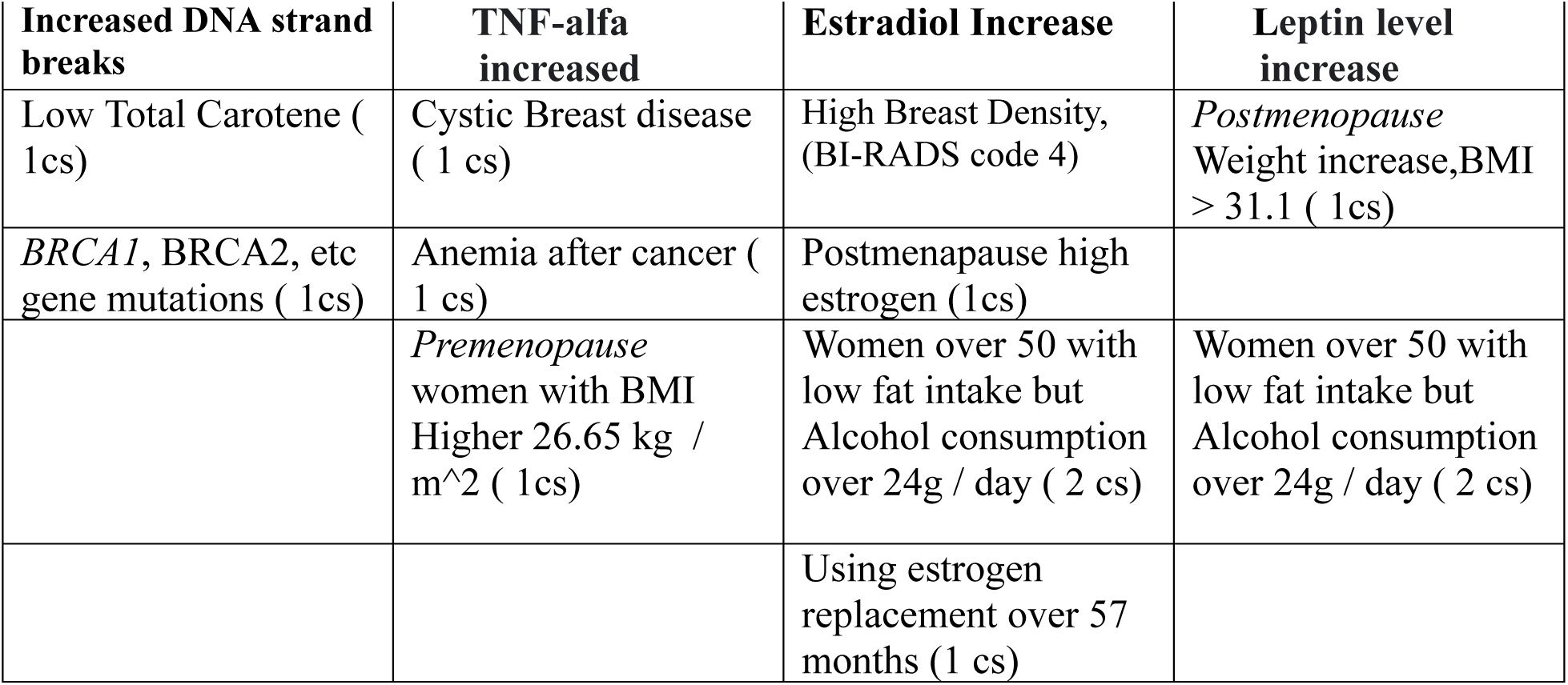
(In the header: 4 physiological parameters changes (beyond approx. 1-sigma) which should be simultaneously present to cause Breast Cancer. In the columns: Disease Causation factors which are either causing the physiological change beyond 1-sigma (or physiological change present if the factor exists). Any set of few Disease Causing factors (one from each column) from each column will cause a Breast Cancer. The factors *cannot* cause Breast Cancer as standalone)

| Increased DNA strand breaks | TNF-alfa increased | Estradiol Increase | Leptin level increase |
| --- | --- | --- | --- |
| Low Total Carotene ( 1cs) | Cystic Breast disease ( 1 cs) | High Breast Density, (BI-RADS code 4) | *Postmenopause* Weight increase,BMI > 31.1 ( 1cs) |
| *BRCA1*, *BRCA2*, etc gene mutations ( 1cs) | Anemia after cancer ( 1 cs) | Postmenopause high estrogen (1cs) |  |
|  | *Premenopause* women with BMI Higher 26.65 kg / m<sup>2</sup> ( 1cs) | Women over 50 with low fat intake but Alcohol consumption over 24g / day ( 2 cs) | Women over 50 with low fat intake but Alcohol consumption over 24g / day ( 2 cs) |
|  |  | Using estrogen replacement over 57 months (1 cs) |  |

**NOTE:** the list of factors in **Table 1.3** is obviously not complete and provided as per author's knowledge of specific research, other factors can be added in the future if new research finds them, etc. At the same time the future factors as current ones will be related to the physiological parameters in the columns of the table as these parameters the real cause of a disease (the factors are only causing their change).

Using an existing research about these physiological parameters change we can build a relationship between them which will create hypothesis of Breast Cancer pathology as below. For additional details on this process please refer to building of Autism pathology hypothesis section.

**Breast Cancer pathology (hypothesis):** *An increased beyond 1-sigma a number of DNA strand breaks (caused by such factors as low total carotene, etc) cannot be repaired due increased beyond 1-sigma TNF-alfa level (caused by Cystic Breast disease, for example) and an increased beyond 1-sigma level of leptin (caused by postmenopause women weight increase, BMI > 31.1, for example) where a leptin level increase reduces energy capabilities of breast cell in particular for repair processes and along with additional level of DNA mutations due to adducts created by high estradiol level beyond 1-sigma deprive the breast cell from the DNA repair capabilities required to compensate for a normal DNA degradation processes and DNA damages and mutation accumulate beyond control. This results in triggering of Breast Cancer*. In short, the disruption of DNA damage / repair homeostasis leads to a breast cancer.

## 2. AUTISM: DETERMINATION OF CAUSES

Autism spectrum disorder (ASD) is a neurological and developmental disorder that affects how people interact with others, communicate, learn, and behave. Although autism can be diagnosed at any age, it is described as a “developmental disorder” because symptoms generally appear in the first 2 years of life.

According to National Institute of Mental Health occurrence of autism in boys as 36.5 per 1000 and in girls 8.8 per 1000 as per data from year 2018. Using a formula (1.0) presented in the beginning of the article we calculate the number of physiological changes (number of disease causes) which are required to trigger Autism in boys and girls:

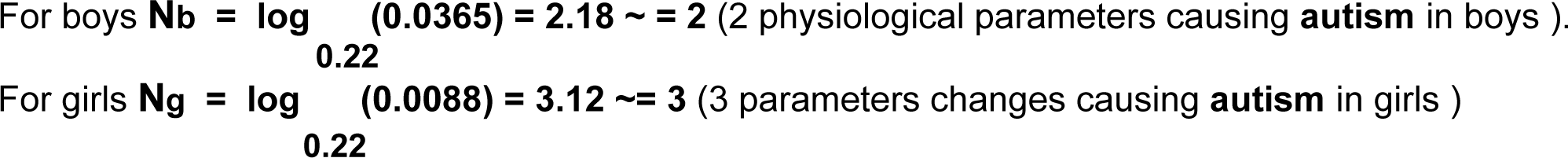

As per calculations above for boys we get Nb = ***2*** *physiological parameters changes which are causing autism in boys* and for girls we get Ng = ***3*** *physiological parameters changes which are causing autism in girls*.

Let’s list the factors which are causing a Autism and explain how they are determined based on Disease Causation criteria provided above and developed in article ***“*A Connection between Factors Causing Diseases and Diseases Frequencies: Its Application in Finding Disease Causes”** *(Alan Olan,* Journal of Clinical Trials, Vol.13, Issue 4.). We are going to populate these factors in a header of **Table 2.1**:

1. A study of Autism (ASD) risk in children analyzed 2,734 children and states: “When examined in combination, only **mothers with obesity and PGDM (hazard ratio 3.91, 95%** confidence interval 1.76– 8.68) and **those with obesity and gestational diabetes (hazard ratio 3.04**, 95% confidence interval 1.21– 7.63) **had a significantly increased risk of offspring AS*D****.” (“The Association of Maternal Obesity and Diabetes With Autism and Other Developmental Disabilities.”*, Li M, Fallin MD, Riley A, Landa R, et al, Pediatrics. 2016 Feb;137(2):e20152206. doi: 10.1542/peds.2015-2206). It is known that HR can be treated as an approximation of RR. So we can write that **RR =∼ HR = 3.91** and **RR=∼HR**=**3.04** which are both within a range of **4.5+/-50%(2.27-6.81)** of **Disease Causation criteria** for 1 physiological parameter change (see article [1]). It means that being a **mother with obesity and PGDM** as well as being **a mother with obesity and gestational diabetes is a causation factor for Autism**(ASD) *which makes 1 physiological parameter change go beyond 1-sigma interva*l (actually, the interval is slightly more narrow). This physiological parameter’s change is one of 2 (for boys) or 3 (for girls) total physiological parameters changes beyond 1-sigma which need to be ***simultaneously*** occurring in a child to cause Autism. We determined these numbers of physiological parameters required to be changed approx. beyond 1-sigma for boys as 2 and 3 for girls above using a formula for number of disease causes (1.0). So it means that being a **mother with obesity and PGDM is not causing Autism a standalone factor**. It *must cause Autism in* boys though if another physiological parameter (which we will find later) is going beyond 1-sigma. The same factor **must** cause Autism in girls if other 2 physiological parameter will go beyond 1-sigma interval. We add this factor in header of Table 2.1 as **Column 9** “Mother with Diabetis + Obesity”.
2. Study of impact of copy number variation (CNV) and ozone exposure states:“**children with high duplication burden** who were **also exposed during the prenatal period to high ozone levels** had nearly **a three-fold greater risk to** develop autism (**OR=2.8** (1.2–6.9)). Even stronger associations were seen for children wit**h high ozone exposures during the first and second years of life**, with **OR=4.2** (1.5–11.7) and **OR=3.4** (1.4–8.6) ” (*“The joint effect of air pollution exposure and copy number variation on risk for autism*.”, Kim D, Volk H, Girirajan S, Pendergrass S, Autism Res. 2017 Sep;10(9):1470-1480. doi: 10.1002/aur.1799.). We see that both **OR=2.8** and **OR=3.4** are within **4.5+/-50%(2.27-6.81)** of **Disease Causation criteria** for 1 physiological parameter change. It means **high CNV and High Ozone exposure during 1-2nd years of life or during prenatal period is a cause of Autism** and it forces 1 physiological parameter to change beyond 1-sigma interval. High CNV and Ozone exposure will not cause Autism as standalone factor but with 1 other factor for boys or 2 other factors for girls it must cause Autism after some time. The causation of disease is not optional as per Disease Causation criteria (see article [1]). We place this factor in Table 2.1 header as **Column 3** and call it **“High CNV + Ozone Exposure (1cs)”.**
3. A study on impact of fever during during pregnancy on development of Autism states: “Risk increased markedly with exposure to **three or more fever episodes after 12 weeks’ gestation (aOR**, 3.12; 1.28–7.63). ASD risk appears to increase with maternal fever, particularly in the second trimester.” (*“Prenatal fever and autism risk.”*, Hornig, M., Bresnahan, M., Che, X. et al. Mol Psychiatry 23, 759–766 (2018). https://doi.org/10.1038/mp.2017.119). We see **OR = 3.12** and it is within **4.5+/-50%(2.27-6.81)** of **Disease Causation criteria** for 1 physiological parameter change. It means **three or more fever episodes after 12 weeks’ gestation is a causation factor for autism** which is changing 1 physiological parameter out of 2 required for boys and out of 3 required for girls to trigger Autism. Such a fever will not cause Autism as a standalone factor but if other physiological parameters 1 (for boys) or 2 (for girls) also were changed beyond 1-sigma it *must* trigger Autism. These other 1 or 2 physiological parameters of course need to be specific parameters which are causing Autism (which will be determined later in this article). We add this causation factor now to Table 2.1 in **Column 4** as “**Fever after 12 week of pregnancy”.**
4. Research on valproate exposure during pregnancy and its impact on risk of Autism development states: “ The **aHRs for ASD and ID after topiramate exposure were 2.8** (95% CI, 1.4-5.7) and 3.5 (95% CI, 1.4-8.6), respectively, and **after valproate exposure were 2.4** (95% CI, 1.7-3.3) and 2.5 (95% CI, 1.7-3.7).” (*“Association of Prenatal Exposure to Antiseizure Medication With Risk of Autism and Intellectual Disability.”*, Bjørk M, Zoega H, Leinonen MK, et al, JAMA Neurol. 2022;79(7):672–681. doi:10.1001/jamaneurol.2022.1269). The **RR=∼aHR = 2.8** and **RR=∼aHR = 2.4 are within a range of 4.5+/-50%(2.27-6.81)** of **Disease Causation criteria** for 1 physiological parameter change. It means **topiramate exposure or valproate exposure during pregnancy is a causation factor for Autism** which changing 1 physiological parameter (which yet to be found) to go beyond 1 sigma out of 2 or 3 (for boys and girls accordingly) required to cause a Autism. Topiramate exposure or valproate exposure during pregnancy does not cause Autism as standalone factor but only if the change to the impacted physiological parameter will coexist with 1 or 2 other required physiological parameters changes beyond 1-sigma (as per Disease Causation criteria in article [1]). We add valproate exposure as disease causation factor to Table 2.1 in **Column 1** as “Valproate (1cs)” where 1cs stands for 1 cause and means the causation factor is changing 1 physiological parameter beyond 1-sigma interval (actually, slightly less)
5. A research on risk of Autism in 2^nd^ pregnancies shows that the risk depends on Inter pregnancy interval (IPI): “**An inverse association between IPI and odds of autism among 662 730 second-born children was observed**. In **particular, IPIs of <12**, 12 to 23, and 24 to 35 months **were associated with odds ratios** (95% confidence intervals) **for autism of 3.39** (3.00–3.82), 1.86 (1.65–2.10), and 1.26 (1.10–1.45) relative to IPIs of ≥36 months.” (*“Closely spaced pregnancies are associated with increased odds of autism in California sibling births.”*, Cheslack-Postava K, Liu K, Bearman PS. Pediatrics. 2011 Feb;127(2):246-53. doi: 10.1542/peds.2010-2371. Epub 2011 Jan 10. PMID: 21220394; PMCID: PMC3387860.). We see that only **OR=3.39** for ITI less than 12 months is within a range **4.5+/-50%(2.27-6.81)** of **Disease Causation criteria** for 1 physiological parameter change. It means **an Interpregnancy interval less than 12 moths is a disease causation factor for Autism** which impact 1-physiological parameter to change beyond 1-sigma interval. This change will not cause Autism as standalone but only together with 1 (for boys) or 2 (for girls) other additional specific disease causation physiological parameters changes beyond 1-sigma will cause Autism. We are adding this disease causation factor to Table 2.1 in **Column 7** as **“Child born within 1st year of previous pregnancy (1cs)”**
6. A study found the risk of Autism in case of maternal depression: “the associations between antidepressant use and autism spectrum disorder. **Any antidepressant use during pregnancy in mothers** of cases was 1.3% compared with 0.6% of controls **equating to an almost twofold increase in risk of autism spectrum disorder (1.90**, 1.15 to 3.14). **These associations too were largely observed due to associations with autism without intellectual disability (2.54**, 1.37 to 4.68) with no increased odds for autism with intellectual disability.” (*“Parental depression, maternal antidepressant use during pregnancy, and risk of autism spectrum disorders: population based case-control study.”*, Rai D, Lee B K, Dalman C, Golding J et al, BMJ 2013; 346 :f2059 doi:10.1136/bmj.f2059). We have **OR = 2.54** which is within a range **4.5+/-50%(2.27-6.81)** of **Disease Causation criteria** for 1 physiological parameter change. We see that **maternal depression during pregnancy and antidepressant usage is a disease causation factor for Autism** impacting 1 physiological parameter to go beyond 1-sigma interval. It will not cause Autism as standalone factor but together with 1 (for boys) or 2 (for girls) other physiological changes beyond 1-sigma must cause Autism in a child. We can add this to Table 2.1 **Column 2** as “**Depression in mother”.**
7. A study on reoccurrence of Child Autism(CA) in relatives measured a relative recurrence risk (RRR)as a hazard ratio (HR) and informs: “The **adjusted sibling RRR for CA was 17.4** (16.3–18.7) with site-specific estimates ranging from 9.2 (6.7–12.5) in Denmark to 24.8 (13.7–45.0) in Finland. The **adjusted RRR for full-siblings was significantly larger than for half-siblings; 18.5** (17.2–19.8) vs. 11.4 (8.9–14.6).” (*“Recurrence Risk of Autism in Siblings and Cousins: A Multinational, Population-Based Study.”*, Hansen SN, Schendel DE, Francis RW, et al, J Am Acad Child Adolesc Psychiatry. 2019 Sep;58(9):866-875. doi: 10.1016/j.jaac.2018.11.017.). We see that for full-siblings the **RR=∼HR=18.5** is within a range **20.6**+/-50% (10.3-30.9) **of Disease Causation criteria for 2 physiological parameters** changes. It means **being a full-sibling of the older Autistic Child is a causation factor for Autism** which changes 2 physiological parameters beyond 1-sigma. We are adding this factors to Table 2.1, Column 8 as “**Having one child with Autism (2cs)”**, where 2 cs means a factor impacting 2 causes as 2 physiological parameters beyond 1-sigma.
8. A study on impact of father’s age and risk of Autism states: “**Compared with offspring born to fathers 20 to 24 years old, offspring of fathers 45 years and olde**r **were at heightened risk of autism (hazard ratio [HR] = 3.45**; 95% CI, 1.62-7.33), attention-deficit/hyperactivity disorder (HR = 13.13; 95% CI, 6.85-25.16), psychosis (HR = 2.07; 95% CI, 1.35-3.20), bipolar disorder (HR = 24.70; 95% CI, 12.12-50.31) ” (*“Paternal Age at Childbearing and Offspring Psychiatric and Academic Morbidity.”*,D’Onofrio BM, Rickert ME, Frans E, et al. JAMA Psychiatry. 2014;71(4):432–438. doi:10.1001/jamapsychiatry.2013.4525). We see that **RR=∼HR=3.45** is within a range **4.5+/-50%(2.27-6.81)** of **Disease Causation criteria** for 1 physiological parameter change. It means that **paternal age over 45 years old is a causation factor for Autism** which changing 1 physiological parameter beyond 1-sigma. This factor will not cause Autism in a offspring as a standalone but with combination of 1 (for boys) or 2 (for girls) others must cause Autism after some time if the factors are not removed quickly. We add this causation factor to Table 2.1, Column 5 as “**Father older 45 year (1cs)”.**
9. A study of traffic-related air pollution (TRP) and its risk of Autism informs on Autism cases: “Cases were **more likely to live at residences in the highest quartile TRP exposure** during pregnancy (OR=1.98, 95%CI 1.20–3.31) and **the first year of life (OR=3.10**, 1.76–5.57) compared to controls.” (“Traffic-related air pollution, particulate matter, and autism.”,Volk HE, Lurmann F, Penfold B, et al, JAMA Psychiatry. 2013 Jan;70(1):71-7. doi: 10.1001/jamapsychiatry.2013.266.). We see that **OR=3.10** is within a range **4.5+/-50%(2.27-6.81)** of **Disease Causation criteria** for **1** physiological parameter change. Other ORs values are below and not enough to be causation factors. It means **a residence in the highest quartile TRP exposure the first year of life is a causation factor** which changes 1 unknown physiological parameter beyond 1-sigma. We put this factor in Table 2.1, Column 11 as **“Air Pollution in 1^st^ year(1cs)”.**
10. A study on iron intake during pregnancy and risk of Autism states: “A significant multiplicative interaction (*P* = 0.002) was found between **low maternal iron intake and older maternal age at delivery, with more than a 5-fold increased risk of ASD for mothers aged 35 years or older with iron intake in the lowest quintile** as compared with younger mothers with iron intake in the highest quintile (**OR = 5.01**, 95% CI: 1.98, 12.69) ” (*“Maternal intake of supplemental iron and risk of autism spectrum disorder.”*, Schmidt RJ, Tancredi DJ, Krakowiak P, et al, Am J Epidemiol. 2014 Nov 1;180(9):890-900. doi: 10.1093/aje/kwu208. Epub 2014 Sep 22.). We see **OR=5.01** is within a range **4.5+/-50%(2.27-6.81)** of **Disease Causation criteria** for **1** physiological parameter change. It means **a pregnancy of women over 35 years old with iron intake in the lowest quintile is a disease causation factor for Autism** which impact 1-physiological parameter to change beyond 1-sigma. This physiological parameter is one of multiple required to be present to cause Autism. The discovered factor make the change to this parameter and this way is helping to trigger Autism. We adding the discovered factor in Table 2.1, **Column 10** as **“Low Iron Intake in Pregnancy over 35(1cs)”**

We need to notice that multiple disease causation factors which we have found for Autism should impact **the same set of 3 physiological parameters** (2 for boys and 3 for girls). It is those 3 parameters’ changes beyond 1-sigma which are causing Autism in children as per research in article [1]. The disease causation factors are just negatively impacting these disease causing physiological parameters’ changes beyond 1-sigma (slightly less, actually) so the disease is triggered. There can be **many different disease causing factors like this but only 3 physiological parameters** which any of these factors impact so they change beyond 1-sigma interval and cause the disease. So *if we have 40 different factors, for example, and only 3 unique and specific physiological parameters they change beyond 1*-sigma interval its seems within a *common sense that there should be a way to determine those physiological parameters* and thus find the real disease causes. We will continue to discover below one way how to do it in practice.

Now we need to find physiological parameter changes related to the disease causation factors we found for Autism. We list them here and group them by related disease causation factors which we have determined above. The physiological parameters we find here will be populated into **Table 2.1** (the most left column):

### Valproate as a factor

1. A study inform that Sodium valproate has next mechanism of actions: “**GABA potentiation, blocks voltage gated sodium channels, epigenetically inhibits histone deacetylase**”. Accoding to the same article valproate has next side effects:**”GI upset, weight gain,..”**. The reasons of negative action of valproate the study suggest can be attributed to : ”.**..the inhibition of histone deactylase with associated changes in gene expression**, **increases in foetal oxidative stress**, or the **antagonism of folate required for DNA synthesis**” (*“Sodium valproate in pregnancy: what are the risks and should we use a shared decision-making approach?”*, Macfarlane A, Greenhalgh T., BMC Pregnancy Childbirth. 2018 Jun 1;18(1):200. doi: 10.1186/s12884-018-1842-x.). We place this physiological parameters in left most column of Table 2.1 and mark **Column 1** (“Valproate”) with letters **“R”** acrooss matching physilogical changes.
2. Another study on Valproate states: “Significant weight gain is one of the most common problems with VPA in both children and adults. In addition, the **chronic use of valproates is associated with an increase in the level of fast plasma insulin (FPI)**, the **development of insulin resistance**, especially in a subgroup of children and women. In addition **to weight gain and insulin resistance**, VPA-MetS is accompanied by **dyslipidemia**, arterial hypertension, and **type 2 diabetes mellitus**.” (*“Valproate-Induced Metabolic Syndrome”,* by Natalia A. Shnayder,Violetta,Vera V. Trefilova, et al,Biomedicines 2023, 11(5), 1499; https://doi.org/10.3390/biomedicines11051499). We add the found related physiological parameters to the most left column of Table 2.1 and add letters “R” across matching parameters in Column 1 (“Valproate”).

### Depresssion as a factor

1. A study on Depression’s biomarkers states: **“IL-6** (*P*<0.001 in all meta-analyses; 31 studies included) **and CRP** (*P*<0.001; 20 studies) **appear frequently and reliably elevated in depression**.<u>^40^</u> Elevated tumor necrosis factor alpha **(TNFα) was identified in early studies** (*P*<0.001)”, also it informs about such biomarkers as “ **hypothalamic–pituitary–adrenal (HPA) axis hyperactivation**, thyroid dysfunction, **reduced dopamine**, noradrenaline or 5-hydroxyindoleacetic acid, **increased glutamate**, increased superoxide dismutase and lipid peroxidation, attenuated cyclic adenosine 3′,5′-monophosphate and mitogen-activated protein kinase pathway activity, **increased proinflammatory cytokines**, **alterations to tryptophan, kynurenine, insulin** and specific genetic polymorphisms.”. Same study also informs on metabolic profile changes in depression: “The main biomarkers associated with metabolic illness include **leptin, adiponectin, ghrelin, triglycerides**, **high-density lipoprotein (HDL), glucose, insulin** and **albumin**.<u>^87^</u> The associations between many of these and depression have been reviewed: **leptin** and **ghrelin** appear lower in depression than controls ” and additionally the study states: “Some **neurotrophic factors are reduced in depression compared to controls (BDNF, NGF, GDNF**)” and “**Insulin resistance may be increased in depression**, albeit by small amounts. **Lipid profiles, including HDL-cholesterol, appear altered** in many patients with depression”. The study informs about Neurotransmitter changes in depression: “5-HT and receptors, NA, DA, glutamate/glutamine, **GABA, histamine**, MHPG, HVA” (*“Biomarkers for depression: recent insights, current challenges and future prospects.”*, Strawbridge R, Young AH, Cleare AJ., et al, Neuropsychiatr Dis Treat. 2017 May 10;13:1245-1262. doi: 10.2147/NDT.S114542. PMID: 28546750; PMCID: PMC5436791.). We add these related to Depression physilogical parameters changes to the most left column and mark the relationships with Depression with letters “R” in **Column 2** (“Depression”) accordingly. This letters “R” mean that this parameters’ changes are observed during the presence of Depression which is a disease causation factor for Autism.

### Ozone exposure as a factor

1. A study on ozone exposure biomarkers reports: “In this longitudinal panel study, we found that short-term **exposure to low concentrations of ozone** may lead to **increased biomarkers of inflammation and oxidative stress**, including **TNF-α, sICAM-1, and MDA**. We also observed **reduced methylation of the mitochondria D-loop region** with ozone exposure.”, it also states: “extensive evidence suggested that **short-term ozone exposure was associated with respiratory inflammation.**“ (*“Acute Effects of Personal Ozone Exposure on Biomarkers of Inflammation, Oxidative Stress, and Mitochondrial Oxidative Damage - Shanghai Municipality, China, May-October 2016.”*, Xia Y, Niu Y, Cai J, et al, China CDC Wkly. 2021 Nov 5;3(45):954-958. doi: 10.46234/ccdcw2021.232. China CDC Wkly. 2021 Nov 5;3(45):954-958. doi: 10.46234/ccdcw2021.232.). We add these related to ozone exposure physiological changes to the most left column (if not yet added) of Table 2.1 and mark them with letters “R” the **Column 3** (“Ozone exposure (1cs)”) across these parameters.

### Fever in pregnancy as a factor

1. A study on fever during pregnancy informs: “A total of 100 pregnant women **who consulted for fever** were included. The etiologies were **common viral infections** (37 %), **influenza** (21 %), **pyelonephritis** (11 %), viral gastroenteritis (6%), chorioamnionitis (5%), other (5%). The etiology was unknown for 15 %”, and also “Of the 32 patients with confirmed fever who had no etiologic diagnosis at the initial work-up in the emergency room, 19/32, 59 % received **presumptive treatment with amoxicillin against *Listeria monocytogenes***.”(*“Causes and consequences of fever during pregnancy: A retrospective study in a gynaecological emergency department.”*, Egloff C, Sibiude J, Couffignal C, et al, J Gynecol Obstet Hum Reprod. 2020 Nov;49(9):101899. doi: 10.1016/j.jogoh.2020.101899. Epub 2020 Aug 24.). We are adding these found physiological changes, including antibiotic, to Table 2.1, **Column 4** (“**Fever after 12** week of pregnancy (1cs)) and mark these relationships to the disease causing factor (fever in pregnancy) with letters **“R”** in this column accordinly.

### Mother with Diabetis and Obesity as a factor

1. A study on TNF-Alfa and obesity and diabetes states: “Recent data have suggested a key role for tumor necrosis factor (TNF)-α in the insulin resistance of obesity and non-insulin-dependent diabetes mellitus (NIDDM). **TNF-α expression is elevated in the adipose tissue of multiple experimental models of obesity**. Neutralization of TNF-α in one of these models **improves insulin sensitivity** by increasing the activity of the insulin receptor tyrosine kinase, specifically in muscle and fat tissues.”(*“Tumor Necrosis Factor α: A Key Component of the Obesity-Diabetes Link.”*, Gökhan S Hotamisligil, Bruce M Spiegelman; Diabetes 1 November 1994; 43 (11): 1271–1278. https://doi.org/10.2337/diab.43.11.1271). We add TNF-alfa increase as physiological paramater to the most left column of Table 2.1 and mark the Column 9 (**“Mother with Diabetis + Obesity (1cs)**”) with letter “R”.
2. Another research on obesity states: “In **obese individuals, there is a significant relation between BMI and adipose tissue TNF-α mRNA levels**.<u>^10^</u> Similarly, there was a significant correlation between BMI and plasma TNF-α levels in the current study. Weight reduction is associated with decreased TNF-α mRNA expression in adipose tissue.”, and also “**High plasma levels of TNF-α were found to be associated with increased triglyceride levels and a low HDL cholesterol.** This relation is also found in patients with early-onset coronary heart disease,<u>^15^</u> another group with an overrepresentation of IR. Similarly, there is a **significant correlation between adipose tissue TNF-α mRNA expression and plasma triglycerides in obese individuals**” (*“Relation Between Plasma Tumor Necrosis Factor-α and Insulin Sensitivity in Elderly Men With Non–Insulin-Dependent Diabetes Mellitus”*, Jan Nilsson, Stefan Jovinge, et al, Arteriosclerosis, Thrombosis, and Vascular Biology. 1998;18:1199–1202 https://doi.org/10.1161/01.ATV.18.8.1199)
3. Another study on Obesity and its biomarkers informs: **“Interleukin-6 is a pro-inflammatory cytokine that is also involved in insulin resistance found in obesity**. IL-6 levels correlates positively with adiposity in humans, wherein, obese individuals have high IL-6 levels and weight loss promotes reduced production of this cytokine”. We add IL-6 and insulin resistance as physiological parameters related to obesity in Table 2.1, Column 9 ((**“Mother with Diabetes + Obesity (1cs)**”) and mark the relationship with letters “R” in this column accordingly. The same study informs about other physiological biomarkers observed in obesity: “**Mechanisms involved in insulin resistance and inflammation present in obesity RBP4**, retinol binding protein-4; **TNF-a,** tumor necrosis factor-a; **IL**, interleukin; **MCP-1**, monocyte chemotactic protein-1; **SFRP5**, secreted frizzled-related protein 5; **LPS**, lipopolysaccharide; **TLR-4**, toll-like receptor 4; **JNK**, c-jun amino (N)-terminal kinase; **STAT3**, signal transducer and activator of transcription 3; NF-?B, nuclear factor kappa B; IKK, I kappa B kinase; IRS-1, insulin receptor substrate-1; IR, insulin receptor; TRAF1, tumor necrosis factor receptor-associated factor 1; GLUT4, translocation of glucose transporter 4; SOCS-3, suppressor of cytokine signaling protein-3; **TRAF**, tumor necrosis factor receptor-associated factor; **IRAK**, interleukin 1 receptor-associated kinase; **PI3K**, phosphoinositide 3-kinase; **JAK**, janus kinase; TAK, transforming growth factor (TGF)-activated kinase ß; CD14, cluster of differentiation-14; **MD2**, Myeloid differentiation protein-2.”. The same study states: “ Studies have **shown that adipose tissue from lean individuals secretes larger amounts of SFRP5**. Moreover, this protein acts by r**educing the production of pro-inflammatory adipokines, such as tumor necrosis factor-α (TNF-α), interleukin-6 (IL-6), and monocyte chemotactic protein-1 (MCP-1).”** We are adding this parameters to left most column of Table 2.1 and mark them with letters “R” in the Column 9 (**“Mother with Diabetes + Obesity (1cs)**”)

### Low iron during pregnancy as a factor

1. An article about iron metabolism states:“**When intracellular iron levels become low**, **the expression of transferrin receptor on the surface of the cell is upregulated** and the **synthesis of ferritin is downregulated** by so-called iron regulatory proteins(IRPs) that bind directly to special loop-like binding sites of the respective messenger RNAs (mRNAs).” (*“Iron Metabolism, Iron Deficiency and Anaemia.From Diagnosis to Treatment and Monitoring.”*, Rolf Hinzmann, Ph.D., Sysmex Journal International, Vol.13 No.2 (2003)). We adding this physiological parameters to the most left column and add letters “R” to **Column 10** (**“Low Iron Intake in Pregnancy over 35(1cs)”)**
2. An article on iron deficiency biomarkers states: “In the healthy population, serum ferritin correlates well with iron stores assessed via liver biopsies **. Serum ferritin is therefore a favoured marker for diagnosing iron-linked disorders.”** and also that **“In iron deficiency, more transferrin is synthesised to enhance iron transport. ”** (*“Diagnosing iron deficiency: Controversies and novel metrics*”, Jody A. Rusch, Diederick J. van der Westhuizen, et al, Best Practice & Research Clinical Anaesthesiology, Volume 37, Issue 4,) We adding this physiological parameter to the most left column and add letters “R” to **Column 10** (**“Low Iron Intake in Pregnancy over 35(1cs)”)**
3. An article on Iron deficiency and glucose metabolism states: “An earlier study showed **that reduced iron stores have a link with increased glycation of hemoglobin A1C (HbA1c)**, leading to false-high values of HbA1c in non-diabetic individuals. ” and also that: “Hashimoto et al. (2) demonstrated tha**t the HbA1c**, but not serum glycated albumin, is **elevated in late pregnancy in 47 nondiabetic pregnant women not receiving iron supplementation**, mean corpuscular hemoglobin (MCH) decreased from 29.9 ± 1.8 pg to 28.7 ± 2.7 pg, due to iron deficiency. **Their Hb A1C levels showed a negative correlation with mean corpuscular hemoglobin (MCH), serum transferrin saturation, and serum ferritin.** ”. (*“Iron deficiency anemia and glucose metabolism”*, Ashraf T. Soliman1, Vincenzo De Sanctis, et al, Acta Biomed 2017; Vol. 88, N. 1: 112-118 DOI: 10.23750/abm.v88i1.6049). We see that several studies show a decrease of iron with increase in Hb1A1C.We place “Hb1A1C Increase” in the most left column and put a letter “R” in **Column 10** (**“Low Iron Intake in Pregnancy over 35(1cs)”)** to mark this relationship.
4. Studies on Hb1A1C showed a significant correlation with Insulin Resistance measure (HOMA-IR) ≥2.5: “Heianza et al. reported that in subjects **without a history of diabetes a HbA_1c_ level of >5.9% were significantly associated with higher HOMA-IR values** [17]. In another study Borai et al. showed that the **correlation between HbA_1c_ and insulin resistance were higher in subjects with normal glucose tolerance** than in patients with pre-diabetes and diabetes ”(*“The independent relationship between hemoglobin A1c and homeostasis model assessment of insulin resistance in non-diabetic subjects*“, Yalcin, Hulya, Toprak,et al,Turkish Journal of Biochemistry, vol. 42, no. 1, 2017, pp. 31-36. https://doi.org/10.1515/tjb-2016-0256). We can use **HbA1C as an estimate of increased insulin resistance.**
5. A study on insulin resistance metrics (HOMA-IR) and biological age (BA) informs: **“**This study is the first to **report that HOMA-IR was positively correlated with BA** and advanced aging after adjusted for covariates. ” (*“HOMA-IR is positively correlated with biological age and advanced aging in the US adult population.”*, Yang, H., Gong, R., Liu, M. *et al.*, *Eur J Med Res* **28**, 470 (2023). https://doi.org/10.1186/s40001-023-01448-1). We put a physiological parameter **“HOMA-IR increase”** in left most column of Table 2.1 and put this dependency as letter “R” in Column 10 for women older than 35 years(**“Low Iron Intake in Pregnancy over 35(1cs)”).** We see that **women older 35 are more insulin resistive than younger women.**

### Air pollution as a factor

1. An article on organic dust exposure (ODE) informs: “Cytochrome c, a peripheral protein of the mitochondrial inner membrane (IMM), is known to function as an electron shuttle between complex III and complex IV of the respiratory chain (<u>Cai et al. 1998</u>). Its activity and its release from the IMM has been implicated in caspase activation and mitochondrial outer membrane permeabilization (MOMP), leading to cell death (<u>Garrido et al. 2006</u>). In our findings, **we observed that upon ODE exposure, there is an increase in cytosolic cytochrome c** and a **deficiency in the levels of COX4i2** (COX subunit 4 isoform 2), a terminal enzyme in the OXPHOS machinery. **Loss of COX4i2 results in decreased COX activity and decreased TAP levels** ”(*“Organic dust exposure induces stress response and mitochondrial dysfunction in monocytic cells”*., Mahadev Bhat S, Shrestha D, Massey N, et al, Histochem Cell Biol. 2021 Jun;155(6):699-718. doi: 10.1007/s00418-021-01978-x.).We add the physiloigical parameters impacted in organic dust exposure to the most left column and add its relationship to **“Air Polution in 1^st^ year”** with letters “R” in **Column 11**. The same article also informs on role of Ca^2+^ ions: ”…our data showing **caspase-1 activation along with the increase in Ca^2+^ levels on ODE exposure** (<u>Yu et al. 2014</u>). This would in turn lead **to IL-1β activation** and release into the extracellular space.”. We add caspase-1 activation, increase Ca^2+^ levels and IL-1β activation as physiological changes in the most left column (if not there yet) and mark its relationship with “Air Polution in 1^st^ year” with letters “R” in **Column 11**.
2. A study of particulate matter (PM) states: “**subway PM generates more reactive oxygen species (ROS)** a**nd oxidative stress** related outcomes compared to other PM, although a direct evidence for the clinical significance of ROS generation *in vivo* is limited”. We can put **increase of ROS** as a physiological parameter related to exposed to dust in the most left column and put letter “R”in **Column 11** (“Air Polution in 1^st^ year”) (*“Particle and metal exposure in Parisian subway: Relationship between exposure biomarkers in air, exhaled breath condensate, and urine.”*, I. Guseva Canu, C. et al, International Journal of Hygiene and Environmental Health,Volume 2372021,113837,ISSN 1438-4639, https://doi.org/10.1016/j.ijheh.2021.113837 )
3. A study on a particulate matter (PM) in classrooms states: “ A **significant association was found between indoor air concentrations (PM2.5 and PM10) with TNF-alpha level**. Children from rural areas are exposed to less air pollutants compared to those from urban area and this study also suggests **that higher exposure to PM2.5, PM10 and NO2 are associated with increasing of TNF-alpha level**.”(*“Tumor necrosis factor-alpha as biomarkers of exposure to indoor pollutants among primary school children in Klang Valley.”*, Jalaludin, Juliana and Syed Noh, et al., American Journal of Applied Sciences, 11 (9). pp. 1616-1630. ISSN 1546-9239; ESSN: 1554-3641). We can place dependancy between higher level of PM and TNF-Alfa level as “R” letter in **Column 11** (**“Air Pollution in 1^st^ year”**).

### Father older 45 year as a factor

1. A study on impact of father’s age on offspring states: **“Aging leads to more DNA replications** during spermatogenesis in testicles. **It increases the risk of copy error mutations such as small deletions and insertions**.It is estimated that spermatic chromosomes are replicated 35 times by age 15, 150 times by age 20, 380 times by age 30, 610 times by age 40, and 840 times by age 50 “(*“Paternal age is affected by genetic abnormalities, perinatal complications and mental health of the offspring.”*,Janeczko D, Hołowczuk M, et al, Biomed Rep. 2020 Mar;12(3):83-88. doi: 10.3892/br.2019.1266.). We add the “Gene Mutations” as a physiological parameter in the most left column of Table 2.1 and mark the relationship with a factor **“Father older 45 year (1cs)”** in **Column 5** by putting letter “R” in this column across the parameter.
2. Another study on impact of father’s health on pregnancy states: “Advanced **paternal age has also been shown to influence patterns of foetal and placental growth**. A study examining singleton pregnancies between 1999 and 2009 in Norway discovered **higher placental weight as well as higher placental to birth weight ratios from men over 50** than from men aged 20–24 years (Strom-Roum et al. 2013). ” (*“REPRODUCTIVE TOXICOLOGY: Impacts of paternal environment and lifestyle on maternal health during pregnancy”*, Afsaneh Khoshkerdar,Ece Eryasar, et al, Society for Reproduction and Fertility 2021, Volume 162: Issue 5, https://doi.org/10.1530/REP-20-0605). We add this physiological changes as **higher placental weight** as well as **higher placental to birth weight ratios** to the most left column of Table 2.1 and put letters “R” in **Column 5** to mark a relationship with the appropriate disease causation factor for Autism “Father older 45 year (1cs)”. The same study also informs: **“**The **KCNQ1OT1 imprinting control region**, and the paternally expressed long ncRNA Kcnq1ot1 derived from it, are responsible for the regulation of several imprinted genes required for normal placental development in both humans and mice (<u>Oh-McGinnis *et al.* 2010</u>).In mice, **advanced paternal age (approximately equivalent to 40–50 years in men) is associated with general DNA hypomethylation of this region in the developing placenta** when compared to tissue derived from the same males at a younger age (Denomme *et al.* 2020). ”. We add **DNA hypomethylation of KCNQ1OT1** as a physiological parameter change in the most left column and place letter “R” in **Column 5** to signify a relationship with a disease causation factor for Autism -“Father older 45 year (1cs)”.

### Child born within 1 year of pregnancy as a factor

1. A study on short inter pregnancies intervals (IPI) inform: “**Short IPIs may leave women with insufficient time to lose excess weight gained during pregnancy.** Maternal **overweight or obesity may result in increased risk of inflammatory up-regulation** [42], and increased levels of inflammatory proteins (cytokines) may lead to cervical ripening.”(*“Interpregnancy intervals and adverse birth outcomes in high-income countries: An international cohort study*“, Gizachew A. Tessema, M. Luke Marinovich,Siri E. Håberg, et al, PLOS ONE Published: July 19, 2021 https://doi.org/10.1371/journal.pone.0255000). We can add “Obesity and Overweight” and “Inflammation Upregulation” as physiological changes to the most left column and put letters “R” appropriately across in the **Column 7** (**“Child born within 1^st^ year of previous pregnancy (1 cs)”)**
2. A study on inter pregnancies interval and low birth weight(LBW) states**: “**Univariate and stratified analyses showed that **the risk for LBW was lowest when the interpregnancy interval was 18-23 months, and increased with shorter or longer intervals”** (*“Effect of Interpregnancy Interval on Infant Low Birth Weight: A Retrospective Cohort Study Using the Michigan Maternally Linked Birth Database”*, Bao-Ping Zhu, Thu Le, Maternal and Child Health Journal 7(3):169-78 DOI:10.1023/A:1025184304391). We place **low birth weight risk** as physiological parameter in the most left column of Table 2.1 and put letter “R” accordingly in **Column 7** (“Child born within 1^st^ year of previous pregnancy (1 cs)”) signify a relationship between this parameter and the disease causation factor for Autism.
3. A study shows: “**Women with short interpregnancy intervals had an average haemoglobin level of 10.9g/dl** (standard deviation=1.16g/dl), while those with **optimal interpregnancy intervals had an average level of 11.3g/dl** (standard deviation=1.32g/dl). ” (*“Short interpregnancy interval and adverse pregnancy outcomes among women in a Middle Eastern country”*, Amira Abdullah Saleh Al-Rumhi, Judie Arulappan, et al, BJM 02 June 2023, ISSN (online): 2052-4307). We place **haemoglobin level reduction** as physiological parameter in the most left column of Table 2.1 and add letter “R” in the **Column 7** (“Child born within 1^st^ year of previous pregnancy (1 cs)”). The same research also states: “Analysis of the prevalence of adverse pregnancy outcomes showed **iron deficiency anemia (56.8% vs 35.8%), gestational diabetes mellitus (31.7% vs 29.6%)**, postpartum haemorrhage (10.6% vs 2.8%), preterm premature rupture of membrane (3% vs 1.5%), preterm birth (13.6% vs 5.8%) and **low birth weight (16.5% vs 6.4%) were more prevalent among women with short interpregnancy intervals**”. We add **iron deficiency risk** as physiological parameter to the most left column and put letter “R” to the **Column 7** (“Child born within 1^st^ year of previous pregnancy (1 cs)”)
4. A study on interpregnancy and folate states: “During pregnancy, folate is mobilized from maternal stores to meet the increasing demands of mother and child. **If dietary supply is low, concentrations begin to decline from the fifth month of pregnancy onward**, and they continue to decline until several weeks after delivery. Repletion of stores may then take several months, and thus **mothers who conceive a subsequent child within these first months after delivery are at greater risk of folate deficiency**.”(*“Association between short interpregnancy intervals and term birth weight: the role of folate depletion.*”,Manon van Eijsden, Luc JM Smits, et al,The American Journal of Clinical Nutrition, Volume 88, Issue 1, 2008,Pages 147-153,ISSN 0002-9165,https://doi.org/10.1093/ajcn/88.1.147.). We add **folate deficiency** as physiological parameter in the most left column and add letter “R” to **Column 7** (“Child born within 1^st^ year of previous pregnancy (1 cs)”).
5. Another study on iron deficiency informs: “Furthermore, women with **short interpregnancy intervals generally have insufficient time to restore the required iron needs** before entering a subsequent pregnancy.More than 1000 published papers have linked low iron stores during pregnancy to adverse pregnancy outcomes in pregnant women and their offspring.”*(“Overview of iron deficiency and iron deficiency anemia in women and girls of reproductive age”,* Richard J. Derman, Anmol Patted,Gynecology and Obsterics, 04 August 2023 https://doi.org/10.1002/ijgo.14950)

### Having one child with Autism as a factor

1. A study on mothers of autistic children report: “Several unique bacterial biomarkers, such as Alcaligenaceae and *Acinetobacter*, were identified. **Mothers of ASD children had more Proteobacteria, Alphaproteobacteria, Moraxellaceae, and *Acinetobacter* than mothers of healthy children**.”(*“Correlation of Gut Microbiome Between ASD Children and Mothers and Potential Biomarkers for Risk Assessment.”*, Li N, Yang J, Zhang J, et al, Genomics Proteomics Bioinformatics. 2019 Feb;17(1):26-38. doi: 10.1016/j.gpb.2019.01.002). We add **Proteobacteria, etc difference** in the most left column of Table 2.1 and add letter “R” to Column 8 (“Having one child with Autism (2cs”)

This way we finished populating Table 2.1 with physiological parameters related to disease causation factors and marked the relationships between them with letters “R”. Let’s find out where physiological parameters related to these factors which are causing disease are intersecting. By looking at the Table 2.1 we can see that next physiological parameters intersect and we can build matrix **Pm** from them as below. As reminder, we call an intersection a place where 2 physiological parameters are the same for 2 different factors. The intersections of physiological parameters in Table 2.1 are marked in gray color.

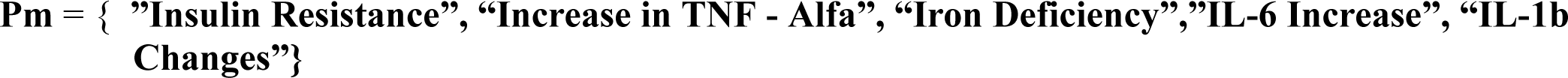

We see that we got 5 physiological parameters in matrix **Pm** but we can have only 3 as the disease is caused by 3 physiological parameters’ changes beyond 1-sigma as it was determined using formula 1.0 in this article. It means we need to eliminate redundant parameters using the rules and algorithm provided in explanation section of the method.

We notice that a disease causation factor “Depression in Women” is impacting physiological parameters such as **”Insulin Resistance”**, **“Increase in TNF - Alfa”**,**”IL-6 Increase”**, **“IL-1b Changes”** (see Table 2.1) which cannot be all 1-sigma changes as the this disease causation factor can only change 1 physiological parameter beyond 1-sigma according to a determination done above per a Disease Causation criteria. We can see that a physiological parameters **“Increase in TNF - Alfa”** has 5 factors’ intersections and this means it is extremely unlikely this is an *random* intersection. The intersection in **”Insulin Resistance”** also cannot be ignored as there are 3 factors which intersect in this physiological parameter, so it is very unlikely it is *a random* intersection as well. The physiological parameters **“IL-6 Increase”** and **“IL-1b Changes”** both have 2 factors intersections. We can try to eliminate them as less probable and build another candidate matrix Pm which now has only 3 physiological changes as required:

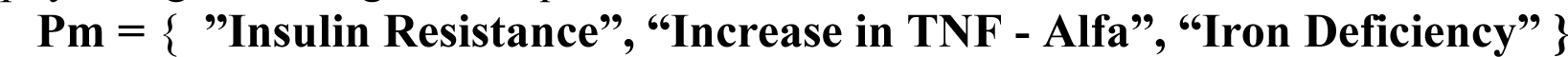

Having this new *candidate* matrix Pm we still see that a disease causation factor “Depression in Women” is impacting 2 physiological parameters **”Insulin Resistance”** and **“Increase in TNF - Alfa”** but we can notice these are 2 dependent on each other parameters as research shows that TNF-alfa does increase an insulin resistance. We also take in a consideration that multiple research show an insulin increase in Depression but not all research shows TNF-alfa increased in Depression, so we can remove the TNF-alfa parameter out of the impacted beyond 1-sigma by “Depression in Women” disease causation factor as 1^st^ the change *not always observed* and 2^nd^ as it is *connected to the observed an insulin resistance* increase. In this case, a “Depression in Women” disease causation factor is impacting only 1 physiological parameter which is **“Insulin resistance”**.

Why do both physiological parameters **”IL-6 Increase”** and **“IL-1b Changes”** have 2 factors’ intersections? This is likely because these cytokines are related to an increase in TNF-alfa, they dependent on it.

We also can see that a disease causation factor **“Mother with Diabetes + Obesity (1cs)”** according to Table 2.1 is causing changes in these physiological parameters **“Increase in TNF - Alfa”**, **”Insulin Resistance”** and **”IL-6 Increase”** but it can change beyond 1-sigma only 1 physiological parameter. We need to find the correct physiological parameter which impacted by this disease causation factor.

In order to do this we look into the already known to us research by Li M, et al (*“The Association of Maternal Obesity and Diabetes With Autism and Other Developmental Disabilities.”*, Li M, Fallin MD, Riley A, Landa R, et al, Pediatrics. 2016 Feb;137(2):e20152206. doi). This research has shown (see **Figure 2.1**) that non-obese pregnant women with diabetes (either with GDM or PGDM) have a risk of having Autistic child as **HR = 1.44** and **HR=1.32** accordingly and according to Disease Causation criteria these HRs are *not* within a range of **4.5+/-50%**(2.27-6.81) and so they are not factors which are causing Autism. The only disease causation factors we see in this research are combinations of Obesity and Diabetes (either with GDM or PGDM) where values RR=∼**HR=3.04** and RR =∼ **HR = 3.91** are both within a range of **4.5+/-50%**(2.27-6.81) of Disease Causation criteria. So only *Obesity together with Diabetes* during pregnancy is 1 cause of Autism.

According to a research on obesity and diabetes by Alzamil:”**Serum TNF-*α* levels in obese diabetic patients were significantly higher than in nonobese diabetic patients**. The *obese diabetic patients* have significant higher serum TNF-*α* levels than the *obese nondiabetic* group”, see **Figure 2.2** (*“Elevated Serum TNF-α Is Related to Obesity in Type 2 Diabetes Mellitus and Is Associated with Glycemic Control and Insulin Resistance”*,Hana Alzamil, Journal of Obesity, vol. 2020, Article ID 5076858, 5 pages, 2020. https://doi.org/10.1155/2020/5076858). We see that for obese people *with* diabetes HOMA-IR exceeds the values of HOMA-IR for obese people *without* diabetes by approximately. 2 pg/mL, which is a significant difference. The same study states that “The **level of TNF-*α* has a strong positive correlation** with HbA1c and **was positively associated with insulin resistance.**”. This association between TNF-alfa and insulin resistance is likely causing simultaneous changes in TNF-Alfa and insulin resistance for disease causation factor **“Mother with Diabetes + Obesity ”**.

At the same time the insulin resistance (measured as HOMA-IR) is high for overweight people ***without*** diabetes as **3.2+/-0.28** (*“Expanded Normal Weight Obesity and Insulin Resistance in US Adults of the National Health and Nutrition Examination Survey”*, Keilah E. Martinez, et al, Journal of Diabetes Research, vol. 2017, Article ID 9502643, 8 pages, 2017. https://doi.org/10.1155/2017/9502643) and can be more higher in people with diabetes (measured as HOMA-IR) **4.15 ± 3.56** (*“Insulin Resistance in Type 2 Diabetes Mallitus by Homa-IR Score”*,Ashish Purohit, Varsha Tiwari Int J Med Res Rev, 2015Feb.28 doi.org/10.17511/ijmrr.2015.i1.02) but according to a Disease Causation criteria “Overweight people without diabetes” or “People with diabetes but not Overweigh” risk factors cannot cause Autism as they risk are not within the required range so the Insulin Resistance changes related to them are not high enough to cause Autism.

In other words we found that only mothers having ***both*** Diabetes and Obesity are disease causation factor for Autism and this factor is significantly changing TNF-alfa which strongly positively correlate with Insulin Resistance but insulin resistance is not causing Autism neither in Obese *only* nor Diabetic *only* mothers despite pretty high values and so their combination is unlikely too. Due to this it is very likely that disease causation factor **“Mother with Diabetes + Obesity”** is causing a change to TNF-alfa in serum beyond 1-sigma in individual and not the Insulin Resistance increase beyond 1-sigma.We can remove “Insulin Resistance” physiological parameter as impacted beyond 1-sigma from “Mother with Diabetes + Obesity” disease causation factor for Autism (adjust a bold frame in Table 2.1).

**Figure 2.1:**
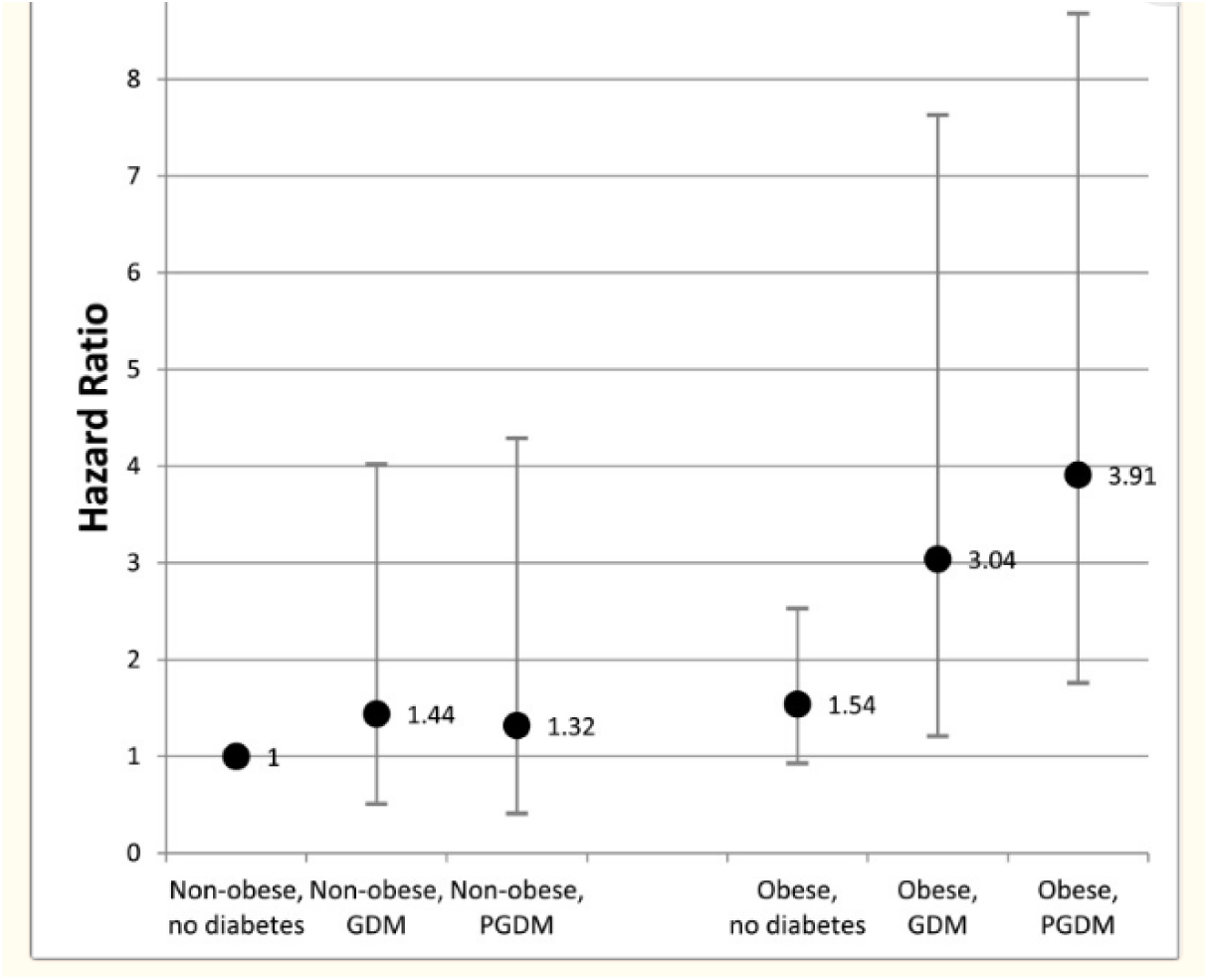
(Source:“The Association of Maternal Obesity and Diabetes With Autism and Other Developmental Disabilities.”, Li M, Fallin MD, Riley A, Landa R, et al, Pediatrics. 2016 Feb;137(2):e20152206. doi). Risk of Autism as HR for cases of Non-Obese mothers, Obese mothers, mothers with Diabetes (GDM and PGDM).

In a nutshell, to eliminate redundant physiological parameters from a matrix **Pm** we used few rules mentioned in method’s explanation, one of which is to eliminate a physiologically dependent parameters, and another rule was to find which physiological changes is less then 1-sigma. Now we can write the final matrix **Pm** which will look like:

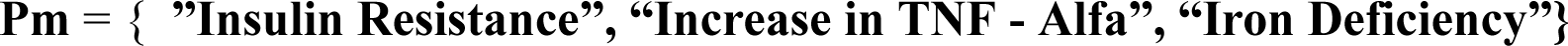

We have eliminated redundant physiologic a parameters **”IL-6 Increase”, “IL-1b Changes”** and explained that there is a relationship between TNF-Alfa and Insulin Resistance which causing observation of these 2 physiological changes simultaneously when a disease causation factor “Mother with Diabetes + Obesity ” is present.

The matrix **Pm** means that simultaneous coexistence of these 3 physiological changes beyond 1-sigma should cause Autism in girls and only 2 of these parameters are needed to cause Autism in boys (as we determine in this article the “Iron Deficiency” is likely needed to cause Autism in girls).

The rows in Table 2.1 which are **surrounded by bold rectangles** represent the final set of physiological parameters found as result of our analysis and parameters’ eliminations and these physiological parameters’ changes are causing Autism.

## Physiological Causes of Autism (Results of Analysis)

Based on the analysis of factors above the set of **3 physiological changes beyond 1-sigma which are taking place at the same time are causing a Autism is:**

1. Insulin Resistance increase beyond ∼1-sigma
2. Increase in TNF - Alfa beyond ∼1-sigma
3. Iron Deficiency (low iron)beyond ∼1-sigma

A child will have Autism if the disease causation factors change 2 of these physiological parameters beyond 1-sigma (actually, slightly less) for boys or 3 of them for girls. In other words a boy will get sick with Autism if Insulin Resistance will be increased beyond 1-sigma **and** TNF-alfa level will be increased beyond 1-sigma simultaneously and long enough. The triggering of Autism is not optional but *a must* as long as disease causing physiological changes are not removed fast enough.

## Validation of Method’s Results for Autism Causes

Now we need to move to a validation phase of our method. Let’s see what the existing research says about the physiological parameters we found. Please note the purpose of validation is not to prove that research used in this validation confirms by itself the causes of the disease. The purpose is to confirm we did not make any errors in using the algorithm by **verifying how consistent our results with the existing research**. In other words if we would be 100% sure we did not make any error in using a method and populated all data correctly in the tables, etc. then the confirmation would not be needed. The algorithm itself gives valid results.

A study on cytokines changes in Autism states: “Although a recent meta-analysis has reported that mainly IL-1β and IL-6 levels are significantly increased in ASD, **a number of studies have reported that TNF-α concentrations are also increased in blood**, **cerebrospinal fluid** or post-mortem brain tissue. Our study has confirmed these latter findings in our cohort of subjects and importantly that **blood concentrations of TNF-α are positively associated with ASD symptom severity**, thus indicating that TNF-α could be an important cytokine biomarker in ASD.” (*“Immunological cytokine profiling identifies TNF-α as a key molecule dysregulated in autistic children.”*, Xie J, Huang L, Li X, Li H, et al, Oncotarget. 2017 Jul 18;8(47):82390-82398. doi: 10.18632/oncotarget.19326. PMID: 29137272; PMCID: PMC5669898.). We can see this study is consistent with our finding of **Increase in TNF - Alfa** as one physiological cause of Autism. Important to notice here that this study also found that TNF-alfa blood concentrations in typically developed children have Mean = **8.77 pg/ml** (SD = 3.168) and that Autistic Children have TNF -alfa level have Mean = **12.15 pg/ml** (SD = 4.627). As we know a disease is triggered by physiological parameter change approximately beyond 1-sigma and this value would be 8.77 + 3.168 = **11.938 pg/ml.** We see that in ASD subject the mean value of TNF-alfa is **12.15 pg/ml** which is exceeding the 1-sigma**. It is an additional confirmation that TNF-alfa increase we found is correct** value.

A study on Diabetes in Obese women states: “**Maternal insulin resistance in obese women appears to negatively affect neurodevelopment at 2 years in their children** compared to children born to non-obese women. The impact appears non-global, specifically affecting motor development and domains such as attention, raising the possibility that specific regions of the fetal brain may be more sensitive to abnormal glucose metabolism than others.”(*“Impact of maternal insulin dependent diabetes mellitus (IDDM) on off-spring neurodevelopment”*, Alison G. Cahill, Amit M. Mathur,et al, AJOG, AUGUSTUS V-VI| VOLUME 216, ISSUE 1, SUPPLEMENT, S56, JANUARY 2017, https://doi.org/10.1016/j.ajog.2016.11.965).

Another study tested Insulin Resistance(IR) in ASD children and states: “In this exploratory investigation, we tested the hypothesis that young individuals with ASD have higher HOMA-IR than peers without. **We found that HOMA-IR is significantly increased by 0.31 unit for the ASD status**, when an unconditional multivariable linear regression model was fitted in order to adjust for confounders and other determinants.”(*“Cross-sectional investigation of insulin resistance in youths with autism spectrum disorder. Any role for reduced brain glucose metabolism?”*,.Manco, M., Guerrera, S., Ravà, L. et al., Transl Psychiatry 11, 229 (2021). https://doi.org/10.1038/s41398-021-01345-3). The same study also states: “In **patients with obesity and T2D, peripheral IR** seems **paralleling brain IR**, i.e., the reduced glucose metabolism in the central nervous system (CNS). **Brain IR has been associated with reduced executive functioning**^7^, **inhibitory control, and cognitive flexibility** in young patients with obesity^8^ and implied in the pathogenesis of dementia in adults suffering T2D or Alzheimer disease^9^.”. We can see that this research is consistent with our finding that insulin resistance **(IR)** is one of the cause of Autism as it affects neurodevelopment.

A study on Iron deficiency states: “**Iron deficiency could cause numerous detrimental effects** in people, such as anemia, fatigue, atrophic glossitis, hair loss, etc., and is also associated with the risk of many diseases. For example, previous studies showed that iron deficiency was associated with an increased risk of type 2 diabetes, Parkinson’s disease, and cancers. **Maternity with a lower mean daily iron intake led to an increased risk of autism in offspring**. **Numerous studies have suggested that iron deficiency was a common phenomenon in autism**, and iron supplementary has been widely advised for pregnant women and children with finicky habitation.”(*“The causal association between iron status and the risk of autism: A Mendelian randomization study”*, Li Chen, Xingzhi Guo1, et al, Sec. Nutrition, Psychology and Brain Health, Volume 9 - 2022, https://doi.org/10.3389/fnut.2022.957600).This study is consistent with one cause of Autism found which is a Low Iron.

**Figure 2.2:**
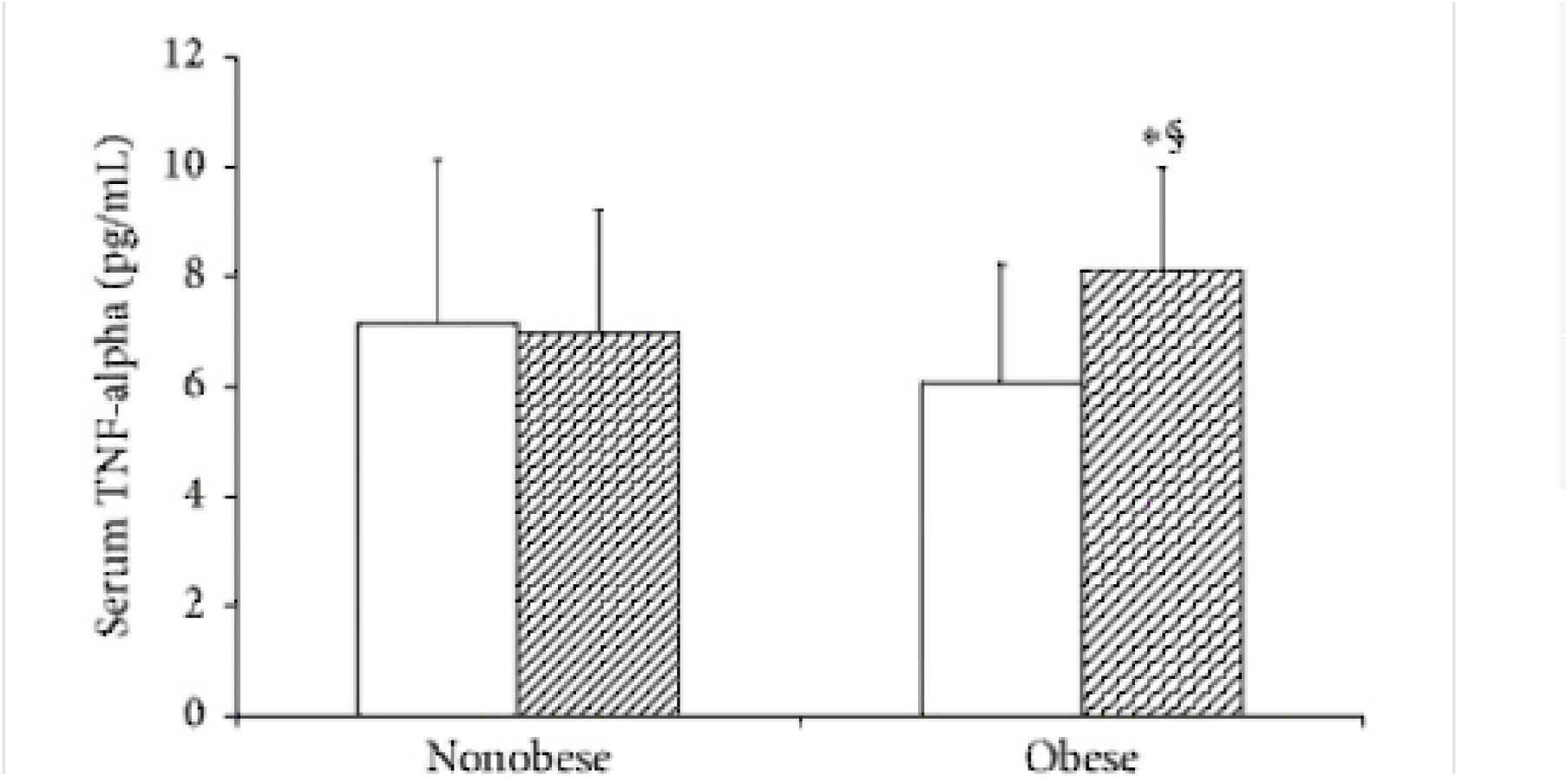
(Source: Alzamil, Journal of Obesity, vol. 2020) Effect of BMI on serum TNF-*α* levels (mean ± SEM). Nonobese nondiabetic group vs obese nondiabetic group;. §Obese diaetic patients vs nonobese diabetic patients (p<0.018). Nonobese nondiabetic group vs nonobese diabetic patients; Obese diabetic patients vs obese nondiabetic group (p<0.006). Gray areas on the right are people with diabetes.)

A study on Iron in Autistic children states: “The **mean value of serum iron levels in autistic children was severely reduced and significantly lower than in control children** (74.13 ± 21.61 μg/dL with a median 74 in autistic children 87.59 ± 23.36 μg/dL in controls) (*P* = 0.003).” (*“Iron and vitamin D levels among autism spectrum disorders children.”,* Bener A, Khattab AO, Bhugra D, Hoffmann GF., Ann Afr Med. 2017 Oct-Dec;16(4):186-191. doi: 10.4103/aam.aam_17_17. PMID: 29063903; PMCID: PMC5676409.). Which is also consistent with our finding of Low Iron, and also during pregnancy a fetus iron is dependent on iron transport through the placenta.

Another study on maternal iron deficiency impact on a child states: “**Lowered iron stores of the newborn child may persist for up to one year and result in iron deficiency anemia**.” and also “Iron is essential for neural metabolism and functioning. **Iron deficiency anemia results in changes in energy metabolism within the brain with defects in neurotransmitter function and myelination**. Therefore, infants and young children with **iron deficiency anemia are at risk of developmental difficulties involving cognitive, social-emotional, and adaptive functions**.Other studies have documented delays in both language and motor development.”: (*“The impact of maternal iron deficiency and iron deficiency anemia on child’s health.”*, Abu-Ouf NM, Jan MM., et al, Saudi Med J. 2015 Feb;36(2):146-9. doi: 10.15537/smj.2015.2.10289. PMID: 25719576; PMCID: PMC4375689). As we can see the study also confirm that iron deficiency impact brain functioning and development which is consistent with our finding that one cause of autism is a Low Iron.

We see our findings of physiological parameters changes beyond 1-sigma which are causing Autism such as ”Insulin Resistance increase”, “Increase in TNF - Alfa”, “Iron Deficiency(low iron)” are supported by existing research which was *not used* during the steps of presented method. Using the method we started off with a research on the different risk factors for Autism, then we moved to a research on variety of physiological parameters changes which are observed when the determined disease causation factors are present. The research on physiological changes was not generally related to Autism. At the end we have validated the results of the method with a separate known research which is consistent with our finding using the method. This way we can confirm the correctness of steps we took in executing the steps of this method for Autism. It also demonstrate how the method works.

Now we move result of analysis from **Table 2.1 to Table 2.2** and verify results of analysis above. As we can see from Table 2.2 we can determine which physiological parameter is impacted by a specific disease causation factor. For example, “Valproate usage”is a disease causation factor of Autism because it is causing Insulin Resistance to go beyond 1-sigma interval. The Valproate usage cannot cause Autism standalone without other 1 or 2 factors (for boys and girls accordingly). “High CNV + Ozone exposure” disease causation factor as per Table 2.2 is causing increase in TNF-Alfa beyond 1-sigma and that is why this factor is one of 3 causes of Autism though it cannot trigger the disease as standalone factor. We can see that disease causation factor as “Fever after 12 week of pregnancy OR high fever while pregnant”is also impacting TNF-alfa so it is increased beyond 1-sigma and that is why this factor is one of 3 causes of Autism but cannot cause the disease as standalone factor. Please note the disease causation factors in the Table 2.2 have a bit symbolic name and for exact meaning of the factor you need to check with research we provided in this article above when we went through the step of the method.

Now we can build Table 2.3 by moving data from Table 2.2 and grouping them by physiological parameters impacted into few columns. The **Table 2.3** below shows external factors which will **cause Autism if combined together** taken from 3 separate columns.

If the factors represent the 2 causes then the number of factors can be less accordingly. We can determine from the Table 2.3 that if a women had “Fever after 12 week of pregnancy” (Increase of TNF-alfa) and has “Depression” (Increase in Insulin Resistance) and has “Low iron” then a baby girl will get Autism if these physiological changes present long enough. Another combination to trigger Autism would be “Fever after 12 week of pregnancy” and “Depression” and “Child born within 1^st^ year of previous pregnancy”.

Please note the disease causation factors names are bit symbolic and for exact meaning of it we need to refer to a research we provided above when we went through the steps of analysis of Autism by this method. These disease causation factors are causing Autism because they change 3 physiological parameter beyond 1-sigma. As long as these or other factors combination do this they will trigger Autism. There can be many more factors found later but many of them will be changing these 3 parameters we found (our matrix **Pm**). There will be though some exceptions as it used to be more physiological parameters needed to cause Autism (this will be explained in more details in a section on the reason why Autism grows last few decades).

**Table 2.1.** (**In the header**: factors determined to be Disease Causation factors for Autism using a Disease Causation criteria. **In the most left column:** physiological parameter changes observed when the factors are present. “R” - signifies a relationship between such a physiological parameter change and a disease causing factor. Areas in gray are *intersections* of physiological parameters for few factors. *Bold frames* are final set of physiological parameters causing Autism.)

| Physiological parameters related to external factors | External factors causing Autism |  |  |  |  |  |  |  |  |  |  |
| --- | --- | --- | --- | --- | --- | --- | --- | --- | --- | --- | --- |
|  | Valproate (1cs) | Depression (1cs) | High CNV + Ozone Exposure (1cs) | Fever after 12 week of pregnancy or High Fever While Pregnant (1cs) | Father older 45 year (1cs) | The column intentionally left blank | Child born within 1<sup>st</sup> year of previous pregnancy (1cs) | Having one child with Autism (2cs) | Mother with Diabetes + Obesity (1cs) | Low Iron Intake in Pregnancy over 35(1cs) | Air Pollution in 1<sup>st</sup> year(1cs) |
|  | 1 | 2 | 3 | 4 | 5 | 6 | 7 | 8 | 9 | 10 | 11 |
| GABA potentiation (of receptor) | R |  |  |  |  |  |  |  |  |  |  |
| Proteobacteria, etc difference |  |  |  |  |  |  |  | R |  |  |  |
| Folate deficiency |  |  |  |  |  |  | R |  |  |  |  |
| Iron deficiency risk |  |  |  |  |  |  | R |  |  | R |  |
| Hemoglobin level reduction |  |  |  |  |  |  | R |  |  |  |  |
| Low birth weight risk |  |  |  |  |  |  | R |  |  |  |  |
| **Obesity and Over weight** |  |  |  |  |  |  | **R** |  |  |  |  |
|  | **1** | **2** | **3** | **4** | **5** | **6** | **7** | **8** | **9** | **10** | **11** |
| **Inflammation Upregulation** |  |  |  |  |  |  | **R** |  |  |  |  |
| **DNA hypomethylation of KCNQ10 T1** |  |  |  |  | **R** |  |  |  |  |  |  |
| **Higher placental to birth weight ratios** |  |  |  |  | **R** |  |  |  |  |  |  |
| **Higher placental weight** |  |  |  |  | **R** |  |  |  |  |  |  |
| **Gene Mutations** |  |  |  |  | **R** |  |  |  |  |  |  |
| **Cytosolic cytochrome c increase** |  |  |  |  |  |  |  |  |  |  | **R** |
| **COX4i2 Deficiency** |  |  |  |  |  |  |  |  |  |  | **R** |
| **COX activity decreased.** |  |  |  |  |  |  |  |  |  |  | **R** |
| **TAP levels decreased** |  |  |  |  |  |  |  |  |  |  | **R** |
| **Caspase-1 activation** |  |  |  |  |  |  |  |  |  |  | **R** |
|  | **1** | **2** | **3** | **4** | **5** | **6** | **7** | **8** | **9** | **10** | **11** |
| **Ca<sup>2+</sup> levels increase** |  |  |  |  |  |  |  |  |  |  | **R** |
| **Increase of ROS** |  |  |  |  |  |  |  |  |  |  | **R** |
| **IL-1β activation** |  |  |  |  |  |  |  |  |  |  | **R** |
| **HOMA-IR increase** |  |  |  |  |  |  |  |  |  | **R** |  |
| **Hb1A1C Increase** |  |  |  |  |  |  |  |  |  | **R** |  |
| **Ferritin is down regulated** |  |  |  |  |  |  |  |  |  | **R** |  |
| **Transferrin increased** |  |  |  |  |  |  |  |  |  | **R** |  |
| **Transferrin receptor up regulated** |  |  |  |  |  |  |  |  |  | **R** |  |
| **SFRP5 increase** |  |  |  |  |  |  |  |  | **R** |  |  |
| **Increased triglyceride levels** |  |  |  |  |  |  |  |  | **R** |  |  |
| **Low HDL cholesterol** |  |  |  |  |  |  |  |  | **R** |  |  |
| **GABA** |  |  |  |  |  |  |  |  |  |  |  |
| Block voltage gated sodium channels | R |  |  |  |  |  |  |  |  |  |  |
| Epigenetically inhibits histone deacetylase | R |  |  |  |  |  |  |  |  |  |  |
|  | 1 | 2 | 3 | 4 | 5 | 6 | 7 | 8 | 9 | 10 | 11 |
| GI Upset | R |  |  |  |  |  |  |  |  |  |  |
| Weight Gain | R |  |  |  |  |  |  |  |  |  |  |
| Foetal Oxidation | R |  |  |  |  |  |  |  |  |  |  |
| Antagonism of folate decreases DNA synthesis | R |  |  |  |  |  |  |  |  |  |  |
| Physiological parameters related to external factors | External factors causing Autism |  |  |  |  |  |  |  |  |  |  |
|  | valproate ( 1cs) | Depression ( 1cs) | High CNV + Ozone Exposure ( 1cs) | Fever after 12 week of pregnancy or High Fever While Pregnant ( 1cs) | Father older 45 year ( 1cs) | The column intentionally left blank | Child born within 1<sup>st</sup> year of previous pregnancy ( 1cs) | Having one child with Autism ( 2cs) | Mother with Diabetes + Obesity ( 1cs) | Low Iron Intake in Pregnancy over 35(1cs) | Air Pollution in 1<sup>st</sup> year(1cs) |
|  | 1 | 2 | 3 | 4 | 5 | 6 | 7 | 8 | 9 | 10 | 11 |
| Increase in Fast | R |  |  |  |  |  |  |  |  |  |  |
| Plasma Insulin (FPI) level |  |  |  |  |  |  |  |  |  |  |  |
| ROS increase |  |  |  |  |  |  |  |  |  |  | R |
| Insulin Resistance in Women (*HOMA-IR* increase) | R | R |  |  |  |  |  |  | R |  |  |
| Dyslipidemia | R |  |  |  |  |  |  |  |  |  |  |
| Type 2 of diabetes Mellitus | R |  |  |  |  |  |  |  |  |  |  |
| Increase in TNF-α | | R | R | R | | | | | R | | R |
| sICAM-1 increases |  |  | R |  |  |  |  |  |  |  |  |
| MDA increased |  |  | R |  |  |  |  |  |  |  |  |
|  | 1 | 2 | 3 | 4 | 5 | 6 | 7 | 8 | 9 | 10 | 11 |
| Reduced methylation of mitochondria |  |  |  |  |  |  |  |  |  |  |  |
| Respiratory Inflammation |  |  | R |  |  |  |  |  |  |  |  |
| Hypometh |  |  | R |  |  |  |  |  |  |  |  |
| ylation of Mithochondria D-loop |  |  |  |  |  |  |  |  |  |  |  |
| Hemoglobin |  |  |  |  |  |  | R |  |  |  |  |
|  | valproate (1cs) | Depression (1cs) | High CNV + Ozone Exposure (1cs) | Fever after 12 week of pregnancy or High Fever While Pregnant (1cs) | Father older 45 year (1cs) | The column intentionally left blank | Child born within 1<sup>st</sup> year of previous pregnancy (1cs) | Having one child with Autism (2cs) | Mother with Diabetes + Obesity (1cs) | Low Iron Intake in Pregnancy over 35(1cs) | Air Pollution in 1<sup>st</sup> year(1cs) |
|  | 1 | 2 | 3 | 4 | 5 | 6 | 7 | 8 | 9 | 10 | 11 |
| Virus & infections |  |  |  | R |  |  |  |  |  |  |  |
| Antibiotic for Listeria monocytogenes |  |  |  | R |  |  |  |  |  |  |  |
| Viral gastroenteritis |  |  |  |  |  |  |  |  |  |  |  |
| Pyelonephritis |  |  |  | R |  |  |  |  |  |  |  |
| Antibiotic Amoxicillin |  |  |  | R |  |  |  |  |  |  |  |
| Cytchrome C protein |  |  |  |  |  |  |  |  |  |  | R |
| increases (leads to cell death) |  |  |  |  |  |  |  |  |  |  |  |
| Proinflammatory markers increase |  | R |  |  |  |  |  |  |  |  |  |
| HPA axis overactive |  | R |  |  |  |  |  |  |  |  |  |
| High Cortisol |  | R |  |  |  |  |  |  |  |  |  |
|  | 1 | 2 | 3 | 4 | 5 | 6 | 7 | 8 | 9 | 10 | 11 |
| Altered metabolic profile |  | R |  |  |  |  |  |  |  |  |  |
| TNF - Alfa changes |  | R |  |  |  |  |  |  |  |  |  |
| IL-6 changes |  | R |  |  |  |  |  |  |  |  |  |
| IL-6 increase |  | R |  |  |  |  |  |  | R |  |  |
| IL-1b changes |  | R |  |  |  |  |  |  |  |  | R |
| BDNF reduced |  | R |  |  |  |  |  |  |  |  |  |
| Chemottractant probe |  | R |  |  |  |  |  |  |  |  |  |
| HDL Profile changes |  | R |  |  |  |  |  |  |  |  |  |
| Insulin Resistance |  | R |  |  |  |  |  |  |  |  |  |
| Albu<br>min |  | R |  |  |  |  |  |  |  |  |  |
| GAB<br>A |  | R |  |  |  |  |  |  |  |  |  |
| Hista<br>min |  | R |  |  |  |  |  |  |  |  |  |
| CRP<br>incre<br>ase |  | R |  |  |  |  |  |  |  |  |  |
| Insuli<br>n<br>chang<br>e |  | R |  |  |  |  |  |  |  |  |  |

## Finding a Physiological Change which a Disease Causing Factor is impacting

So far we could not find all physiological causes for some of the disease causation factors described in Table 2.1, for example for a disease causation factor **“Father older 45 year ”.**

*As we found physiological parameters which are causing Autism* (our matrix Pm) we can use it to find the physiological parameters for this disease causation factor and *same* can be done for other disease causation factors which will be found in future.. We see that our matrix Pm has 3 physiological parameters:

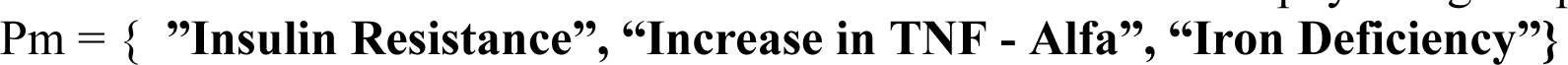

The father age can impact only 1 physiological parameter as we determined with Disease Causation criteria, it either impacts an **Increase of TNF Alfa** or **Insulin Resistance** or cause **Iron Deficiency.**

By cross checking with existing research we can find that it is TNF-alfa which is likely impacted by late fathers’ age.A research by E. Schjenken, et al states: “Detailed analysis of the molecular regulation of the female response to seminal plasma commenced in the 1990s and described a surge in production of several pro-inflammatory cytokines in rat and mouse uterus following mating (<u>191</u>, <u>364</u>, <u>368</u>, <u>399</u>). Key mediators induced in mouse uterus are tumor necrosis factor (TNF), interleukin (IL)-1β, IL-6, colony-stimulating factor 2 (CSF2), and CSF1. Chemokines that induce immune cells to extravasate from the peripheral blood are also elevated during the immediate post-coital phase.” (*“The Female Response to Seminal Fluid”*, John E. Schjenken, Sarah A. Robertson,American Physiological Society 17 APR 2020, https://doi.org/10.1152/physrev.00013.2018). As per research by Janeczko D, et al a father age is significantly increasing a chance of copy error mutations due constantly increased number of spermatic chromosomes replications. This is expected to increase immune reaction to male seminal fluid and even more increase TNF-alfa production

Another research informs: “Upon deposition of seminal plasma into the female reproductive tract a range of uterine cellular and immune cell changes are initiated which influence female reproductive physiology to increase the likelihood of conception.” (*“REPRODUCTIVE TOXICOLOGY: Impacts of paternal environment and lifestyle on maternal health during pregnancy”*, Afsaneh Khoshkerdar, Ece Eryasar, Society for Reproduction and Fertility 2021,Vol.162: Iss. 5, https://doi.org/10.1530/REP-20-0605).

This suggests that it is likely that a reason why Autism risk grows significantly with father age (starting age 45+) because it is causing more genetic mutations in fathers sperm and due to this a stronger immune response from female reproductive tract and in particular increase of TNF-alfa production which as we determined is a one physiological cause of Autism out of 3 required to trigger it. As we determined already that “Father older 45 year” is a disease causation factor for Autism and per discussion above it is likely causing TNF-alfa increase beyond 1-sigma in female reproductive tract but not Insulin Resistance increase beyond 1-sigma or Iron Deficiency beyond 1-sigma. We update Table 2.2 with this finding as it is likely and need to be investigated further.

The above is one example of capabilities of the presented method in mapping new disease causation factors to already found physiological changes found earlier.

## Why girls are less often sick with Autism (3-4 times less)?

The answer to the question why incidence of Autism in girls is much less we actually have found above. As per calculations using formula (1.0) we determined that boys have 2 simultaneous physiological parameters changes beyond 1-sigma and girls have 3 physiological changes beyond 1-sigma interval which trigger the disease in them. According to the article *“A Connection between Factors Causing Diseases and Diseases Frequencies: Its Application in Finding Disease Causes” (Alan Olan*, Journal of Clinical Trials, Vol.13, Issue 4.) the less number of physiological parameters is causing the disease the more the frequency (incidence rate) of the disease. In other words, girls are less frequently sick with Autism because they have 1 more protective physiological parameter and only if this parameter is changed beyond 1-sigma along with 2 others (as in boys) then girls become sick with Autism.

Now, we determined a 3 physiological changes which are causing Autism but which 2 causes are related to girls and boys and which one cause is related to girls onlyLet’s put these 3 causes again as matrix **Pm =** { **”Insulin Resistance ”, “Increase in TNF - Alfa”, “Iron Deficiency” }**

**Table 2.2.**
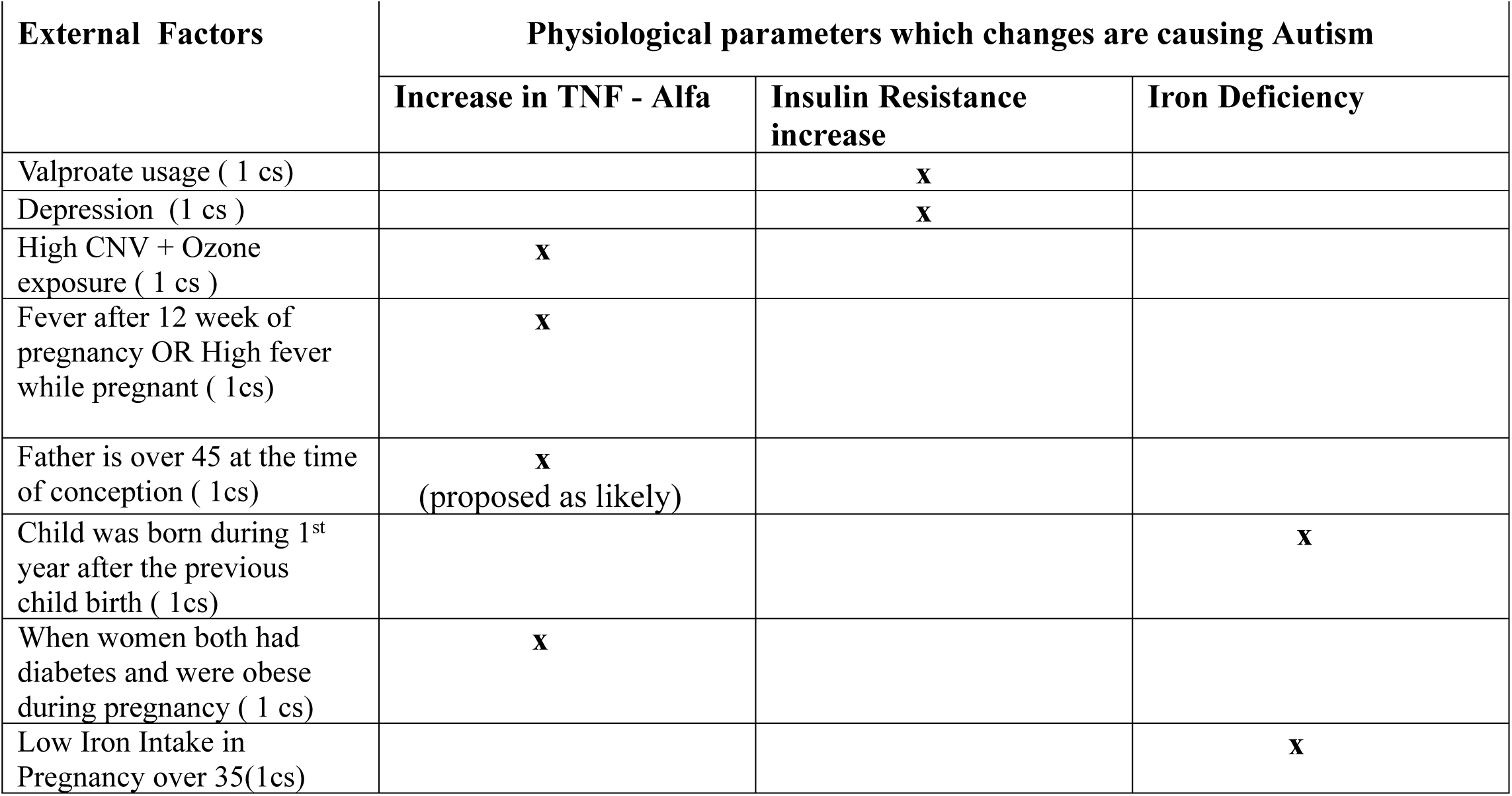
(**In the header:** 3 physiological parameters which change beyond 1-sigma if simultaneously present causes Autism. **In most left column:** some of Disease Causation factors for Autism which *only* can cause it if combined together so all 3 physiological parameters required to trigger Autism are changed. **1cs**, **2cs** (on the left, stand for a cause) - means the disease causation factor changes 1 or 2 physiological parameters beyond 1-sigma as determined by Disease Causation criteria. **x -** means that an Disease Causation factor is causing a change/related to the change in physiological parameter beyond 1-sigma so this parameter becomes one of 3 causes of Autism. **Note:** boys likely don’t require Iron Deficiency for triggering Autism)

To determine which parameters is impacting girls we use a research on insulin resistance (IR) in pregnancy which states: **“Maternal IR during mid-pregnancy was significantly higher in women carrying a female fetus** than in those with a male fetus.”(“*Fetal sex and maternal insulin resistance during mid-pregnancy: a retrospective cohort study.”*, Yamashita, H., Yasuhi, I., Koga, M. et al., BMC Pregnancy Childbirth 20, 560 (2020). https://doi.org/10.1186/s12884-020-03242-x). The insulin resistance increase observed is not related to Autism but it is a normal pregnancies process. So now :

**Table 2.3.** (**In the header:** 3 physiological parameters changes (*beyond approx. 1-sigma*) which should be simultaneously present to cause Autism. **In the columns:** Disease Causation factors which are either causing the physiological change beyond 1-sigma (or physiological change present if the factor exists). Any set of 3 Disease Causing factors (one from each column) from each column will cause Autism. The Disease Causation factors *cannot* cause Autism as standalone. **Note:** low iron is likely required only for girls)

| **Increase in TNF - Alfa** | **Insulin Resistance increase** | **Iron Deficiency** |
| --- | --- | --- |
| High CNV + Ozone exposure ( 1 cs ) | Valproate usage ( 1 cs ) | Low Iron intake during pregnancy for women over 35 |
| Fever after 12 week of pregnancy OR High fever while pregnant ( 1cs) | Depression ( 1 cs ) | Child was born during 1<sup>st</sup> year after the previous child birth ( 1cs) |
| When women both had diabetes and were obese during pregnancy ( 1 cs) |  |  |
| Father is over 45 at the time of conception ( 1cs) (suggested as likely) |  |  |

**NOTE:** the list of factors in **Table 2.3** is obviously not complete and provided as per author's knowledge of specific research, other factors can be added in the future if new research finds them, etc.. At the same time the future factors as current ones will be related to the physiological parameters in the columns of the table as these parameters the real cause of a disease (the factors are only causing their change).

1. We can hypothesize that to compensate for this insulin resistance increase **girls have acquired during evolution some unknown mechanism X.**
2. We have determined earlier in this article that one of causes of Autism is **Iron Deficiency** (including during pregnancy). According to the existing research an **iron level is closely connected to insulin resistance** and its indicators **HbA1c** and **HOMA-IR** and often negatively correlate with it. Ashraf T. Soliman1 et al states: “An earlier study showed **that reduced iron stores have a link with increased glycation of hemoglobin A1C (HbA1c)**, leading to false-high values of HbA1c in non-diabetic individuals. ” (*“Iron deficiency anemia and glucose metabolism*“,Ashraf T. Soliman1, et al, Acta Biomed 2017; Vol. 88, N. 1: 112-118, DOI: 10.23750/abm.v88i1.6049). The same author is mentioning another research on iron deficiency(ID): “Ozdemir A et al. evaluated the effects of correction of ID anemia (from Hb: 9.9 ± 1.8 g/dL to Hb: 13.1 ± 1 g/dL) on insulin secretion in 54 non-diabetic premenopausal women with IDA. **A statistically significant decreases were found** in fasting insulin levels and **homeostatic model assessment (HOMA) scores following correction of anemia in women** <40 years and normal body mass index (BMI <27 kg/m2) ”. Based on this we can hypothesize that the unknown factor X which is compensating Insulin Resistance in girls is **a regulation of iron**. If this is so then girls should become sick with Autism when 2 physiological parameters already beyond 1-sigma (as in boys) and the 3^rd^ parameter the **Iron Deficiency is falling below 1-sigma in girls or girls’ fetuses during pregnancies.** In other words, if our hypothesis is correct then **the 3^rd^ parameter which is causing Autism specifically in girls** (when it goes beyond 1-sigma) **is low iron in girls or in girls fetuses.**

From another side, if the Low Iron is not the 3^rd^ physiological change required to cause Autism *in girls* then these can be either Increased Insulin Resistance (above 1-sigma) or high level of TNF-alfa as they are the 2 other parameters required to cause Autism as it was determined using the presented method. Let’s analyze both of them. At 1^st^ let’s assume that 3rd parameter is TNF-alfa. The TNF-alfa can be easily increased in both girls and boys during infection so it is unlikely this physiological parameter could not potentially protect girls **more** than boys as they both easily impacted by its change. If 2 others are impacted to cause Autism, TNF-alfa would be easily impacted as well. Let’s assume now that this 3^rd^ physiological parameters protecting girls is an Insulin Resistance Increase (above 1-sigma). In this case boys would be getting sick with Autism because of Low Iron (below 1-sigma) and High TNF-alfa (above 1-sigma) but there is no clear connection between these 2 factors **if** Insulin Resistance is within 1-sigma (or does not exist at all) which would build a mechanism of disease pathology and this *mechanism is usually clearly observed after the use of this method* (as will be shown in a section for building hypothesis of Autism). So even this simple analysis points to the fact that 3rd parameter which is causing Autism in girls is likely Low Iron (below 1-sigma).

We can restate our hypothesis in a more short way. We hypothesize that **Iron level reduction** which is one of 3 determined causes of Autism **is the same mechanism** (which in this case broken) which **compensate for the increased Insulin Resistance in girls** (or girls’ fetuses during normal pregnancies) and which is **3**rd parameter which is required to cause Autism in girls.

We already determined using the presented method that Iron level reduction beyond 1-sigma is one cause of Autism and this hypothesis just explains that Iron level reduction is a physiological change which is required to trigger Autism in girls.

## Why Autism incidence rate significantly rose last few decades?

In the late 1980s, 3 or 4 babies out of 100,000 were diagnosed with Autism. Using a formula (1.0) let’s calculate the number of causes for Autism at this time. This is an estimate as the data may vary depending on source.

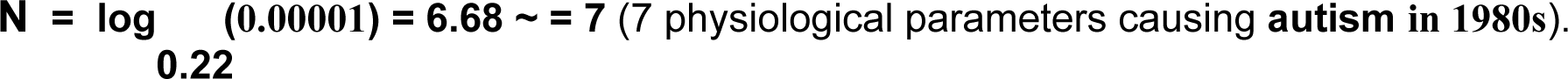

We see the number of physiological change in 1980s was **7** and now it is **3** (for girls) and even less it is **2** for boys. So this means some external factors systematically moved 4 additional parameters beyond 1-sigma (actually, slightly less) for entire populations so they are no longer protect children. According the article [1] **the less number of physiological parameter changes required to cause the non-infectious disease the more the incidence rate of the non-infectious disease** (this conclusion is coming directly from formula (1.0))

## Building a Hypothesis of Autism Pathology

As it was already shown in article [2] *(“Method of Combining Multiple Researches to Determine Non-Infectious Disease Causes, Analysis of Depression and Celiac Disease Causes”*, Alan Olan, Biology and Medicine,vol.16 Iss.1 No: 1000635) this method of determination of disease cause allows “connect the dots” of found physiological parameters changes for a disease and come up with hypothesis of the disease more easily. Let’s demonstrate it here on example of Autism.

We have found by the presented method that Autism is caused by next physiological changes beyond 1-sigma which should be present at the same time **”Insulin Resistance Increase”** beyond 1-sigma**, “Increase in TNF - Alfa”**beyond 1-sigma**, “Iron Deficiency”** beyond 1-sigma. Insulin Resistance in the brain, and insulin resistance is generally translate to brain insulin resistance, is expected to impair brain cell functioning. As a research by Cahill states: “**Maternal *insulin resistance* in obese women appears to negatively affect neurodevelopment at 2 years in their children** compared to children born to non-obese women.”. Also, regarding Insulin Resistance (IR) the research by Manco et al states: “**Brain IR has been associated with reduced executive functioning, inhibitory control, and cognitive flexibility in young patients** with obesity and implied in the pathogenesis of dementia in adults suffering T2D or Alzheimer disease.”

Based on this and other research we can hypothesize that an Insulin Resistance increase beyond 1-sigma in a child (or during the fetus development) reduces capabilities of brain cells to consume glucose which is critical for brain functioning and development. At the same time the disease causation factors which are causing an increase of **TNF-alfa** beyond 1-sigma such as “CNV + Ozone Exposure”, “Air pollution”, etc (see the Table 2.2) are also causing an increase in Insulin Resistance even more (based on a research by Alzamil et al, etc. which shows that TNF-alfa increases insulin resistance). In this case, the increased insulin resistance far beyond 1-sigma (due to TNF-alfa increase beyond 1-sigma as an additional factor) is reducing the ability of energy consumption of *high energy* demanding brain cells of speech and cognitive centers due to inability to acquire enough of glucose and other required nutrients in **boys.** The boys’ brain centers responsible for speech, and cognition are stopping development due to a significant lack of energy from glucose, etc. In girls, as we already hypothesized above, this will not happen because they have a compensating mechanism to address Insulin resistance which is a **controlling of Iron levels** and also because the Iron level serve as 3^rd^ physiological parameter which need to be changed according to the result found per the method. 3^rd^ parameter protects girls from Autism more.

As a fact we know from method results that **low Iron level** beyond 1-sigma is one cause of Autism and hypothesis just says that this physiological parameter is the reason the disease does not start in girls when 2 other physiological parameters already changed beyond 1-sigma.So to summarize, our **<u>hypotheses of Autism pathology is</u>**:

*An increase of insulin resistance in a child’s brain beyond 1-sigma due to known causation factors (see Table 2.3) is getting far more bigger due the existing increase of TNF-Alfa beyond 1-sigma due to another set of known causation factors and the total insulin resistance increase in the brain is causing significant lack of glucose and other nutrients required to satisfy high energy demands of <u>brain cognitive centers</u> in boys and this lack of energy stops the neurons functioning and neural development. This will not trigger development of Autism in girls until the Iron levels in girl’s (or girl’s fetus) drop beyond 1-sigma, which will stop a work of compensational mechanism for insulin resistance based on iron regulation*.

The low Iron level (beyond 1-sigma) is 3rd physiological parameter we have determined which is causing Autism in children if combined together with 2 others. As we already explained, the found physiological changes which are triggering Autism *cannot* cause it if are present standalone or not with **all** required physiological changes.

As readers can see we use a knowledge of physiological parameters causing a non-infectious disease (in this case Autism) discovered with the presented method along with a known research on these physiological parameters to build a meaningful relationships between the discovered physiological changes (“connecting the dots”) and thus building a hypothesis of non-infectious disease pathology.

## Mechanism of Brain Inflammation in Autism (proposed)

We have found that 1 cause of Autism is TNF-alfa increase beyond 1-sigma and the TNF-alfa increase in Autism is confirmed by existing medical research (Xie J et al) as it was discussed above. There is a question how can TNF-alfa level be sustained permanently in Autism?

Multiple research has found that TNF-alfa levels positively correlate with insulin levels.(*“The Heterogeneity of Diabetes: Unraveling a Dispute: Is Systemic Inflammation Related to Islet Autoimmunity?.”*, Massimo Pietropaolo, Emma Barinas-Mitchell,et al, Diabetes 1 May 2007; 56 (5): 1189–1197. https://doi.org/10.2337/db06-0880). As we have shown using the presented method the 2nd cause of autism is an **increased level of insulin resistance**, above 1-sigma. The insulin resistance that this high should in turn cause a compensatory increase of insulin level and the maternal insulin resistance beyond 1-sigma should also increase fetus glucose and insulin level as well as reported by Kullmann, at el *(“Brain Insulin Resistance at the Crossroads of Metabolic and Cognitive Disorders in Humans*“,Stephanie Kullmann,Martin Heni,et al,American Physiological Society,3 August 2016, https://doi.org/10.1152/physrev.00032.2015). The increased insulin level should correspond to an increased TNF-alfa level as they are positively correlated. The increase of TNF-alfa level (which is confirmed by research on Autism) should cause an increase of insulin resistance by acting on Insulin Receptor (IR). As research states: “**TNF-α by inducing serine phosphorylation of insulin receptor substrate-1 can act as an inhibitor of peripheral insulin action which leads to insulin resistance** “ (*“Elevated Serum TNF-α Is Related to Obesity in Type 2 Diabetes Mellitus and Is Associated with Glycemic Control and Insulin Resistance.”*, Alzamil H, J Obes. 2020 Jan 30;2020:5076858. doi: 10.1155/2020/5076858.)

This additional increase of Insulin Resistance in turn should correspond to even higher insulin level and it should correspond to even more increased level of TNF-alfa, etc. We get a viscous cycle leading to permanent inflammation.TNF-alfa is produced mainly by mononuclear phagocytes, antigen-stimulated T-cells; NK cells (Massimo Pietropaolo et al) and as research shows autistic children have an increased level of CD+8 T-cells: “children with autism had impaired cell mediated immunity as evidenced **by low numbers of CD4 T cells with an increased CD8 T cell with decreased CD4/CD8 ratio in comparison to the control children.” (*“****Cellular-mediated and humoral immunity in children with autism”*, Egypt J Pediatr Allergy Immunol 2012;10(1):25-32.)

We can suggest that part of these CD8 T-cells is activated by insulin as auto-antigen and these CD8 T-cells are participating in TNF-alfa production observed in Autism. The research by Eisenbarth, et al states: “**given this predominance of insulin it is not unexpected that autoimmunity directed at insulin is readily demonstrated in both man** and the NOD mouse.”*(“Insulin autoimmunity: prediction/precipitation/prevention type 1A diabetes”*, George S Eisenbarth, Hiroaki Moriyama, et al, Autoimmunity Reviews,Vol 1, Iss. 3, Vol.1, Issue 3, https://doi.org/10.1016/S1568-9972(02)00035-6.)

Another research on diabetes has found that CD8+ cells were found in non-diabetic controls (HC) and diabetic type I people: “**islet-autoreactive CD8+ T cells were present in both HCs** and individuals with diabetes”(*“Autoreactive CD8+ T cell exhaustion distinguishes subjects with slow type 1 diabetes progression”*, Alice E. Wiedeman,Virginia S. Muir, et alJ Clin Invest. 2020;130(1):480–490. https://doi.org/10.1172/JCI126595.). Both researches allow to suggest that it is ***likely that insulin can be auto-antigen in Autism***.

Also, a process of chronic inflammation which is observed in adipose tissue (Hotamisligil GS et al) is likely supported by the similar viscous cycle. The adipose tissues is known to have local chronic low grade inflammation, higher level of TNF-alfa and is also known to have an increased insulin resistance. The similarity of physiological changes (Insulin Resistance, TNF-alfa) in adipose tissue and Autism which both coincide with chronic inflammations and increased levels of TNF-alfa also suggest a common mechanism of the inflammation in both cases.

**Figure 2.3.**
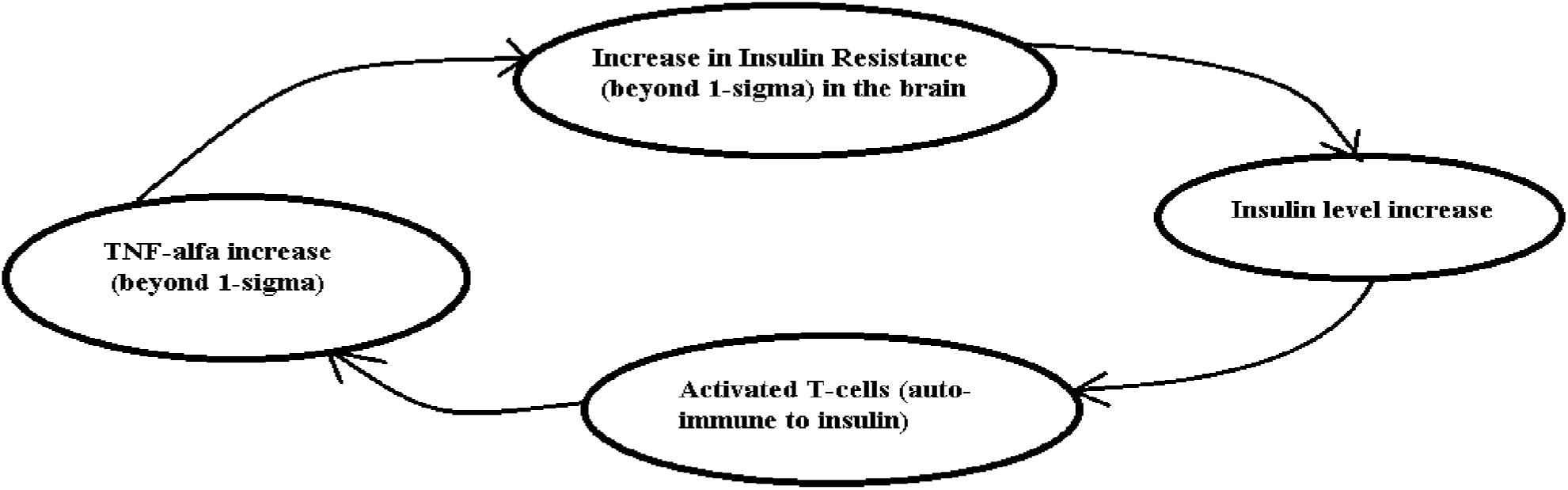
(Sustaining brain inflammation in Autism - proposed process. Increase of TNF-alfa adds up to already existing Insulin Resistance by acting on Insulin Receptors in the brain)

To summarize, we suggest that an increase of insulin level due an increased insulin resistance in Autistic children is expected to trigger some level of auto-immunity to insulin and the activated T-cells proliferation which in turn will be producing TNF-alfa, the increased level of which is observed in Autism. This would explain a mechanism of sustained inflammation in the brain observed in Autism and a permanent increase in TNF-alfa. In this case the **inflammation in the brain would be caused by a auto-immune reaction to an insulin and the higher the insulin level the more inflammation**. This is consistent with multiple research on improvement in Autistic symptoms in children on a ketogenic diet which reduces carbohydrates consumption, increases protein and fats consumption, which should *reduce* the level of glucose and insulin level accordingly (*“Ketogenic Diet and the Treatment of Autism Spectrum Disorder. ”,* Li Q, Liang J, Fu N, Han Y, Qin J. A, *Front Pediatr*, 2021 May 11;9:650624. doi: 10.3389/fped.2021.650624. PMID: 34046374; PMCID: PMC8146910.) and thus may reduce the level of auto-immunity and TNF-alfa and reduce Insulin Resistance in the brain as a result, etc. The diagram on Figure 2.3 demonstrate this *proposed* process which can be further investigated. If we are correct then ***Autism is mainly an increased insulin resistance of the brain which is supported by increased TNF-alfa due to auto immune reaction to the insulin*.**

As we already determined using the presented method Autism is caused by 3 simultaneously present physiological changes beyond 1-sigma which include changes beyond 1-sigma in **Insulin Resistance Increase** and an increase in **TNF-alfa**, beyond 1-sigma (in boys and girls) and additionally a **Low Iron**, beyond 1-sigma (in girls). This viscous cycle supporting inflammation in Autism which we’ve proposed, if confirmed is triggered with these physiological changes.

## Results and Discussion

We introduced a method of finding non-infectious disease causes using multiple researches which give estimates of risks (RR/OR) for different factors in regards to the *same* specific non-infectious disease. Method starts with finding a ***number of disease causes using a formula*** (1.0) when we know a rate of this non-infectious disease in a population (often we use an incidence rate for this to see annual probability of the disease).

According to the method, once the disease causation factors were found using ***disease causation criteria provided*** in this article above we need to find out physiological parameters which are changed when these factors are present. The disease causation factors are selected from *known risk factors* for a non-infectious disease using the disease causation criteria. Once we list all disease causing factors vertically and physiological parameters *related* to them horizontally (or vice versa) we mark the relationship between related factors and physiological parameters changed with a letter **“R”**. Then we find so called intersections between 2 disease causation factors - the places where 2 different disease causation factors are having same physiological parameters marked as related with letter “R”. The physiological parameters are same by **name** (not by value).

We mark the column(or row) in gray or other color to indicate an intersection. We need to find all intersections like this. The physiological parameters where intersections are happening we list as matrix **Pm**. This list or matrix **Pm** is a superset of the physiological parameters changes which are causing a disease if their changes go beyond 1-sigma interval (actually, slightly less than this interval) and are present at the **same time.** By a superset we mean here that some parameters in a matrix **Pm** are disease causing but some may be just related to them and need to be eliminated (we call them such parameters *redundant*).

We eliminate the redundant parameters from this matrix **Pm** by using *few rules of elimination* and following a recommended algorithm if the count of parameters found exceeds the count determined by formula (1.0) calculating the number of disease causes or if a strict rule that a factor should cause only 1 change (or as determined by Disease Causation criteria) is broken.

One of these elimination rules is to check if the disease causation factors where intersection happens are related to each other (for example the same type of disease, or similar experiment like BMI and weight impact on disease, etc) and ignore the intersection for these factors as they are likely due to similarity. Another rule is to check if the physiological parameters for the same factors are related and this causing a change in 2 physiological parameters for the same disease causing factor as one valid physiological parameter is changing a dependent one. The dependent parameter need be eliminated from matrix **Pm**. Remember that most disease causation factors in practice are impacting only 1 physiological parameter change which is causing a disease. If we see a disease causation factor for which we determined only 1 physiological parameter change was found to be causing 2 or more changes in physiological parameters in our table (see example in Table 1.1 or Table 2.1) then the redundant physiological parameters need to be eliminated using the elimination rules either from matrix **Pm** or from the table.

Once the redundant parameters were eliminated we can check the final list of physiological parameters or matrix **Pm** using our *criteria for disease causing factors* (provided above for your reference) if there is an appropriate research regarding risks of these physiological parameters for the disease and confirm that we did not make any errors in steps of the described method. Also, we can use a related research regarding these found physiological parameters to see significance of their impact on the disease and this way to confirm correctness of method’s application and the significance of the found physiological parameters found via the described method.

As example of method’s application we analyzed 2 non-infectious diseases. One was Breast Cancer and another was Autism. We found that according to the method a Breast Cancer is caused by 4 physiological changes beyond approximately 1-sigma interval if **they present at the same time**. The disease must happen if the disease causing condition is satisfied for some time. We notice that Breast Cancer should *not* happen if only 1 of these parameters is beyond 1-sigma interval. We determined that **the physiological parameters which are causing Breast Cancer if they changed beyond 1-sigma (actually, slightly less) are:**

1. Increased DNA strand breaks (beyond ∼ 1-sigma)
2. TNF-alfa increased beyond ∼ 1-sigma
3. Leptin level **increase** beyond ∼ 1-sigma
4. High estradiol (above ∼1-sigma)

By controlling 4(or only few of them) of these disease causation physiological changes we can prevent Breast Cancer in an individual or possibly cure (if a disease in early stages) or treat it. If all the required physiological parameters changes take place at the same time the **Breast Cancer *must*** start after some time unless they are taken under control by forcing them to be within 1-sigma interval. In a healthy individual at least 1 or likely more of these physiological parameters need be close to a mean value (be within 1-sigma interval).

We have determined a connection between different disease causation factors for Breast Cancer and these 4 physiological parameters (see Table 1.2). From the Table 1.2 we can conclude that “Low Total Carotene” disease causation factor for Breast Cancer is causing an increase in DNA strand breaks beyond 1-sigma and that is why it is 1 of few disease causation factors for Breast Cancer. The same impact has genetic mutations in BRCA1, BRCA2 genes as they also increase DNA strand breaks beyond 1-sigma. Cystic breast disease is one cause of Breast Cancer (see Table 1.2) because it impacts TNF-alfa so it increases beyond 1-sigma. These factors mentioned will *not* cause Breast Cancer as standalone as they change only 1 physiological parameters beyond 1-sigma but to trigger Breast Cancer all 4 physiological parameters are required. **The disease causation mechanism for Breast Cancer resembles a padlock with 4 wheels**. If you turn 1 wheel the padlock will not open as you need to turn properly *all* 4 wheels. Once such a “padlock” open the disease will start. It must start no matter which disease causation factor “turned” the required wheel. The disease start because all the “wheels” turned properly and the factors are only the means to do it.

In the next step of the method we also divided disease causation factors for Breast Cancer in the groups according to a related physiological parameter they change beyond 1-sigma (see Table 1.3). The presence of 1 such disease causation factor from each of these groups *must* trigger Breast Cancer in an individual if present together long enough. Using the **Table 1.3** we can conclude, for example, that **if a women has Low Total Carotenes AND has Cystic Breast disease and High Breast Density (BI-RADS code 4) AND she is Postmenopause with Weight increased, BMI > 31.1 such a women *must* a Breast Cancer** if she continuously has these factors. This happens because each of these factors impacts 4 physiological changes shown above so they go beyond 1-sigma. As we already mentioned if we eliminate an impact to at least 1 or better more physiological changes out these 4 provided above, either by removing presence of disease causation factor or by making sure that the physiological parameter it impacts stays within 1-sigma, we should be able to prevent Breast Cancer, treat it or possibly cure (in early stages).

The method allows to build easier a hypothesis of disease pathology using the physiological parameters we determined and we showed this on example of building hypothesis of Breast Cancer pathology.

**Breast Cancer pathology hypothesis:** An increased beyond 1-sigma a number of DNA strand breaks (caused by such factors as low total carotene, etc) cannot be repaired due increased beyond 1-sigma TNF-alfa level (caused by Cystic Breast disease, for example) and an increased beyond 1-sigma level of leptin (caused by postmenopause women weight increase, BMI > 31.1, for example) where a leptin level increase reduces energy capabilities of breast cell in particular for repair processes and along with additional level of DNA mutations due to adducts created by high estradiol level beyond 1-sigma deprive the breast cell from the DNA repair capabilities required to compensate for a normal DNA degradation processes and DNA damages and mutation accumulate beyond control. This results in triggering of Breast Cancer. In short, **the disruption of DNA damage**/ repair homeostasis leads to a breast cancer.

In another example of method’s application we analyzed Autism. We determined that **Autism happens due to 2 physiological parameters changes** beyond 1-sigma interval **in boys** and due **3 physiological parameters** changes beyond 1-sigma **in girls** (according to the method and a mathematical model it is using [1]). The disease must happen if the disease causing condition below is satisfied for some time. We have determined that **the physiological parameters changes beyond 1-sigma (actually, slightly less) which are causing Autism if they are simultaneously present long enough are:**

1. Insulin Resistance increase beyond ∼1-sigma
2. Increase in TNF - Alfa beyond ∼1-sigma
3. Iron Deficiency (low iron) beyond ∼1-sigma (likely, for girls only)

That means if a child has Insulin Resistance (beyond ∼1-sigma) **and** has an Increase in TNF - Alfa(beyond∼1-sigma) **and** has Low Iron (beyond ∼1-sigma, for girls only likely) long enough the child must get Autism. It is *not optional* if Disease Causation conditions are satisfied and kept long enough according to the article [1].By controlling 2 (for boys) or 3 (for girls) of these disease causation physiological changes within 1-sigma interval we can prevent Autism in an individual, treat it or possibly cure it (if a disease in early stages).

We have determined the set of disease causation factors which if combined together must cause Autism (Table 2.2) and we found a **connection between these disease causation factors and these 3 physiological changes beyond 1-sigma** which are causing Autism. According to the Table 2.2 one of disease causation factors for Autism is “Valrpoate usage” as it causing insulin resistance increase beyond beyond 1-sigma in **individual** (it does not mean that a mean value in the group should necessary change beyond 1-sigma though). Another disease causation factor for Autism is “Fever after 12 week of pregnancies or High fever while pregnant” as it impacts Increase in TNF-alfa beyond 1-sigma. We also found that “Father over 45+” at the time of conception is a disease causation factor for Autism and we determined it is very likely happens because it is causing an Increase in TNF-alfa (which is a normal immunological reaction) beyond-1 sigma in pregnant women due to multiple errors in chromosomes which add up with the age in men’s sperm due to multiple replications. This multiple errors cause a stronger immunological reaction in pregnant women.

Based on the knowledge of these 3 physiological causes of Autism we also suggested that an increase of insulin level due an increased insulin resistance in Autistic children brain is expected to trigger some level of auto-immunity to insulin and the activated T-cells proliferation which in turn will be producing TNF-alfa, the increased level of which is observed in Autism. This would explain a mechanism of sustained inflammation in the brain observed in Autism and a permanent increase in TNF-alfa. In this case the **inflammation in the brain observed in Autism would be caused by a auto-immune reaction to an insulin** and the higher the insulin level the more inflammation. If we are correct, then **Autism is mainly an increased insulin resistance of the brain**(beyond 1-sigma) **which is supported by increased TNF-alfa**(beyond-1 sigma) due to **auto immune reaction to the insulin**. This is consistent with a research that a ketonogenic diet improves Autistic symptoms (as shown by Li Q et al and others) and the TNF-alfa increase and Insulin Resistance were found to be causes of Autism as shown above.

We have built **groups of disease causation factors for Autism (Table 2.3) which combination from each column must trigger Autism** if these factors present long enough in an individual (it is not optional). One combination of disease causation factors for Autism is “Fever after 12 week of pregnancy or HIGH fever while pregnant” (increases TNF-alfa) and “Depression” in women (increase Insulin Resistance) and “Child was born during 1^st^ year after previous pregnancy” (reduces Iron). The Low Iron as we have determined mostly should impact girls and should be 3rd physiological parameter which is needed to cause Autism in girls. The **normal Iron level (within 1-sigma) should be protecting girls** from such a strong impact which Autism is having on boys. As stated in article [1] the more physiological changes required to cause a non-infectious disease the less the incidence rate of the disease. In this case an addition of 3^rd^ physiological parameter which need to be changed to cause Autism in girls protect them and significantly reduces an incidence rate of Autism in girls.

We have shown that **incidence of Autism has grown last few decades** because some disease causation factors moved 3-4 other *unknown* yet physiological parameters changes beyond 1-sigma for majority of children (to a new “norm”) and this has reduced a number of physiological protections for Autism and it is causing the increased level of Autism. In 1980s Autism as we determined using the method was caused by 7 physiological parameters changes and and now it is caused by 3 changes(in girls) and 2 changes in boys as was also shown in article [1]. As we already mentioned that according to article [1] **the less number of physiological parameters required to cause a disease the more incidence rate of non-infectious disease.**

We hypothesized that **Iron level reduction** which is one of 3 determined causes of Autism **is the same mechanism** (which in this case broken) which **compensate for increased Insulin Resistance in girls** (or girls’ fetuses during normal pregnancies as shown in one research of pregnancies cited in this article) and what it is an additional physiological parameter required to cause Autism in girls.

In this article we also showed how the method allows to build a hypothesis of non-infectious disease using the set of found physiological parameters which cause the disease and as an example came up with hypothesis of Breasts Cancer and Autism pathologies which are based on the physiological causes we discovered.

**Hypothesis of Autism is**: An **increase of insulin resistance in child’s brain beyond 1-sigma** due to known causation factors(see Table 2.2) **is getting far more bigger due the increase of TNF-Alfa beyond 1-sigma** due to another set of known causation factors(see Table 2.2) **and the total insulin resistance increase is causing significant lack of glucose**, and other nutrients required to satisfy *high energy demands* of brain cognitive centers in boys and this lack of energy stops the neurons functioning and neural development. This will not trigger development of Autism in girls **until the Iron levels in girls (or girl’s fetus) drops** beyond 1-sigma interval, which will stop a work of compensational mechanism for insulin resistance based on iron regulation. The disease causation factors in this hypothesis were determined in this work and provided in Methodology section (Table 2.2, Table 2.3)

The **triggering of the non-infectious disease if the causation conditions are satisfied is not optional.** It is a must.The analogy of this process could be an electrical fuse in a house. We know that if we plug in an electrical iron, air conditioner, etc. and exceed the threshold for the fuse the fuse will burn out. The fuse must burn out, it is not optional. The only random facts here are the electrical appliances which will cause it, these electrical appliances are an analogy of factors causing diseases here. We can have different external factors causing a non-infectious disease but as long as they impact all required physiological parameters a “fuse” of the body will burn out and the disease will start.

We need to notice here that multiple medical researches **often find risk factors for a specific disease which have values close to RR/OR = 4.6 or RR/OR = 20.6**. These are not just a statistically significant risks but these numbers indicate that 1) the **factors are really disease causing factors** 2) the factors are **causing a change in 1 physiological parameter beyond 1-sigma** (for RR/OR = 4.6) or **2 physiological parameters beyond 1-sigma** (for RR/OR = 20.6). One example of these cases was shown in this article as an alcohol consumption by women between 50-59 age (**RR = 18**) in **Table 1.2** and which is a disease causation factor for a Breast Cancer which impacts 2 physiological changes, etc. In some rare cases the risk factors can have **RR/OR=93**+/-50% and this means the disease causing factors are impacting **3 physiological parameters beyond 1-sigma.** The disease causation criteria provided in article *“A Connection between Factors Causing Diseases and Diseases Frequencies: Its Application in Finding Disease Causes***”** (*Alan Olan,* Journal of Clinical Trials, Vol.13, Issue 4) allows to determine more precisely if the risk factors are causing a disease or not and allows to expand validity for these risks beyond the 2 examples of RR/OR just mentioned above. The referred article is using a risk factor **K** but it is easily converted to RR/OR as **RR = K + 1** (same for OR) and the RR/OR can be used as the theoretical values for RR/OR +/-50%. A short reference of these criterions was provided in this article as well in Introduction section.

In this article we have shown that **disease causation factors can be separated into few groups** according to a physiological parameter they change beyond 1-sigma. **Any combination of factors** taken as one from each of these groups **must cause a non-infectious disease.** The real causes of the non-infection disease is a set of changes of certain physiological parameters beyond 1-sigma (slightly less, actually) and the disease causation factors are **only means** to make these changes. There can be many different disease causation factors but **only few (2 or more) physiological changes beyond 1-sigma** which are required to **cause a non-infectious disease.**

## Principle of Indifference

As we discuss a method to find disease causes via experiments we are proposing to use a simple principle which can be observed in physiological processes. **When regulating its own homeostasis separate physiological systems of the body are indifferent to the side effects of this regulation.** We can call this *a principle of indifference*.

We can observe this principle in multiple cases. For example, if a brain needs to increase a blood pressure to improve a supply of nutrients and oxygen it is *indifferent* to the fact that a heart may not be able to sustain this high blood pressure. Another example could be this. If an intestine is trying to regulate its homeostasis of bacteria it can pass the signals to the brain via different biochemical pathways to reduce appetite so in this case an intestine is *indifferent* to that its actions can eliminate some source of energy and needed nutrients to the brain and other systems and worsen their functioning.

This principle allows to explain that supporting a homeostasis in one physiological system of the body can and often does harm functioning of other systems and this way it can cause a non-infectious diseases. If we find a physiological system which is regulating its homeostasis with extreme steps and help it to fix the problems we can address a non-infectious disease of another physiological system which can be caused by this indifference.

## Conclusions

In this article we have showed how a research which estimate a risk of non-infectious disease can be used to determine disease cause as a set of physiological parameters changes beyond 1-sigma (actually, slightly less than this interval). We showed how to determine which physiological parameter is impacted by a specific external factors. It was shown how certain different combinations of external factors can cause a disease if they act together and how to determine these disease causing factors combinations.

Overall a method presented here allows to increase efficiency of existing medical research in finding non-infectious disease causes and also stresses that non-infectious disease are having a multi-factorial causes. These causes are multiple physiological parameters changes beyond 1-sigma occurring at the same time and happening due to actions of external disease causing factors.

by Alan Olan - 2024

**Disclaimer:** This work is not intended to be used in diagnosing and treatment of any disease in any patient, those actions should be done by qualified professionals only. The work is intended as a reference for the researchers only. The author is not responsible for any issues created by unintentional usage or consequences of this work.

## Ethical Compliance

All procedures performed in studies involving human participants were in accordance with the ethical standards of the institutional and/or national research committee and with the 1964 Helsinki Declaration and its later amendments or comparable ethical standards.

## Data Availability

All data produced in the present work are contained in the manuscript

## APPENDIX

### 1. Analogy to clarify a method

A **simplified analogy** which explains why we looking for a match between physiological parameters could be this. Imagine a large town. We observe it from the top. A person arrives to downtown for some *personal business* on a regular day. How likely this person meet *a friend* or colleague in a totally random place of the downtown?..Very unlikely. We observe this person is going to downtown for few days to different places and he never meets a friend or a colleague. Now, we observe from the top that some rare folks meet someone in downtown often and sometime in the same location. We know **there is a pattern explaining these meetings**, they are not random in most cases. There might be someone they have agreed to meet with before (a colleague they travel together with, a friend, etc). If we find these folks meeting we know we found very likely some pattern.

In this analogy, we can treat a downtown as a human body, a person arriving to downtown as a physiological parameter change caused by some disease causing factor. People which meet each other in the downtown are an analogy of physiological parameters which “meet up” as they cause a disease and not just a random meeting. If we find those folks who meet up we know there is some cause there. In the method presented, these “meetings” between physiological parameters are represented by “intersections”.

### 2. Mathematical foundation of the method

Here we will go into a mathematical foundation of the method in details.Let’s look at the simple case of a disease where it was prior determined via experiments that multiple factors are causing changes in only 2 physiological parameters of human body (parameters further).

Let the factors be **F1, F2, … Fn** and the parameters be **C1, C2.** Now, let’s look at the case where *F1, F2,…, Fn factors separately causing **only 1** change* in physiological parameters either C1 or C2 beyond 1-sigma (this actually often takes place in practice). That means that only C1 *or* C2 changes by some factor Fj (j E {1,2,..n }). Let’s **P1, …, Pn**, where n > 2 be *the sets of all physiological parameters which are related to factors F1, F2,.. Fn* accordingly. For example, P1: { r12, r15, r29, 43 }, P2: { r15, r28, r34, r89, r34, r12, r98 }, etc.

Let’s look at standalone factor Fj. As we know, a factor Fj (where *j is some integer from 1 to n*) impacts the specific physiological parameters either C1 or C2 then we know that *this params C1 or C2 should be part of its set of* Pj as it contains ALL the related to Fj parameters (a complete set). *Let’s take a factor F1 such that its set P1 contains C1, and choose some F2 such that its set P2 contains C2* (it is possible as we know factors impact either C1 or C2*), then if we choose any other factor as F3 then its set of P3 must contain either C1 or C2 (as F3 also impacts these physiological parameters - either C1 or C2 and P3 is a complete set). If P3 contains C1 then it intersect with P1. If P3 contains a C2 it intersects with P2. So P3 must intersect with either P1 or P2 (either in C1 or C2). In similar way we can apply this to P4, P5,… Pn. So this brings us to conclusion that a set of physiological parameters Pn, where n > 2 must intersect with either P1 or P2 either in C1 or C2. This means parameters Pn intersect with each other either in C1 or C2.* We can see representation of this set’s behavior on the Pic 2. *All sets Pj, Pk,… matching to Fj, Fk,.. on the Pic 1 are crossing and only in C1 or C2 but not both*.

**Pic 2.**
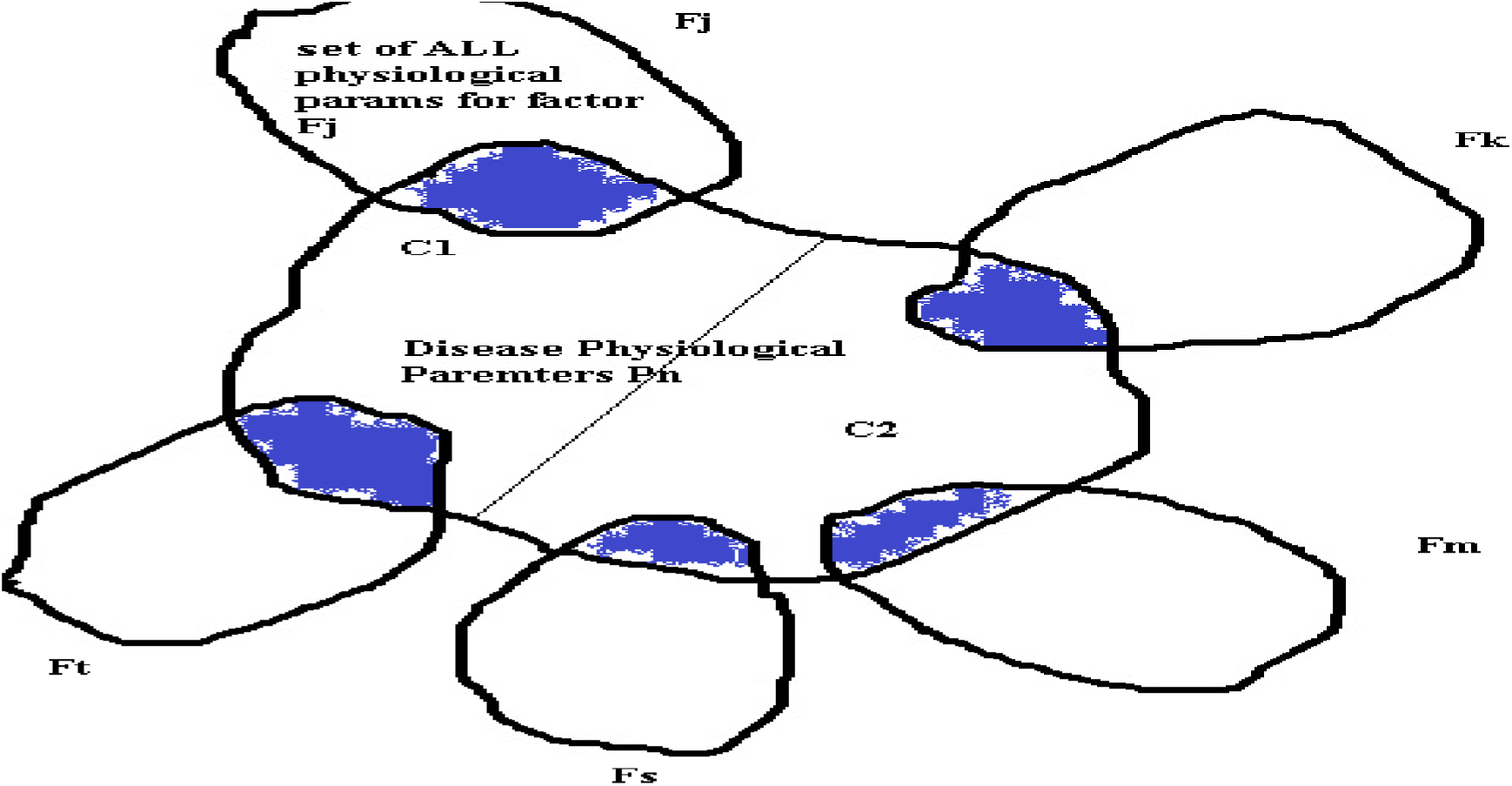
(Blue areas are area where Pj, Pk, etc for factors Fj, Fk, etc. intersect with a set of physiological parameters C1, C2 which are part of set Pn and which are causing diease)

We don’t know the values of C1 and C2 but if *we can find where parameters Pn intersects with each other we can determine a subset of physiological parameters Pm:* { Ry, Rx, Rz,.., Rt } which contains values of C1 and C2. This subset of Pm will be much smaller then set of all possible params included in P1, P2, .. Pn (as it is a subset and similarities in params of P1,.. Pn are not very probable and that is addressed below) but may contain more then 2 parameters and *only 2 parameters of this subset Pm* can be real physiological parameters causing a disease as they are C1 and C2.

In order to eliminate the incorrect parameters from subset Pm we need to notice:

1) that the params C1 or C2 should *be such so all P1, P2, …, Pn intersect in them* and if some parameters of *Pm:* { Ry, Rx, Rz,.. Rt } don’t fit this rule *their need to be eliminated.* Practically it means this. We take random (or using a common sense) a combination of some 2 parameters Rk and Rm from a set of Pm *and check if the P1, … P2 all intersect in them if not then the Rk and Rm combination is not a valid set of C1, C2 and we may need to check another set of 2 parameters Rk and Rg*
2) if some parameter of set *Pm:* { Ry, Rx, Rz,.. Rt } *is known as not changed beyond 1-sigma it should be eliminated* as disease is caused by change in param beyond 1-sigma (as per our model).
3) if some parameter of *Pm:* { Ry, Rx, Rz,.. Rt } *is causing some set Pn intersect 2 times with some other set Pk then it should be eliminated* as factors F1, F2, .. Fn can *only* impact 1 physiological parameter in this case and cannot impact / intersect 2 or more due to this.

The method above was described for a case of factors F1, …, Fn impacting *only 2* parameters but it can be extended to 3 and more parameters.

### How likely are random matches between physiological parameters?

As we discussed above the set of physiological parameters *Pm:* { Ry, Rx, Rz,.., Rt } where we observe intersections may contain more parameters then needed (more than 2 in our case and due to other reasons).

We need to be concerned with a question such as if we find one intersection of sets P1 and P2 in a physiological parameter belonging to 2 different external factors how likely it can be a random intersection? To answer this question let’s formulate the problem mathematically.

Let’s have a set A of integers from k = 1 to very large N. Let’s randomly select n numbers in set of P1 = {Ak, Ag, .., At } and then randomly select n numbers into set P2 = { Af, As, … Al } from our original set A (k= 1 to N) such that each element repeats only once in set P1 and only once in P2 (it is a unique element to sets P1, P2). For example, if we chose a number 3 as part of the set P1 then it only exist one time in the set P1. What is a probability that we find element **Ai** in set P1 and P2 ?

To answer this question let’s do next steps. Let’s limit set A by some top element enumerated by t (so set is not infinite).

1. We can take **n** elements from **t** elements of set A with number ways **tCn**
2. Number of ways to take **n** elements with an element **Ai** equals the number of ways to select **n-1** elements (we exclude Ai) from **t -1** (set of A elements) and is **t-1Cn-1**
3. Then probability to take n elements which include element **Ai** in set P1 (or P2) is P (Ai E Psel) = **t-1Cn-1 / tCn**, where Psel is P1 or P2 sets
4. The probablity that element **Ai** will be in P1 and P2 is P (Ai E P1 and **Ai** E P2) = P (Ai E P1) * P (Ai E P2) as events independent
5. So probability P (Ai E P1 and **Ai** E P2) = P (Ai E P1) * P (Ai E P2) = (**t-1Cn-1 / tCn**) ^ **2**
6. Or finally, the probablity that element **Ai** wil be in P1 and P2 is P (Ai E P1 and **Ai** E P2) = (**t-1Cn-1 / tCn**) ^ **2**

Using a formula above let’s calculate a probability of match in element **Ai** if we take randomly elements from a sequence of numbers from 1 to 1000 (t = 1000, assuming so many physiological parameters exist) and take only n = 10 element into sets P1 and P2 accordinly. P (Ai E P1) = 999C9 / 1000C10 = (2.63 * 10^21) / (2.63 * 10^23) = 1 / 10^2 = 0.01 the same is fare for P (Ai E P2) = 0.01 and so the probability of getting element **Ai** in sets P1 and P2 is

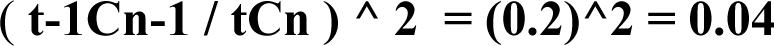

This is a probability of radom match. The probability of non-random match is 1 - P (Ai E P1 and **Ai** E P2) = 1 - 0.0001 = 0.9999 ∼= 1 so very close to 1. It means ***if we see a match between set P1 and set P2 in some element Ai it exremely likely it is not random.*** This is an important conclusion. *The only matches we find practically are not random but are caused by some reason* and in our case it is due to same physiological parameter impacted by 2 different factors. We need to notice that number of paramaters actaully much more as most agree that there are around 20,000 different proteins in our body and each is a potential physiological parameter. So the probability of the match selecting 10 of 20,000 will be much smaller!

In practice there are about over t=150 physiological parameters known to medicine and for a single factor we usually find about n=30 related paramters. Doing calculations for this case we get P (Ai E P1) = 149C29 / 150C30 = (6.43 * 10^30) / (3.2 * 10^31) = 0,2 and so the probability of getting element **Ai** in sets P1 and P2 is

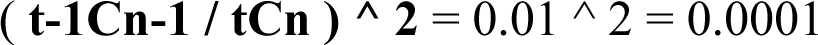

We see the probability of random match is higher in practice (4%) and so in practice we can see more random matches. The probability of at least one random match will also increases as we find intersections for dozens of different causation factors.

This random matches still can be eliminated with methods described in this article by applying other restrictive conditions, including using our criteria for disease causes.

